# Beyond tumor volume: connectomic measures of tumor burden provide superior prognostic information across tumor compartments in glioblastoma

**DOI:** 10.64898/2026.09.14.26362015

**Authors:** Joan Falcó-Roget, Augusto Ielo, Anna Janus, Sara Lillo, Alfonso Fasano, Michela Matteoli, Federico Pessina, Letterio S. Politi, Ludovico Coletta, Laura Vavassori, Silvio Sarubbo, Paolo Avesani, Jan K. Argasinski, Alberto Cacciola

**Affiliations:** Computational Neuroscience Group, Sano Centre for Computational Medicine, Kraków, Poland; IRCCS Centro Neurolesi Bonino Pulejo, Messina, Italy; Department of Neurophysiology and Chronobiology, Institute of Zoology and Biomedical Research, Faculty of Biology, Jagiellonian University, Krakow, Poland; Radiation Oncology Unit, Clinical Department, National Center for Oncological Hadrontherapy (CNAO), Pavia, Italy; Department of Internal Medicine and Therapeutics, University of Pavia, Pavia, Italy; Department of Biomedical Sciences, Humanitas University, Via Rita Levi Montalcini 4, Pieve Emanuele, 20072 Milan, Italy; IRCCS Humanitas Research Hospital, Via Alessandro Manzoni 56, Rozzano, 20089 Milan, Italy; Neuroinformatics Laboratory (NiLab), Bruno Kessler Foundation (FBK), Trento, Italy; McConnell Brain Imaging Centre (BIC) and Centre of Excellence in Epilepsy at The Neuro (CEEN), Montreal Neurological Institute, McGill University, Montreal, Quebec, Canada; Department of Neurosurgery, Azienda Provinciale per i Servizi Sanitari (APSS), “S. Chiara” University-Hospital, Trento, Italy; Department of Cellular, Computational and Integrative Biology (CIBIO), Center for Medical Sciences (CISMed), Center for Mind and Brain Sciences (CIMeC), University of Trento, Trento, Italy; Faculty of Physics, Astronomy and Applied Computer Science, Jagiellonian University, Krakow, Poland

**Keywords:** *glioblastoma*, *survival*, *tumor volume*, *tractography*, *connectomics*

## Abstract

**Background:** Glioblastoma (GBM) interacts with distributed brain networks, suggesting that its prognostic impact may extend beyond local tumor burden, yet imaging-based prognostic assessment remains largely focused on tumor volume. Despite more than 30 years of research, the association between tumor volume and survival remains inconsistent. Brain-wide tractography-derived measures may capture additional prognostic information, but their robustness remains uncertain.

**Methods:** We analyzed 999 patients with newly diagnosed GBM pooled from multiple cohorts. For each of five tumor compartments, we quantified volume and derived tract-density measures by mapping the compartment onto a normative tractogram. Associations with overall survival (OS) were evaluated using log-rank tests across stratification percentiles and survival horizons, with the Benjamini–Bogomolov procedure accounting for multiple testing within and across compartments. Cox models assessed associations after adjustment for clinical covariates.

**Results:** Associations between tumor volume and OS varied across compartments and stratification thresholds, with only contrast-enhancing volume consistently associated with survival. Tract-density measures showed more consistent associations across survival horizons, tissue compartments, and stratification thresholds. The whole-tumor tract-density measure yielded the most parsimonious model and remained associated with OS after adjustment for age, MGMT promoter methylation status, and extent of resection.

**Conclusions:** The prognostic value of tumor volume in GBM depends strongly on the compartment assessed and the stratification threshold. Brain-wide tractography-derived measures provide more consistent survival associations across compartments and retain prognostic information after adjustment for established clinical factors. These findings support moving beyond local volumetric assessment to incorporate connectomic biomarkers into prognostic modeling in GBM.

**Key Points:**

- Tumor volume shows inconsistent prognostic value for GBM survival, except for the contrast-enhancement volume.
- A brain-wide tractography-derived measure, the lesion-tract density index, shows robust survival associations across tumor compartments.
- Whole-tumor lesion-tract density yields the most parsimonious survival model.

## 1. Introduction

Glioblastoma (GBM) remains the most aggressive primary brain tumor in adults, with median overall survival rarely exceeding 15 months despite maximal multimodal treatment^1–3^. Reliable preoperative biomarkers capable of stratifying patient prognosis are therefore essential for surgical planning, clinical trial design, and personalized therapeutic decision-making. Magnetic resonance imaging (MRI) plays a central role in this effort by providing in vivo characterization of tumor burden and its impact on brain structure and function, while being routinely used in clinical practice.

Among imaging-derived biomarkers, preoperative tumor volume has been investigated for more than three decades as a potential predictor of survival^4^. Advances in image analysis have transformed volumetric assessment from simple two-dimensional approximations to automated three-dimensional segmentation of biologically distinct tumor compartments, including contrast-enhancing tumor, necrosis, and peritumoral non-enhancing abnormalities^5,6^. Yet, despite extensive study, the prognostic significance of preoperative tumor volume remains surprisingly uncertain^7^. Reported associations with survival vary substantially across cohorts, tumor compartments, and statistical designs, as well as analytical and numerical frameworks. This naturally raises questions about whether volumetric measurements capture clinically meaningful aspects of disease biology.

Regardless of this variability, GBM volume measurements are routinely integrated into clinical and radiomic frameworks^5,8^, with the expectation that powerful modern machine learning and deep learning models integrate multiple imaging features and identify prognostically informative patterns that may not be captured by individual biomarkers^9^. Tumor volume contains biological information, but as a macroscopic biomarker, it may lack specificity and might instead be an epiphenomenon of several more fundamental biological mechanisms^10,11^. Indeed, the latest preclinical investigations have uncovered a complex neuron-glioma system^12^ in which neuronal activity promotes tumour growth, thus identifying tumour–brain interactions as potential new therapeutic targets^13^. Because these interactions occur within the structural and functional architecture of the brain, they are unlikely to be explained solely by local cellular mechanisms. Instead, they suggest that glioma behaviour is shaped by the organization of large-scale brain networks, motivating a connectomics-based framework for understanding tumour evolution^14,15^. Connectomics is a relatively new neuroimaging framework capable of providing quantitative estimates of the brain’s multiscale architecture to inform estimates of patient survival^16–21^ and neurobiology^22–33^, as well as surgical and neurological outcomes^34–39^.

In particular, diffusion MRI tractography offers a unique opportunity to quantify structural interactions between gliomas and white matter pathways, generating imaging biomarkers that extend beyond conventional anatomical measurements. Tractography-derived indices condense the complexity of white matter organization into clinically interpretable measures of tumor-network interaction. Recent approaches have focused on either local^40,41^ or widespread white matter involvement^42,43^, with the latter showing superior prognostic performance. However, whether these connectomic biomarkers provide clinically meaningful information beyond conventional imaging markers remains unsettled. Existing studies have not systematically compared tractography-derived indices with standard volumetric measures across tumor compartments and different statistical modelling frameworks. Consequently, establishing the incremental prognostic value of structural network biomarkers is an important prerequisite for their clinical translation.

Here, in a cohort of 999 patients with newly diagnosed IDH-wildtype GBM, we systematically compared conventional volumetric measurements against the Lesion–Tract Density Index (L-TDI), a connectomic measure of structural brain-tumor interactions. Our analysis across tumor compartments, stratification thresholds, and survival horizons reveals that GBM prognosis is more consistently associated with tumor involvement of brain-wide white-matter architecture than with local tumor volume. These findings suggest that tractography-derived biomarkers show more consistent prognostic associations than conventional volumetry across statistical designs, highlighting the potential value of brain-wide measures of tumor involvement for GBM prognostication and understanding.

## 2. Methods

### 2.1. Imaging, clinical, and survival data

In this study, four retrospective, independent, and publicly available cohorts comprising a total of N=999 patients were employed. All the GBM patients included in the final sample were confirmed IDH-wildtype, following the WHO 2021 classification. Clinical characteristics can be found in Table 1.

**Table 1.** Sample characteristics and available measurements for every cohort and the total sample. The number of C+E.size and C+E.L-TDI need not coincide with the number of samples available for each tumor compartment alone, since C+E used both necrosis and contrast-enhancing tissue when available, or one of both in the case of one missing. Note that the preoperative KPS performance status was included for transparency but not used in any of the following analyses due to a significant reduction in the sample size. The percentage of right censoring, neighboring 20%, did not compromise the stability of the survival estimates. See also Fig. 1b-e.

| Feature | UCSF | UPENN | TCGA | RHUH | Total |
| --- | --- | --- | --- | --- | --- |
| Age (%) | 367 (100) | 496 (100) | 103 (100) | 33 (100) | 999 (100) |
| [years ± std] | [62 ± 12] | [63 ± 12] | [59 ± 13] | [64 ± 9] | [62 ± 12] |
| Sex (%) | 367 (100) | 496 (100) | 103 (100) | 33 (100) | 999 (100) |
| [M/F] | [218/149] | [297/199] | [63/40] | [22/11] | [600/399] |
| KPS performance status (%) | 0 (0) | 75 (15.1) | 85 (82.5) | 33 (100) | 193 (19.3) |
| MGMT status (%) | 355 (96.7) | 279 (56.3) | 65 (63.1) | 0 (0) | 699 (70) |
| [-/intermediate/+] | [104/5/246] | [151/25/103] | [37/0/28] | [0/0] | [292/30/377] |
| EOR (%) | 367 (100) | 472 (94.6) | 0 (0) | 33 (100) | 872 (87.3) |
| [biopsy/STR/GTR] | [41/112/214] | [0/180/292] | [0/0/0] | [0/0/33] | [41/292/539] |
| Right censored (%) | 144 (39.2) | 14 (2.8) | 21 (20.4) | 9 (27.3) | 188 (18.8) |
| Identifiable W.size (%) | 367 (100) | 496 (100) | 103 (100) | 33 (100) | 999 (100) |
| Identifiable C.size (%) | 363 (98.9) | 494 (99.6) | 103 (100) | 33 (100) | 993 (99.4) |
| Identifiable E.size (%) | 367 (100) | 493 (99.4) | 102 (99) | 33 (100) | 995 (99.6) |
| Identifiable NE.size (%) | 367 (100) | 496 (100) | 103 (100) | 33 (100) | 999 (100) |
| Identifiable C+E.size (%) | 367 (100) | 494 (99.6) | 103 (100) | 33 (100) | 997 (99.8) |
| Identifiable W.L-TDI (%) | 367 (100) | 496 (100) | 103 (100) | 33 (100) | 999 (100) |
| Identifiable C. L-TDI (%) | 355 (96.7) | 493 (99.4) | 103 (100) | 33 (100) | 984 (98.5) |
| Identifiable E. L-TDI (%) | 367 (100) | 493 (99.4) | 102 (99) | 33 (100) | 995 (99.6) |
| Identifiable NE. L-TDI (%) | 367 (100) | 496 (100) | 103 (100) | 33 (100) | 999 (100) |
| Identifiable C+E. L-TDI (%) | 367 (100) | 494 (99.6) | 103 (100) | 33 (100) | 997 (99.8) |

#### 2.1.1. The University of California San Francisco (UCSF) cohort

From the original 501 samples available^44^, we included a total of 367 patients with a confirmed GBM diagnosis and available overall survival (OS) and status (death=1; lost-to-follow-up=0). The inclusion criteria were the same as those previously described and are only summarized here for completeness^42^. Discarded samples included missing OS data, IDH mutation status different from wildtype, longitudinal samples, and a small number of low-grade IDH-wildtype due to potential inconsistencies. The final sample contained high-quality multimodal imaging data and tumor multi-tissue segmentations obtained from automated deep learning models, and manually corrected if necessary^44^.

#### 2.1.2. The University of Pennsylvania (UPENN) cohort

From the original 671 samples available^45^, we included a total of 496 patients with a confirmed GBM diagnosis and available OS and status (death=1; lost-to-follow-up=0). Similarly, the inclusion criteria were identical to those already described and are summarized here for completeness^42^. Discarded samples included missing OS data, IDH mutant, not otherwise specified (NOS/NEC) IDH mutation, and longitudinal samples. The final sample contained high-quality multimodal imaging data and tumor multi-tissue segmentations obtained from automated deep learning models, and manually corrected if necessary^45^.

#### 2.1.3. The Cancer Genome Atlas (TCGA) cohort

The original TCGA database includes 1122 entries with a comprehensive molecular profiling of heterogeneous adult diffuse gliomas^46^. A subset of those contained imaging data available in The Cancer Imaging Archive^47^, split into low-grade (TCGA-LGG; https://www.cancerimagingarchive.net/analysis-result/brats-tcga-lgg/), which were discarded, and high-grade (TCGA-GBM; https://www.cancerimagingarchive.net/analysis-result/brats-tcga-gbm/). From the 135 patients, we included 103 entries with confirmed IDH-wildtype (4 IDH-mutant, 28 unknown), available OS, and status (death=1; lost-to-follow-up=0). The imaging data were available through the Brain Tumor Segmentation Challenge, which required the identification of the 103 samples included by cross-referencing the subject IDs. Crucially, 75 subjects belonged to the training dataset and had automatically segmented and manually inspected multi-tissue tumor segmentations, while 28 belonged to the validation dataset without openly available segmentations. Consequently, we used the BraTS MRI segmentation bundle from the MONAI Model Zoo^48^ to perform an automated deep learning-based segmentation of the 28 multi-tissue tumor masks^49^. The final outputs were manually inspected, and 1 of them was manually corrected by a board-certified radiation oncologist. Surgical resection data were unfortunately not recoverable, but 85 had available preoperative Karnofsky performance status (KPS) scores, 65 patients had available MGMT promoter status, and a percentage of right censoring of 20.4%.

#### 2.1.4. The Río Hortega University Hospital (RHUH) cohort

From the original 40 samples available^50^, we included 36 with confirmed IDH wildtype. We then discarded 2 patients with recorded previous treatment and 1 due to incomplete survival data. We note that right-censoring was coded the opposite way, with status 0 for recorded death and 1 for lost-to-follow-up. We reversed this coding to match the other cohorts. No information on MGMT promoter status was available. The extent of resection (EOR) was originally encoded as follows: gross total resection (GTR), defined as no residual tumor enhancement, and near-total resection, defined as EOR exceeding 95% of the preoperative enhancing volume. Crucially, this definition conflicted with that of the UCSF and UPENN cohorts, in which GTR was defined as removing at least 95% of the preoperative enhancing volume. Consequently, we homogenized both by considering the 33 patients in the RHUH cohort as having undergone GTR. The final sample thus included 33 patients with preoperative imaging, available preoperative KPS score, and surgical resection information. A total of 9 patients were right-censored. The final sample contained high-quality multimodal imaging data and tumor multi-tissue segmentations obtained from automated deep learning models, and manually corrected if necessary^50,51^.

### 2.2. Survival data across institutions

A further source of heterogeneity in the final sample (999 patients with imaging and survival data) was the entry point from which OS was recorded: days from diagnosis in UCSF, days from surgery in UPENN and TCGA, and days from histopathological diagnosis in RHUH. Because histopathological diagnosis requires tissue obtained at surgery, the RHUH entry point is effectively equivalent to that of UPENN and TCGA. Consistent with this, pairwise log-rank tests detected no survival differences among these three cohorts (all p > 0.05). We therefore pooled them into a single site (OTHER), contrasted with UCSF. In previous work^42^, we attributed the survival difference between the two sites to the earlier entry point in UCSF and corrected it by rescaling the recorded OS of each patient as

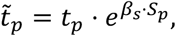

with the *s_p_* = {0,1} for the UCSF and OTHER sites, respectively. Here *β_s_* is the “site” log-hazard ratio from a Cox model with “site”, age, biological sex, EOR, and MGMT promoter status. This correction treats *e^βs^*^⋅*S*^*^p^* as a time ratio, which holds exactly only under an exponential baseline hazard. Instead, here we examined the site difference more thoroughly, in six steps that are listed here but described in Suppl. B.

i. Covariate balance and missingness between sites, quantified with standardized mean differences.
ii. An adjustment ladder, starting from site as the sole covariate and sequentially adding age, biological sex, EOR, and MGMT promoter status.
iii. The duration of follow-up per site, as median potential follow-up from reverse Kaplan–Meier curves.
iv. The completeness of follow-up at 12 and 24 months, using a completeness index and person-time rates^52,53^.
v. Whether covariates predicted censoring, using Cox models with censoring and death as competing outcomes within each site.
vi. The sensitivity of the adjusted site effect to informative censoring, through a delta-adjusted tipping-point analysis with multiple imputation.

### 2.3. Normalization to the Montreal Neuroimaging Institute template

Similarly, we followed an already established pipeline to normalize all the neuroimaging data to the common Montreal Neuroimaging Institute (MNI ICBM 2009b NLIN Asymmetric) 0.5mm^3^ isotropic space; hereafter referred to as the MNI template. For convenience, we summarize it here, but for all the nuances and details, we refer the reader to our previous work. The structural and multi-label lesion segmentations were nonlinearly normalized to the MNI template. Before normalizing, the multilabel segmentation masks were binarized and inverted to obtain a complementary binary image, ensuring that 1) pathological tissue – here defined as the whole tumor segmentation – was masked out and not used during the non-linear optimization step, and 2) the shape of the lesion is preserved after the transformation^54^. Then, we mapped the T2 images to the T2 MNI template and applied the transformation to the tumor masks using the *antsRegistrationSyNQuick* routine^55^. The normalized images were overlaid onto the MNI template to visually assess the alignment of anatomical landmarks (e.g., sulci and gyri). Quality control outputs are publicly available at https://github.com/JoanSano/NormWM_4_Glioblastoma/tree/master.

### 2.4. Computation of the tumor volumes and the lesion-tract density indices

The volume of the tumors was computed in cubic centimeters in MNI. More specifically, the number of voxels in a predefined binary mask was transformed into cubic centimeters in the following way:

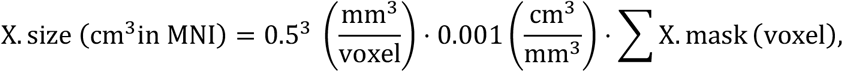

where “X.size” stands for “W.size” (size of the whole tumor), “C.size” (size of the tumor core or necrosis), “NE.size” (size of the non-enhancing tissue, edema, or T2/FLAIR hyperintensity), “E.size” (size of the contrast-enhancing tissues), or “C+E.size” (size of the joint consideration of necrosis and contrast-enhancing tissue). Identically in the case of “X.mask”.

The Lesion-Tract Density Index (L-TDI) was obtained following the steps in the original definition using a normative tractogram composed of 11.8 million streamlines modeling average white matter tracts from healthy individuals^20,42^. We selected the streamlines that intersected a given binary mask and obtained the corresponding lesion-tract density map by counting the number of streamlines passing through every voxel. The L-TDI was the tract density within the lesion-tract density map (up to an arbitrary normalization constant), thus encompassing the total white matter density of the whole brain-lesion normative circuit.

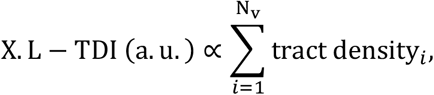

where “X.L-TDI” stands for L-TDI derived from “W.L-TDI” (the whole tumor mask), “C.L-TDI” (the necrosis or core mask), “NE.L-TDI” (the non-enhancing tissue, edema, or T2/FLAIR hyperintensity mask), “E.L-TDI” (the contrast-enhancing mask), or “C+E.L-TDI” (the union of the necrosis and contrast-enhancing masks). The number of voxels (N_v_) within a given lesion-tract density map was taken as all voxels being traversed by at least 1 streamline. The proportionality constant was taken as the number of voxels in the MNI template.

### 2.5. Statistical analysis, multiple comparisons, and the Benjamini-Bogomolov procedure

For convenience, throughout this section, the term “imaging-derived measurements” (IDMs) is used to denote the ten quantitative metrics examined in this study: W.size, C.size, NE.size, E.size, C+E.size, W.L-TDI, C.L-TDI, NE.L-TDI, E.L-TDI, and C+E.L-TDI. All the statistical procedures were performed separately for IDMs related to tumor size and IDMs related to the L-TDI.

Initially, we tested for similarities across measures by computing the Pearson correlation between pairs of IDMs. Then, to simplify initial OS associations, we discarded patients who did not experience an event (i.e., status=0) and obtained the Pearson correlation (*ρ*) between OS and imaging-derived measures in patients who were known to be deceased (i.e., status=1). However, although useful for exploratory analysis and easily interpretable, Pearson or even Spearman tests may be underpowered and do not account for censored data. Consequently, we proceeded to more nuanced statistical tests.

Patients were divided into two categories depending on whether they were dead or alive at a given time, e.g., 6 months after being diagnosed. Then, we tested for differences in IDMs in each category using non-parametric Mann-Whitney U tests. This procedure was performed for several time horizons, ranging from 6 to 48 months after initial diagnosis, every 6 months. This enabled us to independently evaluate the prognostic value of each IDM and identify the specific survival horizon at which a metric’s predictive power was lost. Afterwards, we proceeded to perform Kaplan-Meier curve analysis. For a given IDM, we obtained the underlying distribution and defined a given percentile (p) to stratify the patient population into two groups: IDM<=p and IDM>(100-p). We then obtained the corresponding survival estimators, computed the Kaplan-Meier curves, computed the median OS defined as the time at which each respective Kaplan-Meier curve reaches 0.5 (i.e., 50% survival probability), and obtained the corresponding p-value using the log-rank test. Although commonly employed in retrospective survival analyses, percentile-based stratification introduces an unavoidable element of arbitrariness. Thresholds around the 20th–30th percentiles are frequently used, effectively contrasting patients at the extremes of the biomarker distribution. Thus, to determine the extent to which statistical inference depends on this choice, we systematically varied the stratification percentile p from 10 to 50 in increments of 0.1 and examined the resulting survival associations across the full spectrum of thresholds. When applicable, the 95% confidence intervals around the observed median OS were obtained by bootstrapping with 200 resamples.

The statistical framework we designed involved numerous statistical tests using measures expected to be highly correlated. In cases where a single statistical test was performed for single measurements (e.g., Pearson correlation between L-TDIs), we applied the Benjamini-Hochberg method to control the false discovery rate (FDR) with a corrected significance level of 0.05. However, an important part of the statistical design had a hierarchical component in which associations with OS were tested in all IDMs. Therefore, we grouped the hypotheses tested in families of hypotheses, each family comprising the statistical tests performed with a given IDM (e.g., non-enhancing tumor). In such cases, using the Benjamini-Hochberg method within each family independently, or applying it to all hypotheses from all families, does not ensure an appropriate level of confidence about the filtration of errors within the selected families. Consequently, in tests that had this nested component, we applied the Benjamini-Bogomolov procedure^56^. Additionally, this procedure contains a family selection step, in which p-values within each family are combined to discard families in a statistically principled manner. In other words, it allows the experimenter to conclude which families of hypotheses contain phenomena that survive stringent FDR controls based solely on statistical outcomes rather than subjective assessments of the individual tests within such a given family. Lastly, the Benjamini-Bogomolov procedure reverts to the Benjamini-Hochberg method if all the families are selected. Hereon, “FDR” refers to the canonical Benjamini-Hochberg procedure, while “BB” refers to the Benjamini-Bogomolov procedure.

The BB procedure performs statistical corrections on two levels: family-wise and within each family. Initially, all p-values were grouped into 5 different families (i.e., whole tumor, core, non-enhancing, contrast-enhancing, core plus contrast-enhancing), and a p-value per family was obtained using the Simes combination procedure^57^. We then selected the families with an FDR corrected p-value lower than 0.05. Families that did not meet this criterion were excluded, and all the p-values within said family were set to the unit value. For families that met such a criterion, p-values were FDR corrected using a modified significance threshold,

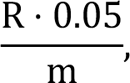

where R denotes the number of selected families and m=5 the total number of families. From the equation above, it becomes clear that if all families are selected, the BB procedure reverts to the canonical FDR procedure, and no clear statistical rationale exists for preferring one family of hypotheses over another.

### 2.6. Univariate and multivariate Cox proportional models

To provide a complementary picture beyond survival categorization and patient stratification, we fitted univariate Cox survival models using every IDM as the sole covariate. Furthermore, to assess the incremental prognostic value of an individual IDM, we fitted multivariate Cox models comprising common clinical features and a single IDM (e.g., W.size). A reference clinical-only model was also fitted to provide a baseline for comparison. In multivariate models, continuous covariates were z-scored and categorical covariates were min-max scaled. Thus, hazard ratios were expressed per one-standard-deviation change, making their magnitudes directly comparable across predictors measured in different natural units, without altering the model’s partial likelihood, overall fit, or significance tests. We fitted stratified Cox models, which estimate a separate non-parametric baseline hazard for each stratum while constraining covariate effects to be shared across strata. Within each cross-validation split, covariates were standardized using training-set statistics, which were then applied to the corresponding test set to avoid information leakage.

Hazard ratios (HRs), corresponding 95% confidence intervals (CIs), and statistical significance were obtained using two-sided Wald tests. Model discrimination was quantified using Harrell’s concordance index (C-index). In the case of univariate models, where the significance of the C-index might be doubted, the statistical significance of the C-index was assessed using one-sided permutation testing (5,000 permutations), in which the association between survival outcomes and the predictor was randomly disrupted. For both univariate and multivariate models, the 95% CIs of the C-index were computed using bootstrapping with 5000 resamples.

Models that were nested with respect to the clinical-only regression were compared using log-likelihood ratio tests with one degree of freedom (i.e., coming from the additional covariate). Models that were not nested, e.g., comparing the size of the contrast-enhancing tumor and the L-TDI derived from the whole tumor, were compared with the difference in their corresponding Akaike Information Criteria (AIC) scores. Commonly, a difference above 10 is taken as strong evidence in favor of the model with a lower AIC, while differences in AIC between 2 and 10 are taken as moderate evidence in favor of the model with a lower AIC. The fitting procedure was performed with the *lifelines* Python package, using Breslow’s and Efron’s methods to estimate the baseline hazards and to break ties, respectively.

## 3. Results

### 3.1. Multi-site survival harmonization

To increase statistical power, we analyzed all four cohorts jointly. Overall survival (OS) differed across cohorts (Fig. 1a; χ² = 14.0271, p = 0.0029; df = 3, log-rank test), driven by longer OS in the UCSF cohort. Patients were predominantly younger males (Fig. 1b–c), with little variability across cohorts (Table 1). EOR was recorded in all cohorts except TCGA, although only UCSF included biopsy-only patients. Preoperative KPS was not used in this study since it was unavailable for the entire UCSF cohort and most other patients. Methylated MGMT promoter status, associated with benefit from temozolomide^60^, was markedly more frequent in UCSF. In a complete-case Cox model (i.e., after dropping missing entries), adjustment for age, sex, EOR, and MGMT accounted for 55% of the overall “site” log-hazard ratio, mostly through MGMT (Fig. S1a). Follow-up also differed substantially (Fig. S1b; χ² = 182.8751, p < 0.0001; log-rank test). Within UCSF, censoring was predicted by older age, biological sex, and incomplete resection, the same factors that predicted death, suggesting that patients lost to follow-up were at higher risk. A tipping-point analysis showed that the residual site effect would vanish if censored UCSF patients died ∼1.6 times faster than comparable patients remaining in follow-up (see Suppl. B). Therefore, although the UCSF survival advantage was mainly attributable to the MGMT promoter, and to a lesser extent, age imbalance, a residual “site” effect could not be categorically excluded (HR = 1.1481, p = 0.1706; two-sided Wald’s z test). It was compatible with either differing entry points or informative censoring in UCSF, which these data could not distinguish. Consequently, we analyzed uncorrected OS data, but repeating all analyses with site-corrected survival times yielded virtually identical results.

**Figure 1.**
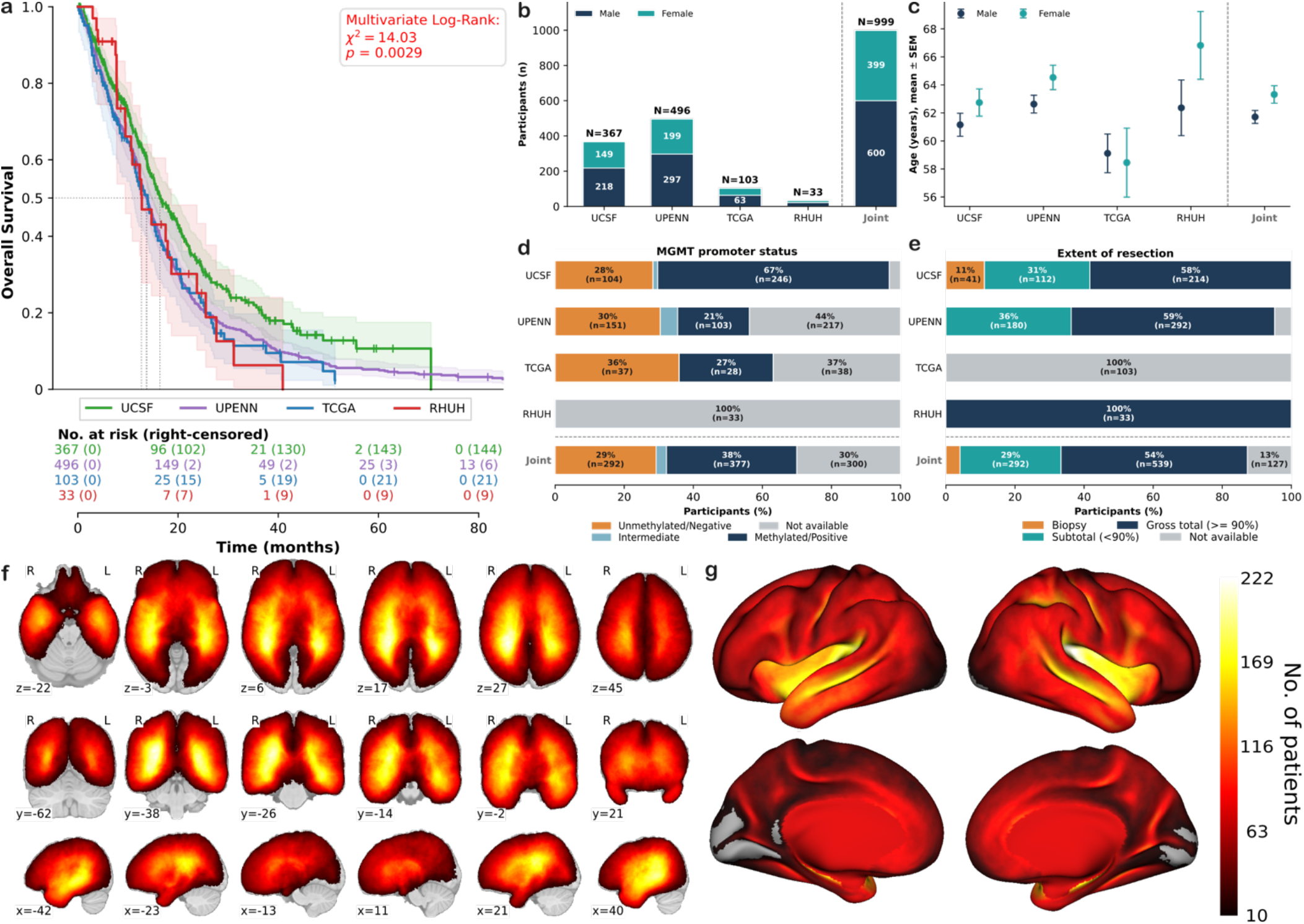
Data description. **(a)** Kaplan-Meier curves from all 4 cohorts. Significant differences in survival rates were present (χ²=14.0271, p=0.0029; df=3, log-rank test), driven mainly by the MGMT promoter imbalance in the UCSF cohort (Fig. S1; see also Suppl. B). **(b-e)** Descriptive plots for the 4 cohorts analyzed for **(b)** biological sex, **(c)** age, **(d)** MGMT promoter, and **(e)** extent of resection (see also Table 1). **(f-g)** Overlap of the whole tumor binary mask overlaid on the **(f)** volumetric and **(g)** surface MNI templates. The colormap, shared between plots, depicts the number of patients showing a GBM in each voxel or vertex.

The resulting cohort comprised up to 999 patients with newly diagnosed IDH-wildtype GBM collected across four independent institutions (UCSF, n=367; UPENN, n=496; TCGA, n=103; RHUH, n=33), with the final sample size determined by the availability of the relevant imaging-derived measurements (IDMs) and clinical data (Table 1). For the final cohort, the mean age was 62 ± 12 years, and 60.1% of patients were male. Overall, 18.8% of patients were right-censored (i.e., were alive at the last available follow-up), although censoring rates varied across cohorts. IIDMs were available for nearly all patients.

Tumors were evenly distributed throughout the brain parenchyma, except for the occipital lobe and cerebellum (Fig. 1f). The highest spatial overlaps were observed in central brain regions, consistent with the known predilection of GBM for periventricular white matter and deep structures. At the cortical level, the insular cortex exhibited the greatest degree of overlap across patients (Fig. 1g). This finding most likely reflects the anatomical position of the insula at the brain’s geometric center, such that tumors arising in adjacent frontal, temporal, and parietal territories will inevitably encroach upon it independent of any true biological tropism.

### 3.2. Survival associations depend on tumor volume compartment and stratification threshold

The volumes of the different tumor compartments were correlated with one another (Fig. 2a). As expected, W.size was most strongly associated with NE.size (*ρ*=0.89, p<0.0001, two-sided exact test FDR corrected), indicating that variation in total tumor burden was largely driven by the extent of T2/FLAIR hyperintensity and edema. The same held for the size of contrast-enhancing tissue (i.e., “E.size”) and the necrosis/core of the tumor (i.e., “C.size”), albeit to a smaller degree. The correlations between the “NE.size” and “E.size” as well as the “NE.size” and “C.size” were the lowest but still significant, while “E.size” and “C.size” were highly correlated. This likely highlights distinct growth mechanisms for cancerous cells in the non-enhancing regions compared with the processes of tumor promotion within the enhancing and necrotic regions. Overall, these findings highlight substantial interdependence among volumetric measures and underscore the difficulty of disentangling the contributions of each tissue compartment.

**Figure 2.**
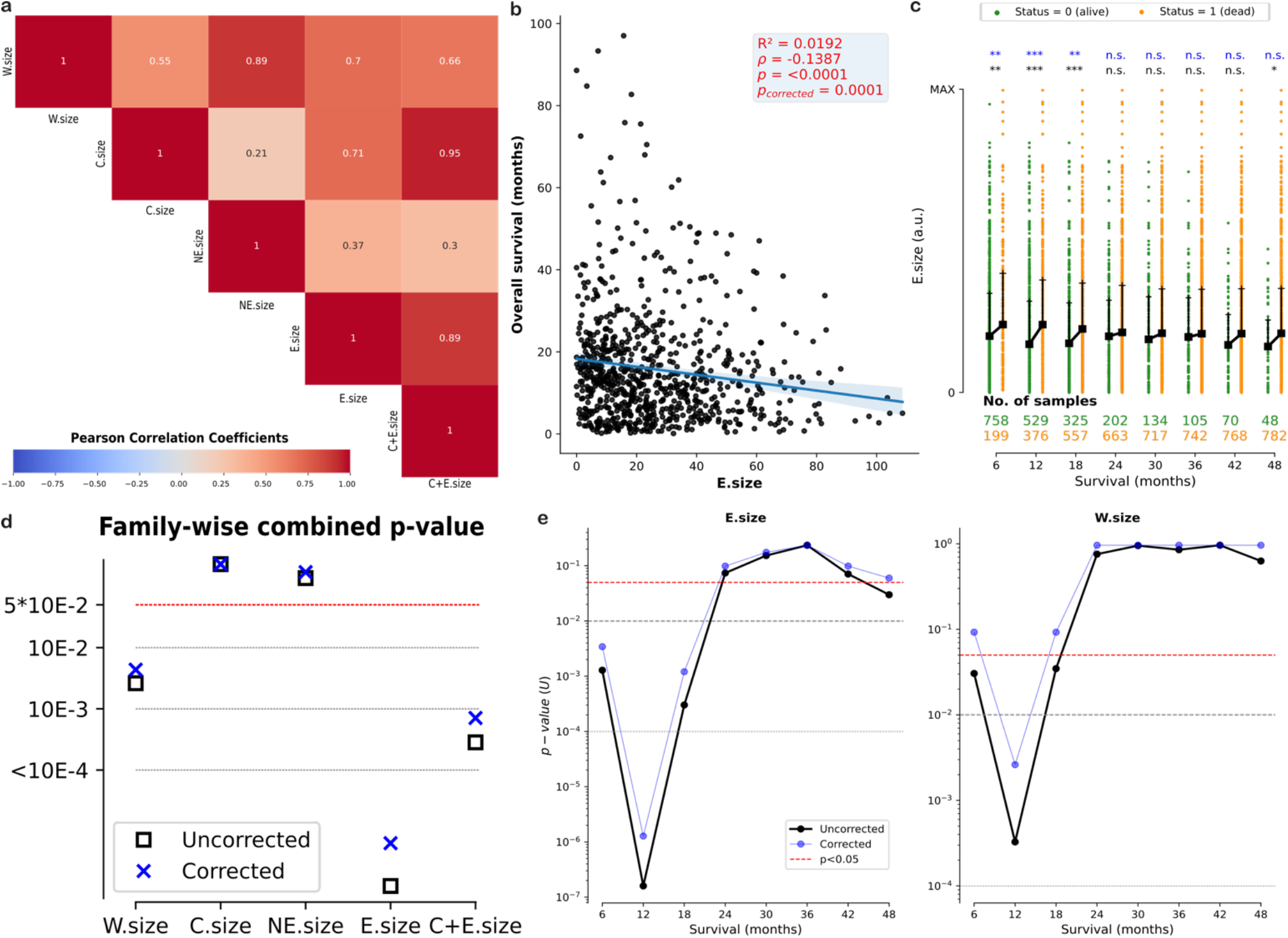
Tumor volume differences across survival times. **(a)** Pearson correlation between the sizes of the different tissues (p<0.0001, FDR corrected exact test). W.size: whole tumor size; C.size: size of the core or necrosis; NE.size: size of the non-enhancing or edema; E.size of the enhancing tissue; C+E.size: size of the combined core and enhancing tissues. **(b)** Linear correlation between the overall survival (OS) and the size (number of voxels in MNI) of the enhancing tissue (FDR corrected exact test). Only patients who experienced an event were considered (status=1). The line depicts the corresponding linear fit, and the shaded areas the 95% confidence interval. **(c)** Volume distributions of the enhancing tissue of dead (orange) and alive (green) patients across multiple survival thresholds. Black squares and lines show the median and third quartiles, respectively. In black text, the p-values of each comparison (‘***’ p<0.001, ‘**’ p<0.01, ‘*’ p<0.05, and ‘n.s.’ p>0.05, two-sided Mann-Whitney U-tests). In blue, BB corrected p-values. **(d)** Simes combined p-values (black squares), shown in log scale for clarity. Blue crosses indicate the FDR corrected family-wise p-values. Dashed horizontal lines depict different significance thresholds (red: p<0.05, gray: p<0.01, p<0.001, and p<0.0001, respectively). **(e)** Mann-Whitney U-tests, also shown in **(c)**, for different survival thresholds comparing the sizes of the enhancing tissue (left) and whole tumor (right). Each p-value tests for differences in tissue sizes in patients who died before a given threshold. Raw and BB corrected p-values are shown in black and blue, respectively.

To obtain an initial estimate of prognostic relevance, we discarded right-censored patients and computed the Pearson correlation between tissue size and OS (Fig. 2b; see also Fig. S2a). Among all volumetric measures, only the size of the contrast-enhanced tissue was significantly associated with worse prognosis (*ρ*=-0.1357, p=0.0001, two-sided exact test FDR corrected), indicating shorter survival in patients with larger enhancing lesions. The combined volume of necrotic and enhancing tissue (C+E.size) showed a weaker association (*ρ*=-0.1048, p=0.0039, two-sided exact test FDR corrected), whereas no significant relationships were observed for the remaining compartments.

We next assessed whether volumetric differences could distinguish patients surviving beyond predefined time horizons (Fig. S2b-c). Consistent with the correlation analyses, contrast-enhancing volume differed significantly between surviving and deceased patients up to 18 months after diagnosis (Fig. 2c; p<0.05, two-sided Mann-Whitney U-test, BB corrected), with significantly lower E.size for surviving patients. This effect remained significant after multiple-comparison correction across all tested survival horizons (Fig. 2d; p<0.0001, two-sided Simes combination test, FDR corrected). These differences persisted with a considerably smaller effect when considering W.size, but only for early survival thresholds (<12 months, Fig. 2e). The size of the whole tumor and core+enhancing tissue remained statistically significant, albeit with 2 orders of magnitude less than that of the contrast-enhancing tissue (Fig. 2d). Taken together, these findings suggest that the association between the overall tumor size and survival may be less robust than previously assumed. Instead, they indicate that this relationship depends on tumour compartment and patient stratification, which could explain the large variability in the literature.

Because retrospective survival analyses commonly rely on percentile-based patient stratification, we next evaluated the sensitivity of volumetric associations to the choice of threshold. Stratification based on the median W.size failed to identify significant differences in survival rates, whereas the median E.size yielded clear separation (Fig. 3a). A similar situation was observed when using the 40/60th percentiles, but when stratifying based on the 25/75th percentiles, both measurements yielded significant differences (Fig. S3a-b; p<0.05, log-rank tests), indicating that the apparent prognostic value of tumor volume may depend on the aggressiveness of the selected stratification.

**Figure 3.**
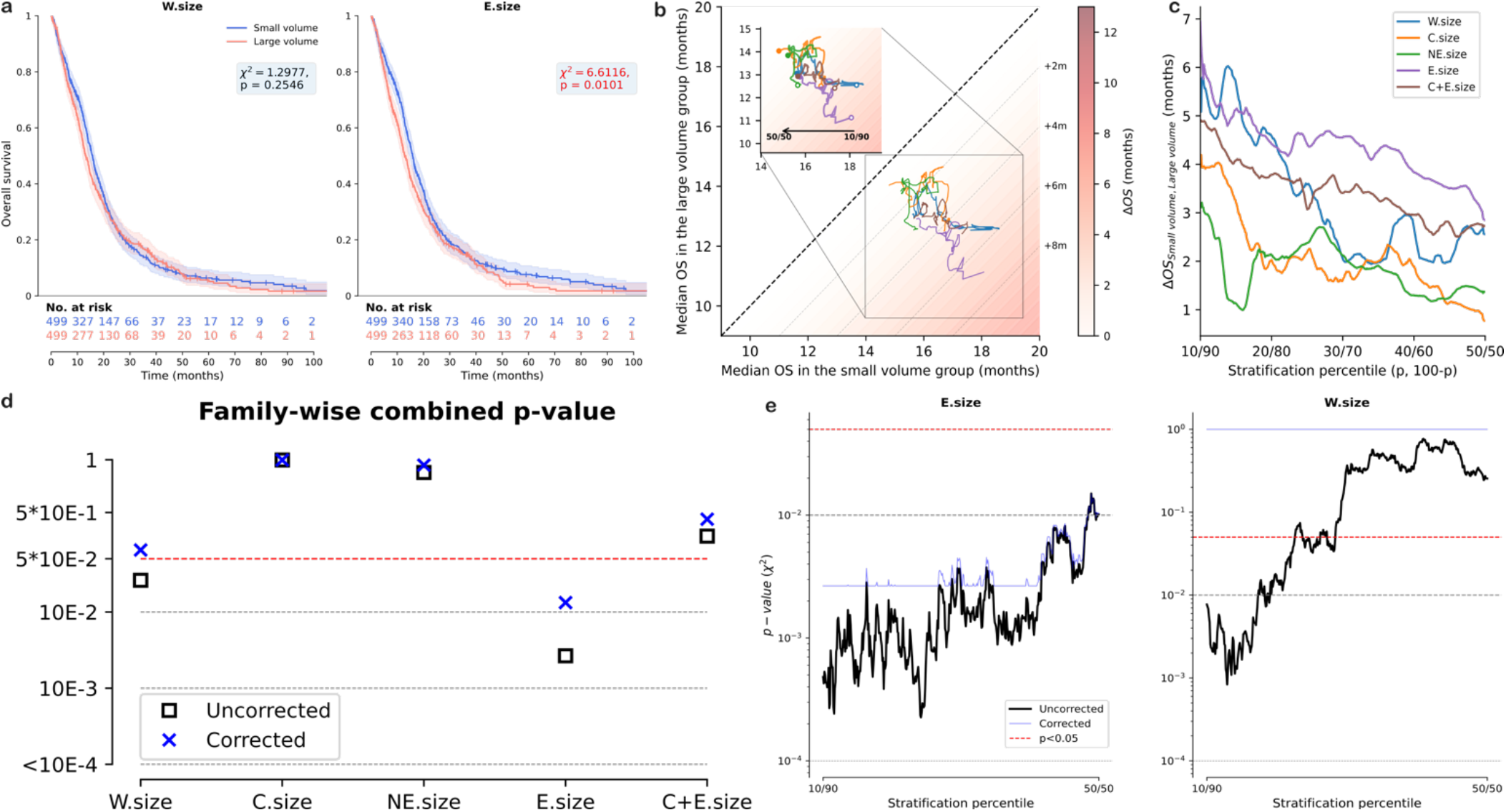
Differences in survival rates and tissue sizes across multiple stratification thresholds. **(a)** Kaplan-Meier curves for the two strata at the 50th (i.e., median size) percentile of the whole tumor (left) and enhancing tissue (right) volume. No differences in survival rates were observed when considering the size of the whole tumor (χ²=1.3154, p=0.2514; log-rank test). The group with small enhancing volumes (blue) exhibited higher survival rates than the group with large enhancing volumes (red; χ²=7.1708, p=0.0074; log-rank test). The shaded areas correspond to the 95% CI. Small vertical dashes indicate right-censored entries. **(b)** Median OS, i.e., the time at which the Kaplan–Meier curves in panel **(a)** reach 0.5, is shown as the difference between small- and large-volume groups across varying stratification thresholds. The dashed diagonal line indicates where the median OS of the two groups is identical. Light gray dashed diagonal lines represent increasing differences in median OS (e.g., +2 months). The graded red background illustrates the magnitude of these differences. As the stratification percentile becomes more extreme (filled circle: 50/50 split; white circle: 10/90 split), the curves deviate further from the diagonal. **(c)** Median OS for each group is shown as a function of percentile-based volume stratification (aggressive: 10/90; conservative: 50/50) for the different tissues identified. Shared legend with **(b),** showing different tumor tissue types. **(d)** Simes combined p-values (black squares), shown in log scale for clarity. Blue crosses indicate the FDR corrected family-wise p-values. Dashed horizontal lines depict different significance thresholds (red: p<0.05, gray: p<0.01, p<0.001, and p<0.0001, respectively). **(e)** Log-rank tests, also shown in **(a)** for the 50/50 split, comparing survival rates for a given stratification percentile using the enhancing tissue (left) and whole tumor (right). Raw and BB corrected p-values are shown in black and blue, respectively.

Median OS, defined as the time at which the Kaplan-Meier curves reach a survival probability of 50%, provides an additional measure of the separation between the small- and large-volume groups. Because these estimates depend on the percentile used for stratification, their behavior across thresholds can also inform the robustness of the observed survival difference. A progressive divergence from the diagonal indicates increasingly distinct median OS between groups, whereas an irregular pattern of divergence and convergence suggests that ΔOS is sensitive to the selected stratification threshold P. Indeed, the median OS of the small- and large-volume groups showed progressive divergence across stratification thresholds for contrast-enhancing volumes, whereas a more irregular pattern of divergence and convergence was observed for non-enhancing volumes (Fig. 3b). For the case of whole tumor sizes and intermediate picture emerged, with a rather consistent departure from the diagonal at aggressive thresholds and a more chaotic behavior at less aggressive thresholds. Directly plotting the ΔOS as a function of the stratification percentile provided a complementary picture, with differences vanishing for W.size and C.size as the percentiles approached the 50^th^ percentile (Fig. 3c).

Yet, the statistical significance of differences in median OS should be assessed via log-rank tests, which again can be computed for multiple thresholds. Crucially, among all volumetric measures, only the E.size was robustly associated with differences in survival rates, i.e., not threshold-dependent (Fig. 3d; p<0.01, two-sided Simes combination test, FDR corrected); see also Fig. S3c. This emerged due to a maintained statistical significance across aggressive and conservative thresholds (Fig. 3e left; p<0.01, log-rank tests, BB corrected). In contrast, W.size exhibited significant prognostic value but only for aggressive stratification thresholds (Fig. 3e right), whereas the sizes of both the non-enhancing and necrotic areas provided poor prognostic information.

Collectively, these results indicate that the prognostic information contained in preoperative tumor volume is largely confined to the contrast-enhancing compartment, while the apparent effect of total tumor burden is highly dependent on the chosen statistical design. Albeit in partial agreement with the literature, the overall statistical associations do cast some doubt on the effective and informative value of tumor volume as a reliable marker for patient prognosis.

### 3.3. Tractography-derived measures provide robust prognostic information across tumor compartments

Tractography-derived biomarkers are becoming more studied. Our previous work introduced the Lesion-Tract Density Index (L-TDI), a circuit-oriented description of the involvement of human GBMs within the overall white matter architecture of the human brain. Consequently, we explored whether this biomarker, shown to carry both local white matter and overall tumor morphology, presented the same *tissue-dependence* observed for the size of the tumoral mass. Upon repeating the same statistical procedures described for X.size, a rather different picture emerged.

L-TDI values derived from different tumor compartments were strongly correlated with one another (Fig. 4a; p<0.0001, two-sided exact tests, FDR corrected), indicating that distinct tissue compartments were embedded within regions of similar normative white-matter density. Correlations were lowest between L-TDIs derived from non-enhancing and contrast-enhancing tissue, suggesting some degree of compartment-specific anatomical involvement. Nevertheless, the overall correlation structure was stronger than that observed for volumetric measurements, indicating greater consistency across compartment definitions.

**Figure 4.**
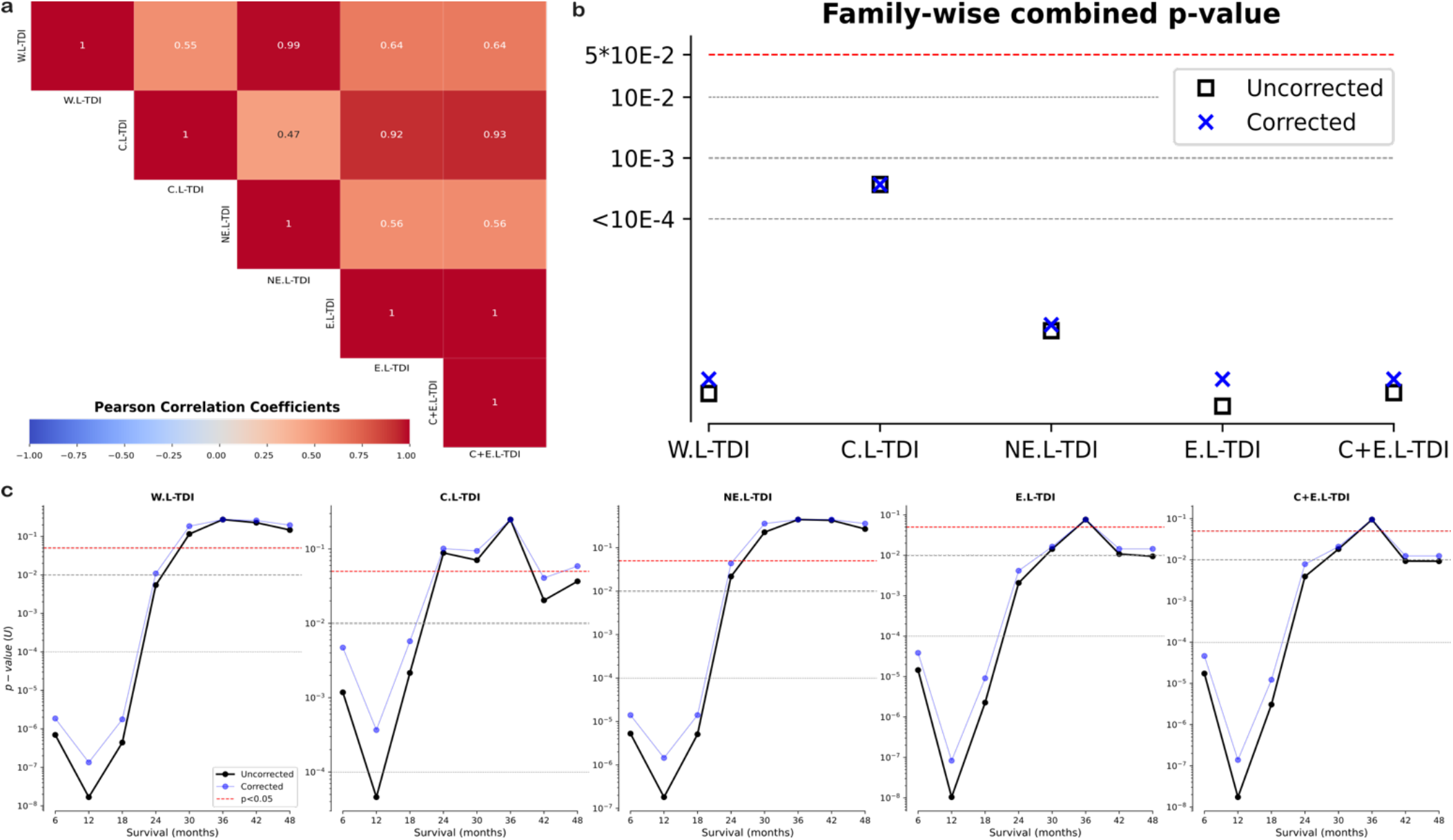
Lesion-tract density indices (L-TDI) differences across survival times. **(a)** Pearson correlation between the L-TDIs derived from the different tissues (p<0.0001, FDR corrected exact test). L-TDIs derived from whole tumor (W.L-TDI), the core or necrosis (C.L-TDI); the non-enhancing or edema (NE.L-TDI); the enhancing tissue (E.L-TDI); and the combined core and enhancing tissues (C+E.L-TDI). **(b)** Simes combined p-values (black squares), shown in log scale for clarity. Blue crosses indicate the FDR corrected family-wise p-values. Dashed horizontal lines depict different significance thresholds (red: p<0.05, gray: p<0.01, p<0.001, and p<0.0001, respectively). **(c)** Mann-Whitney U-tests for different survival thresholds comparing L-TDIs across tissues. Each p-value tests for differences in tissue L-TDIs in patients who died before a given threshold. Raw and BB corrected p-values are shown in black and blue, respectively.

Unlike tumor volume, all tractography-derived measures exhibited significant linear associations with OS in patients with observed events (Fig. S4a). The strongest correlation was observed for the E.L-TDI (*ρ*=-0.1630, p<0.0001, two-sided exact test, FDR corrected), followed closely by the combined contrast-enhancing and necrotic L-TDI (C+E.L-TDI) and the whole-tumor L-TDI (W.L-TDI). Notably, all L-TDI-derived associations were consistently stronger than those observed for the corresponding volumetric measures. Similarly, we observed robust differences in the median L-TDIs for patients who survived an arbitrary number of months since the date of diagnosis (Fig. S4b), and even if the overall robustness of such differences was weaker in the case of the C.L-TDI, all tissues resulted in robust statistical associations (Fig. 4b; p<0.0001, two-sided Simes combination test, FDR corrected). Indeed, these differences persisted beyond the previously noted limit of 18 months for the size of the contrast-enhancing tissue and remained significant for the E.L-TDI marker even at ∼3.5 years (i.e., 42 months) after diagnosis (Fig. 4c; p<0.01, two-sided Mann-Whitney U-test, BB corrected). Even in the C.L-TDI, the differences lost statistical significance from the 2-year threshold, highlighting a more stable effect to be detected through Kaplan-Meier curve analysis.

Stratification at the median L-TDI yielded significant survival differences for all tissue compartments (Fig. 5a), a result not observed for most volumetric measures. Across the full range of stratification thresholds, median OS trajectories displayed a monotonic decreasing separation between low- and high-L-TDI groups (Fig. 5b-c), depicting a continuous rather than erratic effect, which was reflected in a tissue-independent association between the L-TDI and survival rates (Fig. 5d). Such effect was stronger for the E.L-TDI, but virtually indistinguishable from that of the W.L-TDI (p<0.0001, two-sided Simes combination test, FDR corrected). Accordingly, all L-TDIs remained significantly associated with survival across threshold choices (Fig. 5e), with the strongest effects observed for the E.L-TDI (p<0.0001, log-rank tests, BB corrected) and W.L-TDI (p<0.001, log-rank tests, BB corrected).

**Figure 5.**
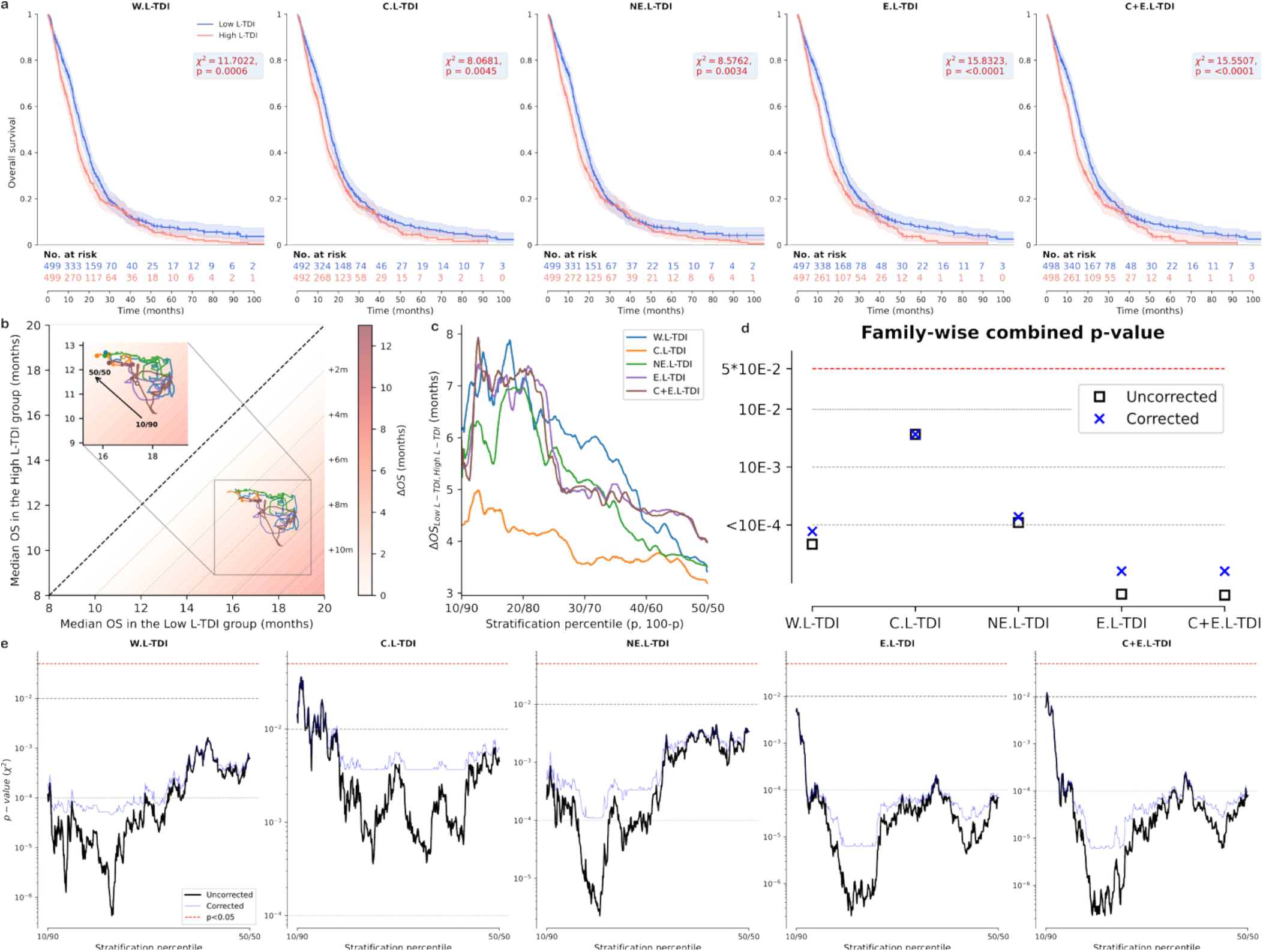
Differences in survival rates across multiple L-TDI stratification thresholds. **(a)** Kaplan-Meier curves for the two strata at the 50th (i.e., median) percentile of the L-TDI derived from the different. Differences in survival rates were observed in all cases (p<0.01; log-rank tests). The shaded areas correspond to the 95% CI. Small vertical dashes indicate right-censored entries. **(b)** Median OS, i.e., the time at which the Kaplan–Meier curves in panel **(a)** reach 0.5, is shown as the difference between low- and high-L-TDI groups across varying stratification thresholds. The dashed diagonal line indicates where the median OS of the two groups is identical. Light gray dashed diagonal lines represent increasing differences in median OS (e.g., +2 months). The graded red background illustrates the magnitude of these differences. As the stratification percentile becomes more extreme (filled circle: 50/50 split; white circle: 10/90 split), the curves deviate further from the diagonal. **(c)** Median OS for each group is shown as a function of percentile-based L-TDI stratification. Shared legend with **(b),** showing different tumor tissue types. **(d)** Simes combined p-values (black squares), shown on a log scale for clarity. Blue crosses indicate the FDR corrected family-wise p-values. Dashed horizontal lines depict different significance thresholds (red: p<0.05, gray: p<0.01, p<0.001, and p<0.0001, respectively). **(e)** Log-rank tests, also shown in **(a)** for the 50/50 split, comparing survival rates for a given stratification percentile using the L-TDIs derived from the different tissues. Raw and BB corrected p-values are shown in black and blue, respectively.

Collectively, these findings demonstrate that tractography-derived measures provide substantially more robust prognostic information than conventional volumetric assessments. Unlike tumor volume, whose prognostic value depends strongly on tissue compartment and stratification threshold, L-TDI remains predictive of survival across tumor compartments, survival horizons, and statistical designs, supporting the use of connectomic biomarkers as a more comprehensive measure of glioblastoma burden.

### 3.4. Univariate Cox models identify tractography-derived measures as the strongest imaging predictors of survival

To obtain a threshold-independent assessment of prognostic performance, we fitted univariate Cox proportional hazards models using each IDM. Hazard ratios (HRs), concordance indices (C-indices), and associated significance levels are summarized in Fig. 6a-b and Table 2. Consistent with the stratified survival analyses, the prognostic value of volumetric measurements depended strongly on the tumor compartment considered. Among all volumetric markers, contrast-enhancing volume (E.size) exhibited the strongest association with OS, yielding the largest HR and C-index within the volumetric family. Larger enhancing volumes were associated with an increased risk of death (HR=1.0069, 95% CI=1.0034–1.0105, p=0.0006, two-sided Wald’s test FDR corrected) and yielded the highest C-index among volumetric markers (C-index=0.5487, 95% CI=0.5258–0.5717, p=0.0010, one-sided permutation test). In contrast, whole-tumor volume (W.size), necrotic volume (C.size), and non-enhancing volume (NE.size) showed weaker effects and limited discriminatory performance, despite reaching statistical significance in some cases.

**Figure 6.**
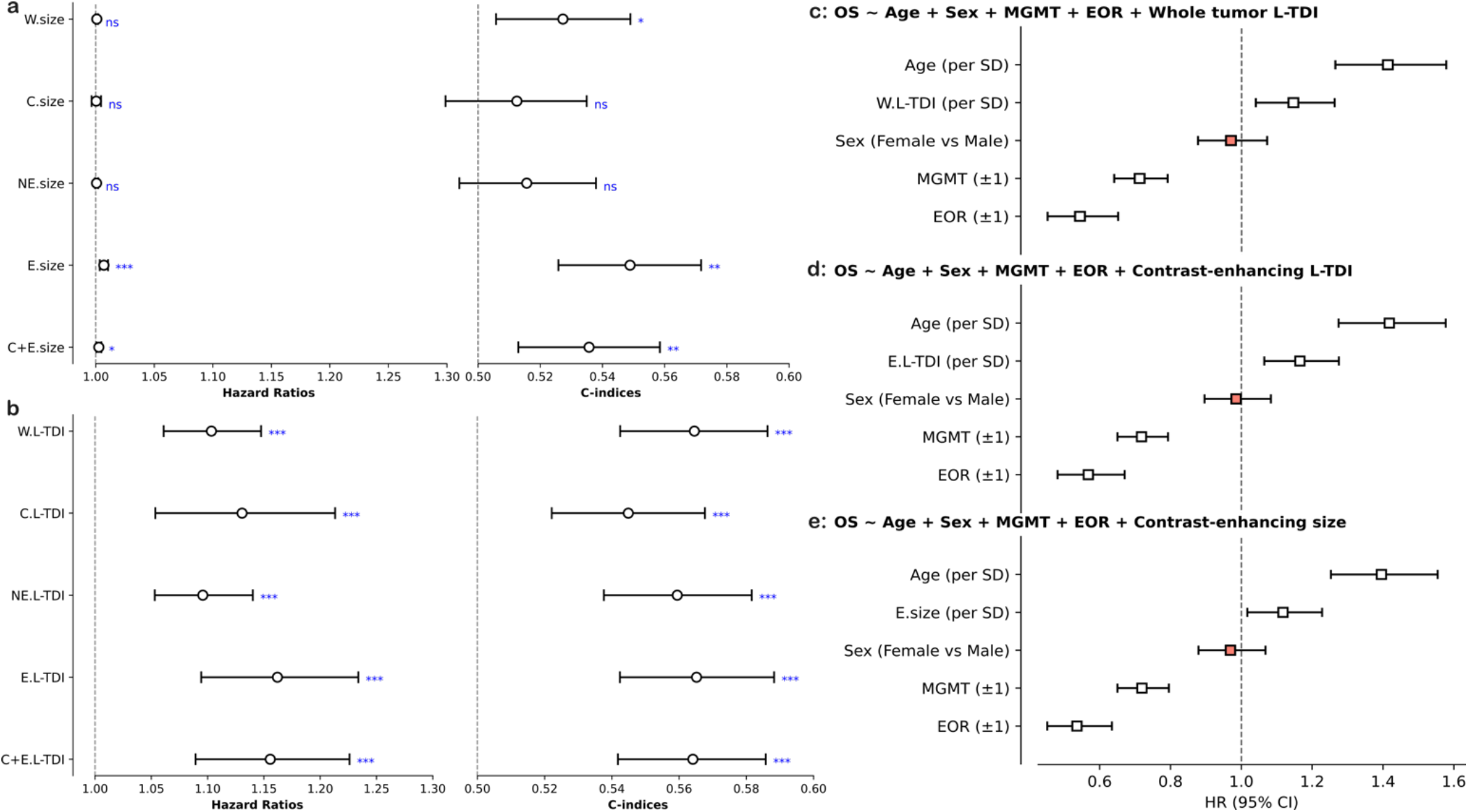
Hazard ratios and concordance indices of different tissues. **(a)** Hazard ratio (left) and concordance index (right) derived from univariate Cox proportional survival models for each tissue volume. **(b)** Identical to **(a)** but using the L-TDIs derived from each tumor tissue. ‘***’ p<0.001, ‘**’ p<0.01, ‘*’ p<0.05, and ‘n.s.’ p>0.05; two-sided Wald tests for the hazard ratios and one-sided n=5000 permutation tests for the concordance indices; FDR corrected. **(c-e)** Multivariate stratified Cox survival models with standardized features available post-surgically. Hazard ratios corresponding to models incorporating **(c)** the L-TDI derived from the whole tumor lesion mask, **(d)** the L-TDI derived from the contrast-enhancing tumor mask, and **(e)** the size of the contrast-enhancing tumor. Black error bars depict hazard ratios with statistical significance (two-sided Wald’s z test), while red markers depict otherwise.

**Table 2.**
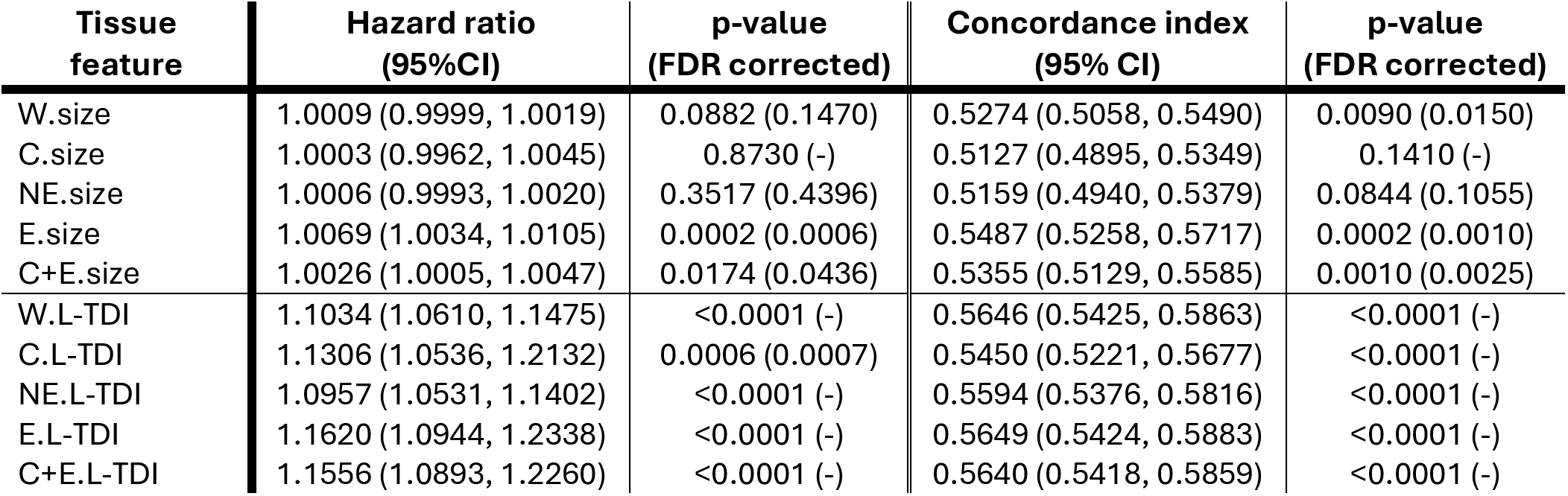
Univariate hazard ratios and concordance indices. Values within parentheses show the 95% confidence intervals for the hazard ratios and C-indices, while they show the false discovery rate corrected (FDR) p-values. “-“ depicts an unchanged p-value after the application of the FDR correction (to the decimal precision). See Table S1 for the results using Uno’s C-index.

| Tissue feature | Hazard ratio (95%CI) | p-value (FDR corrected) | Concordance index (95% CI) | p-value (FDR corrected) |
| --- | --- | --- | --- | --- |
| W.size | 1.0009 (0.9999, 1.0019) | 0.0882 (0.1470) | 0.5274 (0.5058, 0.5490) | 0.0090 (0.0150) |
| C.size | 1.0003 (0.9962, 1.0045) | 0.8730 (-) | 0.5127 (0.4895, 0.5349) | 0.1410 (-) |
| NE.size | 1.0006 (0.9993, 1.0020) | 0.3517 (0.4396) | 0.5159 (0.4940, 0.5379) | 0.0844 (0.1055) |
| E.size | 1.0069 (1.0034, 1.0105) | 0.0002 (0.0006) | 0.5487 (0.5258, 0.5717) | 0.0002 (0.0010) |
| C+E.size | 1.0026 (1.0005, 1.0047) | 0.0174 (0.0436) | 0.5355 (0.5129, 0.5585) | 0.0010 (0.0025) |
| W.L-TDI | 1.1034 (1.0610, 1.1475) | <0.0001 (-) | 0.5646 (0.5425, 0.5863) | <0.0001 (-) |
| C.L-TDI | 1.1306 (1.0536, 1.2132) | 0.0006 (0.0007) | 0.5450 (0.5221, 0.5677) | <0.0001 (-) |
| NE.L-TDI | 1.0957 (1.0531, 1.1402) | <0.0001 (-) | 0.5594 (0.5376, 0.5816) | <0.0001 (-) |
| E.L-TDI | 1.1620 (1.0944, 1.2338) | <0.0001 (-) | 0.5649 (0.5424, 0.5883) | <0.0001 (-) |
| C+E.L-TDI | 1.1556 (1.0893, 1.2260) | <0.0001 (-) | 0.5640 (0.5418, 0.5859) | <0.0001 (-) |

Non-local descriptions, derived from tractography-based descriptors of tumor burden, appeared robust to the choice of the tumor compartment, achieving higher C-indices than any volumetric measurement. The highest prognostic information was observed for E.L-TDI (C-index=0.5649, 95% CI=0.5424–0.5883), W.L-TDI (C-index=0.5646, 95% CI=0.5425–0.5863), and C+E.L-TDI (C-index=0.5640, 95% CI=0.5418–0.5858), indicating remarkably similar predictive performance across tissue compartments. In light of these, threshold-independent analyses corroborate the findings from the stratified survival analyses and demonstrate that tractography-derived measures capture more robust prognostic information than conventional volumetric assessments across tumor compartments.

### 3.5. Lesion-tract density indices retain independent prognostic value with clinical variables

To determine whether IDMs provided prognostic information beyond established clinical factors, we fitted multivariable Cox proportional hazards models incorporating age, sex, MGMT promoter methylation status, and extent of resection (EOR). Based on the preceding analyses, we selected three representative imaging markers: contrast-enhancing volume (E.size), whole-tumor lesion–tract density index (W.L-TDI), and contrast-enhancing lesion–tract density index (E.L-TDI). To make the different models directly comparable, we used the 995 subjects that had simultaneous measurements of E.size, W.L-TDI, and E.L-TDI (see Table 1). Considering post-surgical and genetic markers, a total of 615 patients had complete data. The following results were obtained with cohort-stratified Cox models after standardizing the features (see Methods), although the conclusions remained unchanged regardless of this choice (see Suppl. C1-C3).

In a pre-surgical context (Fig. S6), with age and sex as the sole clinical covariates, the W.L-TDI model yielded a C-index of 0.6305 (95% CI: 0.6098–0.6515) and an AIC of 7928.42, with W.L-TDI itself carrying a standardized hazard ratio of 1.1793 (95% CI: 1.0978–1.2669; p<0.0001, two-sided Wald’s z test). The E.L-TDI model achieved the lowest AIC among the three markers (7922.21) and a comparable C-index of 0.6298 (95% CI: 0.6085–0.6512), with a standardized hazard ratio of 1.2076 (95% CI: 1.1257–1.2954; p<0.0001, two-sided Wald’s z test). The E.size model reached a C-index of 0.6224 (95% CI: 0.6007–0.6441; AIC=7940.73), with a standardized hazard ratio of 1.1124 (95% CI: 1.0347–1.1960; p=0.0039, two-sided Wald’s z test). In all three models, age was the strongest covariate (HR 1.3683–1.3909, all p<0.0001), while sex did not reach significance (HR 0.9632–0.9648, all p>0.30). Each imaging feature conferred additional prognostic value when compared to the baseline model (W.L-TDI: ΔAIC=−18.39, p<0.0001; E.L-TDI: ΔAIC=−24.60, p<0.0001; E.size: ΔAIC=−6.08, p=0.0045; df=1, two-sided log-likelihood ratio tests). Pairwise, E.L-TDI’s AIC advantage was 6.21 points over W.L-TDI and 18.52 points over E.size.

We next assessed performance in a post-surgical setting by additionally incorporating MGMT promoter status and EOR (Fig. 6c-e). All three imaging markers retained independent associations with survival after adjustment for clinical variables. Particularly, the W.L-TDI model reached a C-index of 0.6786 (95% CI: 0.6516–0.7046) and an AIC of 4383.39, with W.L-TDI retaining a standardized hazard ratio of 1.1391 (95% CI: 1.0384–1.2497; p=0.0058, two-sided Wald’s z-test). The E.L-TDI model again achieved the lowest AIC (4380.20) with a comparable C-index of 0.6766 (95% CI: 0.6493–0.7024), and a standardized hazard ratio of 1.1653 (95% CI: 1.0648–1.2753; p=0.0009, two-sided Wald’s z-test). The E.size model reached a C-index of 0.6740 (95% CI: 0.6467–0.7004; AIC=4385.81), with a standardized hazard ratio of 1.1156 (95% CI: 1.0165–1.2243; p=0.0212, two-sided Wald’s z-test). In all full models, age (HR 1.3892–1.4177, all p<0.0001) and EOR (HR 0.5431–0.5680, all p<0.0001) were the strongest prognostic factors, MGMT methylation retaned independent information (HR 0.7181–0.7266, all p<0.0001), and sex did not reach significance in any model (HR 0.9695–0.9853, all p>0.50). Each imaging feature again conferred additional prognostic value when compared to baseline models including only MGMT, EOR, age, and sex (W.L-TDI: p=0.0059; E.L-TDI: p=0.0010; E.size: p=0.01232; df=1, two-sided log-likelihood ratio tests). Compared with the clinical baseline model (C-index=0.6719; AIC=5009.91), the addition of the E.L-TDI, W.L-TDI, and E.size improved model fit by 8.76, 5.57, and 3.15, respectively.

Taken together, these results demonstrate that tractography-derived measures provide independent prognostic information beyond established clinical predictors and outperform conventional volumetric measurements. Although information-based metrics slightly favor the L-TDI derived from the contrast-enhancing tissue, C-indices are virtually indistinguishable, and the statistical effect overlaps with the L-TDI derived from the whole tumor mask. This is further illustrated in the last two sets of analyses, which point to a more independent effect present in the L-TDI derived from the whole tumor lesion.

Using leave-one-out validation across independent testing cohorts, W.L-TDI consistently yielded higher C-indices (mean Cox C-index = 0.637 ± 0.0016 SD) than the baseline model (0.621 ± 0.014 SD), while exhibiting lower between-cohort variability than contrast-enhancing volume (Table S2), suggesting that its prognostic value is robust and transportable across independent institutions. Similar results held for models with E.L-TDI. Additionally, a bivariate Cox model combining W.L-TDI and E.size, yielded a significant HR for the former (1.1811 [1.0772, 1.2950], p=0.0004, two-sided Wald’s z test) but not for the latter (1.0338 [0.9397, 1.1374], p=0.4949, two-sided Wald’s z test). By contrast, a joint model of E.L-TDI and E.size retained statistical significance for both variables, despite their relatively uninformative hazard ratios (E.L-TDI: 1.1988 [1.1160, 1.2527], p<0.0001; E.size: 1.1374 [1.0572, 1.2238], p=0.0006).

## 4. Discussion

For decades, tumor burden in GBM has been approximated through anatomical measurements of lesion size^4,5^. Our findings challenge this assumption. Across a large, molecularly homogeneous cohort of 999 patients with newly diagnosed GBM IDH-wildtype, the prognostic information contained in tumor volume proved highly dependent on tissue compartment and statistical design. In contrast, tractography-derived measures of tumor–brain interaction remained consistently associated with survival across analytical frameworks. These observations confirm that the clinically relevant burden imposed by GBM may be determined less by the amount of tissue occupied by the lesion than by the extent to which the tumor engages the brain’s structural architecture.

A first important finding concerns the long-debated prognostic value of tumor volume. Despite more than three decades of investigation, the literature remains divided on whether larger tumors are associated with poorer survival and on which tumor compartment carries the most relevant prognostic information^7^. Our results provide a potential explanation for this inconsistency. While volumetric measures were often statistically associated with survival under specific analytical conditions, these associations were neither uniform across compartments nor stable across stratification thresholds. Notably, the contrast-enhancing compartment consistently emerged as the strongest volumetric predictor of outcome, exhibiting reproducible associations across correlation analyses, survival stratification, and Cox proportional hazards models. Whole-tumor volume, by comparison, showed substantially weaker and more threshold-dependent effects, while necrotic and non-enhancing compartments provided limited prognostic information.

These findings indicate that the heterogeneity of previous volumetric studies may not simply reflect differences in cohort composition or sample size but may also arise from a more fundamental limitation of tumor volume as a biomarker. Volume quantifies anatomical extent but provides little information regarding the biological significance of the tissue being measured or the neural systems affected by the lesion. Two tumors of comparable size may therefore impose markedly different burdens on the surrounding brain. From this perspective, the variable prognostic performance of whole-tumor volume becomes unsurprising and highlights the need for biomarkers that extend beyond purely anatomical descriptions of disease.

The consistent prognostic value of contrast-enhancing tissue is nevertheless biologically informative. Contrast enhancement reflects blood–brain barrier disruption, angiogenesis, increased cellular density, and active tumor proliferation, all hallmarks of aggressive glioblastoma biology^61^. The fact that this compartment outperformed all other volumetric measures suggests that the prognostic information contained in tumor volume is concentrated within regions of active disease rather than uniformly distributed throughout the lesion. Importantly, however, even the strongest volumetric associations remained modest in magnitude. In a cohort of this size, statistical significance should not be conflated with clinical relevance. The relatively low hazard ratios and C-indices observed for volumetric measures indicate that anatomical burden alone provides only a limited description of patient outcome.

In contrast, the lesion–tract density index (L-TDI) remained significantly associated with overall survival across tissue compartments, survival horizons, stratification thresholds, and multivariable models. Moreover, these associations consistently exceeded those observed for volumetric measurements and remained detectable after adjustment for established clinical variables. These findings indicate that tractography captures prognostic information that is not fully explained by conventional clinical or anatomical descriptors.

It is worth noting that, although the observed C-indices were statistically significant, their absolute values were modest, indicating only limited discriminative performance. This finding should be interpreted in the context of GBM prognosis, which is driven by a highly complex interplay of biological, molecular, clinical, and treatment-related factors that cannot be fully captured by a single preoperative imaging biomarker. Accordingly, the C-indices observed here are comparable to those reported for imaging-derived prognostic models with medium and large sample sizes^6,32,33^. Furthermore, after accounting for demographic, surgical, and epigenetic markers, C-indices increased without disregarding the independent effect of imaging markers. Nonetheless, we acknowledge that Harrell’s C-index may exhibit optimistic bias in the presence of right censoring, but the proportion of censored observations in our cohort was relatively low, making any potential bias unlikely to affect the conclusions (Table S1).

Within this context, the superior performance of L-TDI is particularly noteworthy, as it suggests that incorporating the brain’s structural connectivity captures prognostically relevant information not fully captured by conventional volumetric descriptors, consistent with a growing conceptual shift in neuro-oncology^15^. Historically, GBM has been viewed primarily as a localized lesion whose clinical impact is determined by its size, anatomical location, and histopathological features. Recent pre-clinical evidence has challenged this view by demonstrating extensive bidirectional interactions between glioma cells and neural circuits^62,63^. Neuronal activity can promote glioma proliferation, while tumors actively remodel the surrounding neural environment, creating a dynamic tumor–brain system that extends well beyond the visible lesion, contributing to disease progression^65,66^. Within this framework, prognosis may depend not only on local tumor aggressiveness but also on the extent to which tumors involve the brain’s structural scaffold^13,43^. Although the L-TDI and neuroimaging in general cannot directly assess neuron–glioma communication^14^, its robust prognostic association is consistent with the possibility that network-level involvement represents a clinically meaningful dimension of GBM burden that is not fully captured by conventional lesion-centric descriptions.

Considering these, our results further suggest a distinction between anatomical and connectomic burden. Anatomical burden describes the physical extent of disease, whereas connectomic burden describes the degree to which disease is integrated within large-scale brain networks. While these dimensions are related, they are certainly not equivalent^67^. Current research frames this as a Russian doll picture: anatomical burden is nested within connectomic burden, meaning that the visible lesion represents only a local expression of a broader network-level involvement, and lesion-centric descriptions therefore provide an inherently limited account of GBM’s true impact^42,43^. Volumetric and tractographic measures are correlated, i.e., larger tumors tend to engage more white-matter pathways (Table S3), yet the prognostic performance of L-TDI substantially exceeded that of tumor volume, suggesting greater disease specificity. This gap is further illustrated by a recent statistical parameter mapping study that confined volumetric survival effects to tumors with equivalent connectomic burden^68^. In tumors engaging comparable brain circuits, volume adds meaningful prognostic information, but network engagement itself remains the primary determinant. Tumor volume alone cannot capture this, requiring at least measures of local normative white matter density^40^. The underlying reason is the brain’s specialized functional and structural architecture. Evidently, GBMs engage distinct subsystems depending on location^27^, and their volume is blind to that variation. Instead, widespread tract involvement captures both dimensions simultaneously without sensitivity to the choice of tumor compartment. Although the L-TDI derived from contrast-enhancing tissue produced the most parsimonious multivariable models, whole-tumor L-TDI achieved nearly identical discriminatory performance with slightly more stable hazard ratios across cohorts. This suggests that distinct imaging phenotypes ultimately engage overlapping white-matter systems and that clinically relevant connectomic burden is distributed across tumor compartments rather than confined to a single imaging phenotype. Connectomic burden, therefore, represents a more comprehensive and biologically meaningful descriptor of disease than anatomical burden alone.

Beyond the biological implications, the present study also highlights an important methodological issue for large-scale neuro-oncology research. Publicly available datasets increasingly enable the assembly of cohorts approaching the scale required for robust prognostic analyses. However, differences in survival definitions, follow-up procedures, and clinical annotations can introduce substantial heterogeneity. While prospective studies mitigate these issues through pre-specified protocols, retrospective studies must address them analytically or, at minimum, disclose them^69^. Otherwise, predictive models risk introducing systematic biases that inflate performance and yield potentially inaccurate survival estimates. That said, GBM treatment is relatively standardized following established chemoradiotherapy guidelines^70^ and comparable surgical strategies^71^, with relatively short and homogeneous times between diagnosis and surgery. Thus, in the absence of individualized treatment times, harmonization might still allow for estimating variance attributable to different entry points rather than biological differences, follow-up procedures, or treatment-related heterogeneities. Crucially, immortal time bias cannot be fully eliminated without recorded diagnosis-to-surgery intervals, even if these are expected to be short. Results should therefore be replicated within individual cohorts, or across groups of directly comparable cohorts. Upon repeating the BB procedure for the UCSF and OTHER cohorts independently (Figs. S7-S8), the L-TDI framework exhibited stronger associations than volumetric measures, with the L-TDI derived from the whole-tumor compartment emerging as a more stable metric.

Several limitations should also be acknowledged. First, the study was retrospective and therefore subject to the inherent biases of observational datasets. Second, tractography analyses were based on a normative structural connectome rather than patient-specific diffusion MRI, which may retain residual streamline density biases related to bundle geometry and tracking algorithms and potentially underestimate interindividual variability in white-matter architecture. Although some studies found no significant differences between normative and patient-specific estimates^72^, this remains an important consideration, as connectomic measures may be sensitive to this choice^73^. However, tractography in the presence of brain tumors remains a technically challenging and unresolved problem^74^, since it may simultaneously involve disruption, infiltration, and displacement of white matter fibers related to mass effects^24,75^. Third, although the study incorporated four independent cohorts encompassing almost a thousand patients, continued replication in additional datasets will be important to establish the generalizability of the findings.

Taken together, these findings support a shift from local volumetric assessments toward network-based measures of tumor burden in glioblastoma. While anatomical volume remains a useful descriptor of disease extent, it provides an incomplete representation of how tumors affect the brain. In contrast, connectomic biomarkers capture the degree to which tumors are embedded within the structural architecture of the nervous system and may therefore offer a more clinically informative and biologically meaningful characterization of disease burden. More broadly, our results suggest that prognostic modeling in glioblastoma may benefit from moving beyond measurements of lesion size toward quantitative descriptions of tumor–brain interactions, an approach that aligns with the emerging view of gliomas as diseases of distributed neural systems rather than isolated anatomical masses.

## 5. Conclusions

We provide a statistical framework that accounts for the broad heterogeneity of volumetric outcomes in glioblastoma, demonstrating that contrast-enhancing tissue volume is the only morphological measure to withstand stringent scrutiny. Moving beyond local geometric descriptors, we show that connectome-informed markers derived from normative tractography confer systematic advantages over conventional volumetry, both for risk stratification and survival prognostication. Across the tissue compartments examined, normative tractography-based features exhibited largely comparable effect sizes, with a modest preference for those anchored to the contrast-enhancing region. Crucially, however, the prognostic benefit of the L-TDI was not contingent on any single tissue definition, suggesting that connectomic indices capture a robust signal that transcends the boundaries of individual tumor compartments. Finally, the extent to which GBM engages the brain’s structural architecture appears more informative for prognosis than the physical volume of the lesion itself. Taken together, these findings support a shift from local volumetric assessments toward brain-wide connectomic measures of tumor burden.

## Code availability

The code to perform the normalization to MNI, computation of the tract density indices, and the statistical analyses are publicly available in Open Science Framework (https://doi.org/10.17605/OSF.IO/P8XGH) and/or can be requested from the corresponding authors. All the figures in the main and supplementary text were generated with custom Python code, saved as scalable vector graphics (SVG) files, and assembled in Inkscape.

## Author contributions

*Conceptualization*: J.F.-R. and A.C. *Methodology*: J.F.-R. and A.C. *Software*: J.F.-R. and A.I. *Validation*: J.F.-R., A.I., A.J., S. L., L.S.P., J.K.A. and A.C. *Formal analysis*: J.F.-R., A.I., and A.C. *Investigation*: J.F.-R. and A.C. *Resources*: J.F.-R., J.K.A., and A.C. Data Curation: J.F.-R., A.I., A.J., and A.C. *Writing – Original Draft*: J.F.-R. and A.C. *Writing – Review & Editing*: J.F.-R., A.I., A.J., S.L., A.F., M.M., F.P., L.S.P, L.C., L.V., S.S., P.A., J.K.A., and A.C. *Visualization*: J.F.-R. and A.C. *Supervision*: A.C. *Project administration*: J.F.-R., J.K.A., and A.C. *Funding acquisition*: J.F.-R., J.K.A., and A.C.

## Data Availability

The UCSF cohort was retrieved from The University of California San Francisco Preoperative Diffuse Glioma MRI repository in The Cancer Imaging Archive (https://doi.org/10.7937/tcia.bdgf-8v37). The UPENN cohort was retrieved from the multi-parametric magnetic resonance imaging (mpMRI) scans for de novo Glioblastoma (GBM) patients from the University of Pennsylvania Health System collection in The Cancer Imaging Archive (https://doi.org/10.7937/TCIA.709X-DN49). The TCIA cohort was retrieved from the Segmentation Labels for the Pre-operative Scans of the TCGA-GBM collection in The Cancer Imaging Archive (<u>10.7937/K9/TCIA.2017.KLXWJJ1Q</u>). The RHUH cohort was retrieved from the Río Hortega University Hospital Glioblastoma dataset: a comprehensive collection of preoperative, early postoperative and recurrence MRI scans in The Cancer Imaging Archive (https://doi.org/10.7937/4545-c905). Lastly, the normative tractogram is also freely available (https://doi.org/10.6084/m9.figshare.c.6844890).

## Acknowledgements

This research was funded in part by the National Science Centre, Poland No UMO-2024/53/N/NZ4/03513 (J.F.-R., A.C.). This project has received funding from the European Union’s Horizon 2020 research and innovation programme under grant agreement No 857533 and from the International Research Agendas Programme of the Foundation for Polish Science No MAB PLUS/2019/13 (J.F.-R., A.J., J.K.A.). The publication was created within the project of the Minister of Science and Higher Education “Support for the activity of Centers of Excellence established in Poland under Horizon 2020” on the basis of the contract number MEiN/2023/DIR/3796 (J.F.-R., A.J., J.K.A.). We gratefully acknowledge Polish high-performance computing infrastructure PLGrid (HPC Center: ACK Cyfronet AGH) for providing computer facilities and support within computational grant no. PLG/2026/019442 (J.F.-R., A.J., J.K.A.). L.C. was supported by Fondazione Caritro (grant ID: CARITRO 2024.0302).

## Ethics statement

Cohort UCSF: Data collection was performed in accordance with relevant guidelines and regulations and was approved by the UCSF institutional review board with a waiver for consent. Cohort UPENN: Collection, analysis, and release of the UPenn-GBM data has happened in compliance with all relevant ethical regulations. The protocol was approved by the Institutional Review Board at the UPHS, and informed consent was obtained from all participants. Cohort RHUH: The study was conducted following the principles of the Declaration of Helsinki. Written consent was obtained from all patients, and approvals were granted by the Institutional Review Board of Río Hortega University Hospital and the Ethics Committee for Drug Research (CEIm) of the West Valladolid Health Area (Ref. 22PI-208).

## SUPPLEMENTARY MATERIAL A

**Figure S1.**
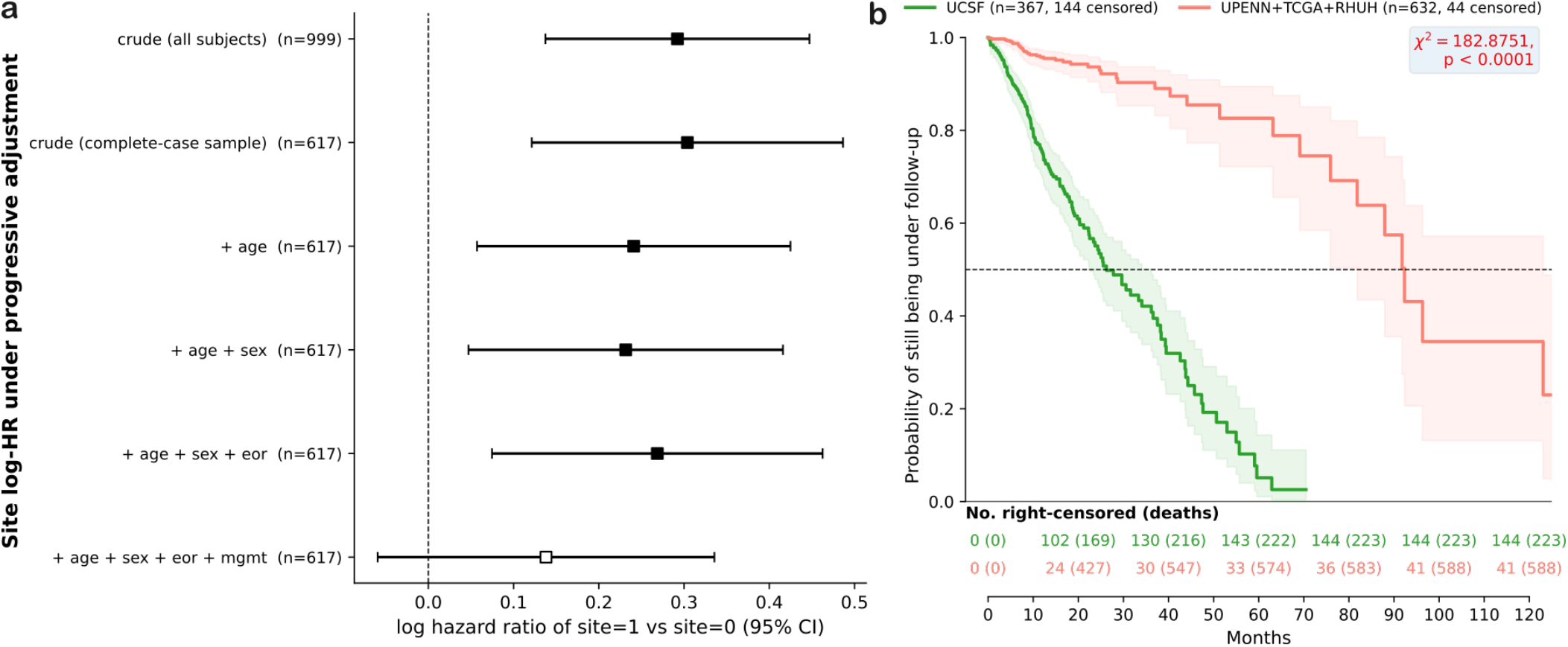
Site differences in covariate distribution and follow-up. **(a)** Log-hazard ratio of the “site” covariate along the adjustment ladder. The “crude” effect is the overall HR difference attributable to the site (i.e., a univariate Cox model with “site” as the unique covariate). As more covariates are introduced, site-related differences in survival rates persisted until MGMT promoter status was considered, which accounted for ∼55% of the variance in survival rates. Even with the full set of clinical covariates, a considerable, albeit non-significant, effect attributable to the “site” was still visible (log HR=0.1381, p=0.1706; two-sided Wald’s z test). **(b)** Reverse Kaplan-Meier curves for the UCSF and OTHER cohorts. The right-censoring rates differed significantly (χ²=182.8751, p<0.0001; multivariate two-sided log-rank test), with UCSF showing a shorter median potential follow-up than the OTHER cohorts. The shaded areas correspond to the 95% confidence interval. Although these differences in follow-up and/or right censoring could explain the observed differences in survival rates, the effect of different entry points (i.e., OS recorded from diagnosis or surgery) could not be ruled out with 100% certainty. This figure should be read jointly with Suppl. B for a more comprehensive overview.

**Figure S2.**
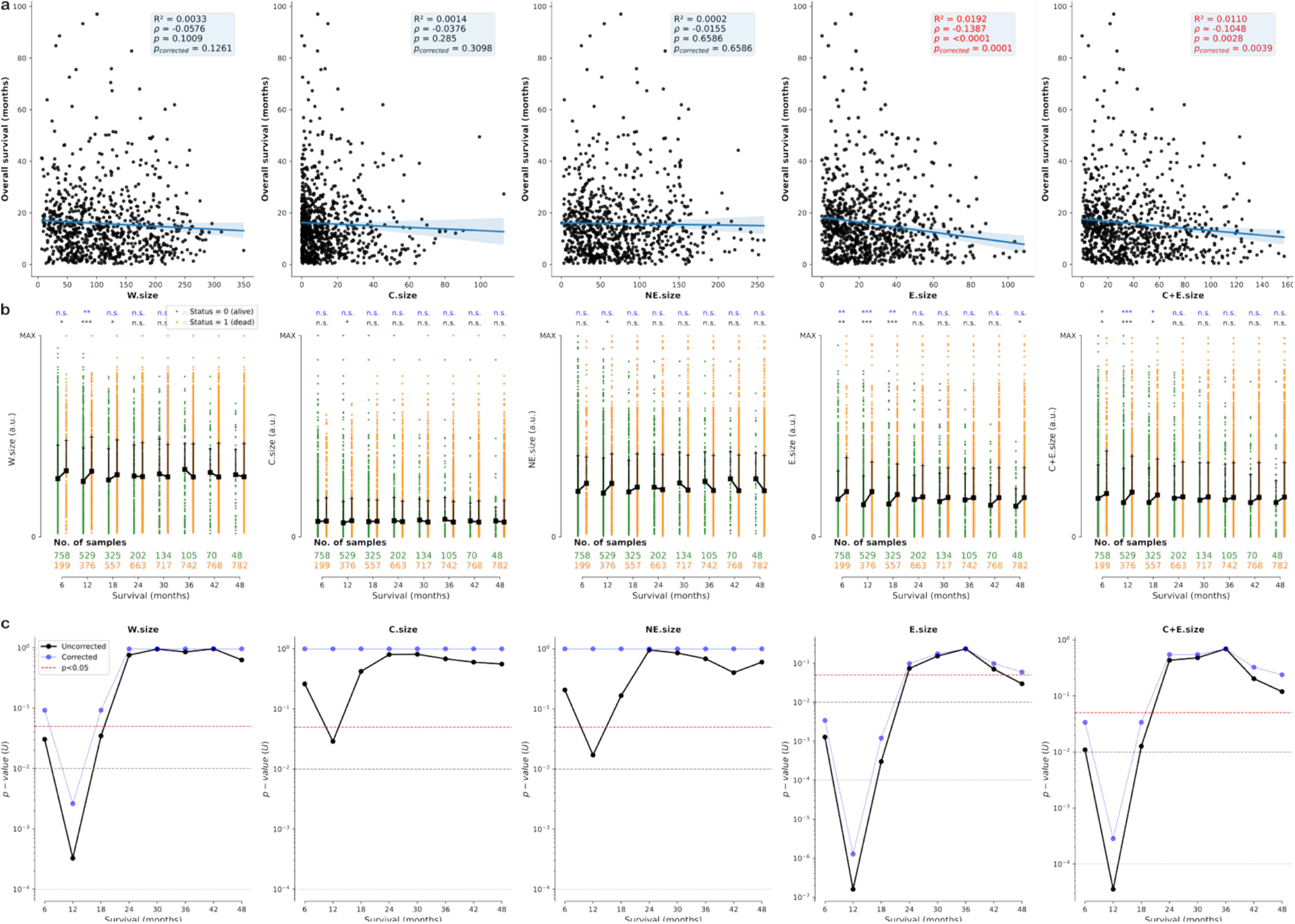
Associations between survival and tumor tissue size. W.size: whole tumor size; C.size: size of the core or necrosis; NE.size: size of the non-enhancing or edema; E.size of the enhancing tissue; C+E.size: size of the combined core and enhancing tissues. **(a)** Linear correlation between the overall survival (OS) and the size (number of voxels in MNI) of the different tissues (FDR corrected exact test). Only patients who experienced an event were considered (status=1). Lines depict the corresponding linear fit, and the shaded areas the 95% confidence interval. **(b)** Volume distributions of the different tissues of dead (orange) and alive (green) patients across multiple survival thresholds. Black squares and lines show the median and third quartiles, respectively. In black text, the p-values of each comparison (‘***’ p<0.001, ‘**’ p<0.01, ‘*’ p<0.05, and ‘n.s.’ p>0.05, two-sided Mann-Whitney U-tests). In blue, BB corrected p-values. **(c)** Mann-Whitney U-tests, also shown in **(b)**, for different survival thresholds comparing the sizes of all the tumor tissues. Each p-value tests for differences in tissue sizes in patients who died before a given threshold. Raw and BB corrected p-values are shown in black and blue, respectively. Dashed horizontal lines depict different significance thresholds (red: p<0.05, gray: p<0.01, p<0.001, and p<0.0001, respectively).

**Figure S3.**
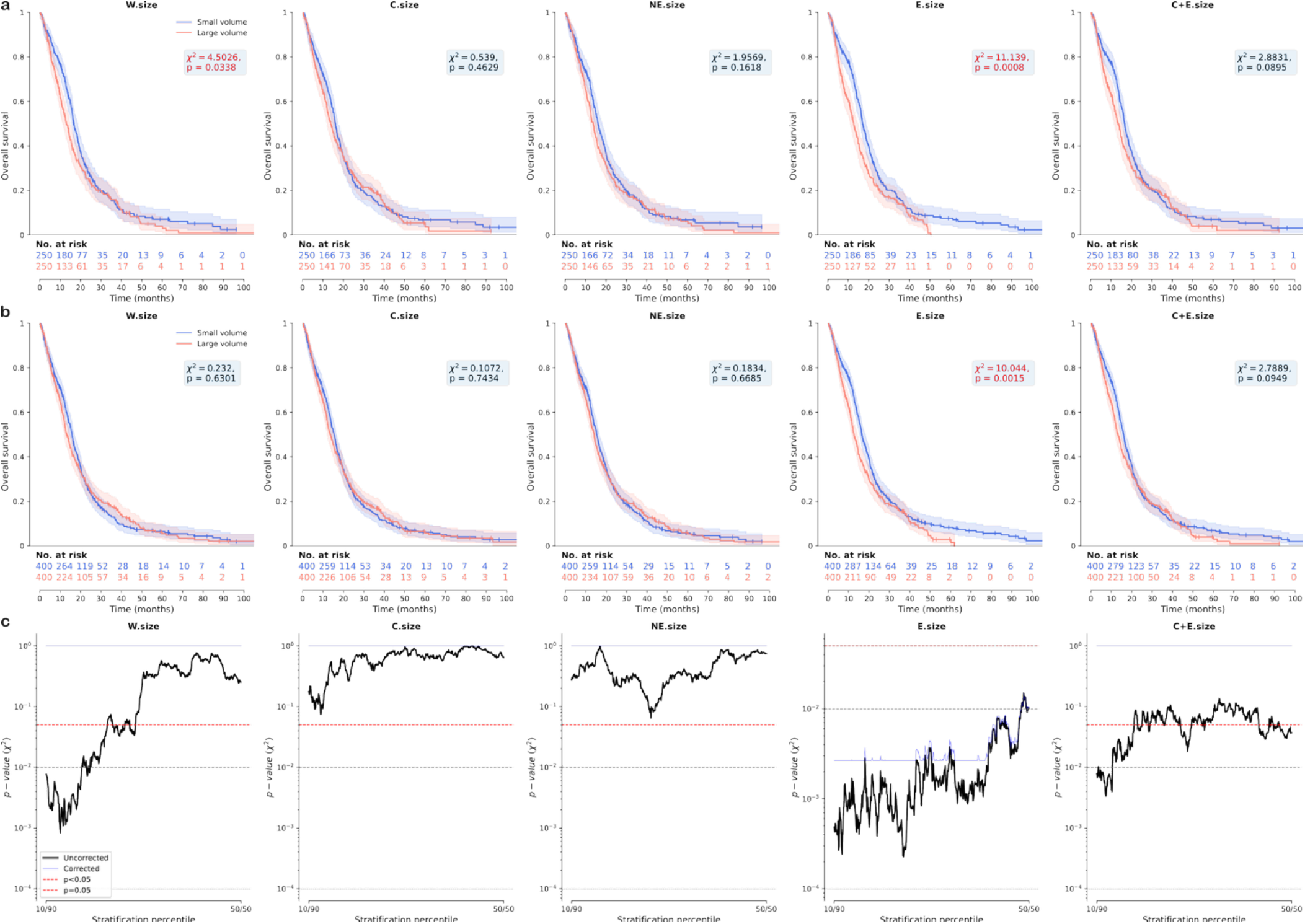
Kaplan-Meier curves and survival rates stratified by tissue volume. **(a)** Kaplan-Meier curves for the two strata at the 25th and 75th percentiles of the different tumor tissue volumes. The shaded areas correspond to the 95% CI. Small vertical dashes indicate right-censored entries. Two-sided log-rank tests were uncorrected. **(b)** Identical to **(a)**, but stratifying according to the 40th and 60th percentiles. **(c)** Two-sided log-rank tests, also shown in **(a-b)** for the 25/75 and 40/60 splits, comparing survival rates for a given stratification percentile across tissues. Raw and BB corrected p-values are shown in black and blue, respectively. Dashed horizontal lines depict different significance thresholds (red: p<0.05, gray: p<0.01, p<0.001, and p<0.0001, respectively).

**Figure S4.**
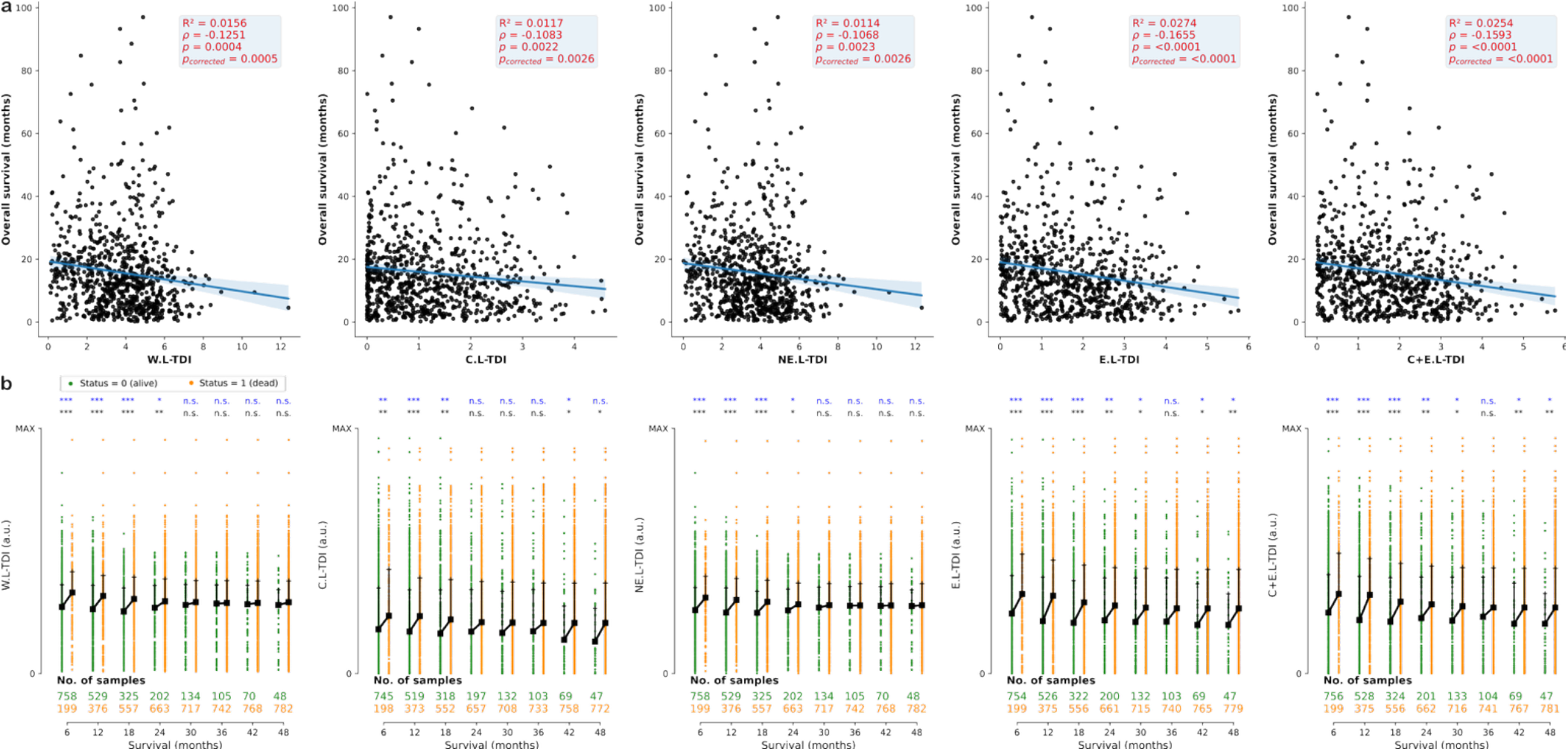
Associations between survival and the L-TDI derived from the different tumor tissues. L-TDIs derived from whole tumor (W.L-TDI), the core or necrosis (C.L-TDI); the non-enhancing or edema (NE.L-TDI); the enhancing tissue (E.L-TDI); and the combined core and enhancing tissues (C+E.L-TDI). **(a)** Linear correlation between the overall survival (OS) and the L-TDI derived from the different tissues (FDR corrected exact test). Only patients who experienced an event were considered (status=1). Lines depict the corresponding linear fit, and the shaded areas the 95% confidence interval. **(b)** Distributions of L-TDI values derived from the different tissues of dead (orange) and alive (green) patients across multiple survival thresholds. Black squares and lines show the median and third quartiles, respectively. In black text, the p-values of each comparison (‘***’ p<0.001, ‘**’ p<0.01, ‘*’ p<0.05, and ‘n.s.’ p>0.05, two-sided Mann-Whitney U-tests). In blue, BB corrected p-values.

**Figure S5.**
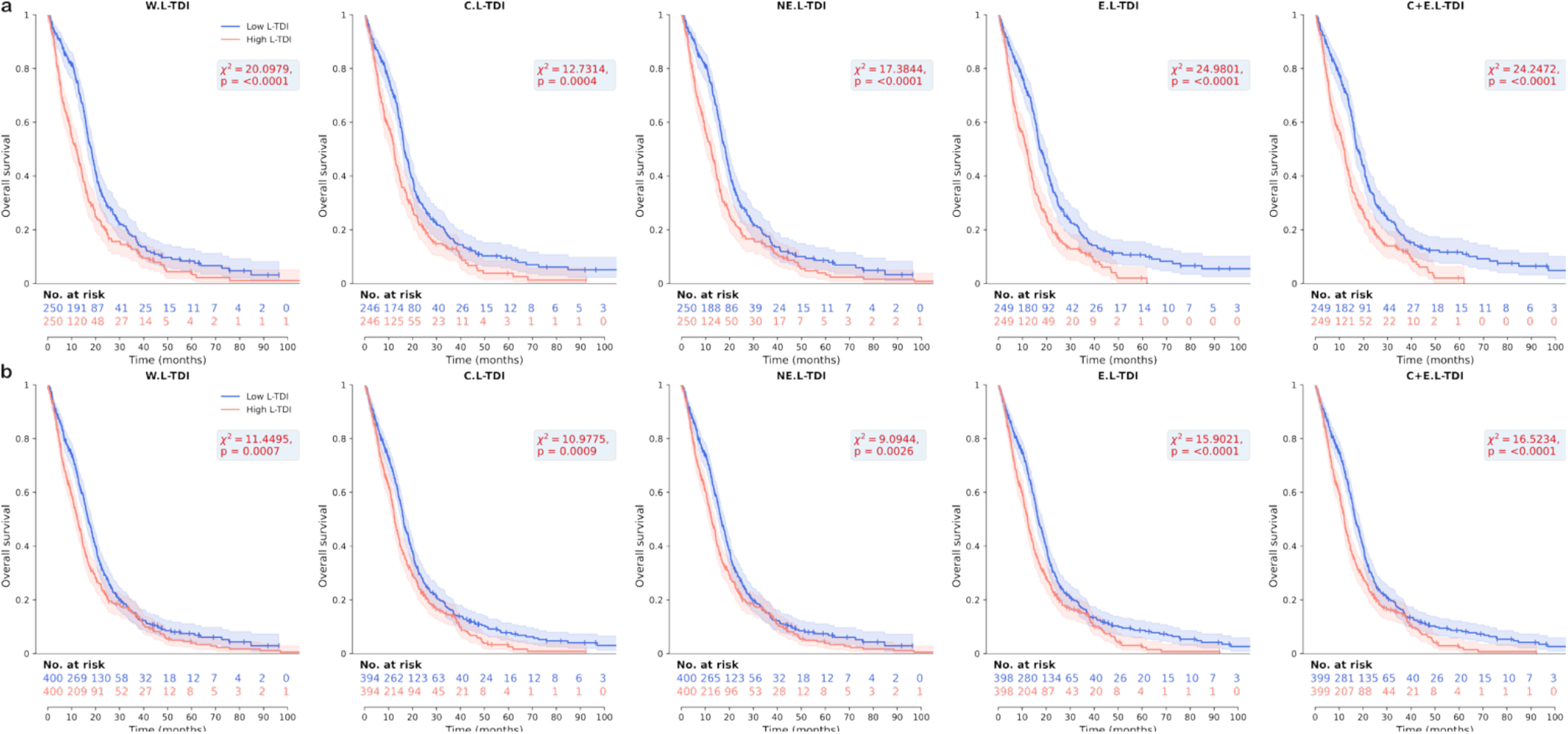
Kaplan-Meier curves and survival rates stratified by the L-TDI derived from the different tissues. **(a)** Kaplan-Meier curves for the two strata at the 25th and 75th percentiles of the L-TDI derived from the different tissues. Differences in survival rates were observed in all cases (p<0.01; two-sided log-rank tests). The shaded areas correspond to the 95% CI. Small vertical dashes indicate right-censored entries. **(b)** Identical to **(a)**, but stratifying according to the 40th and 60th percentiles.

**Figure S6.**
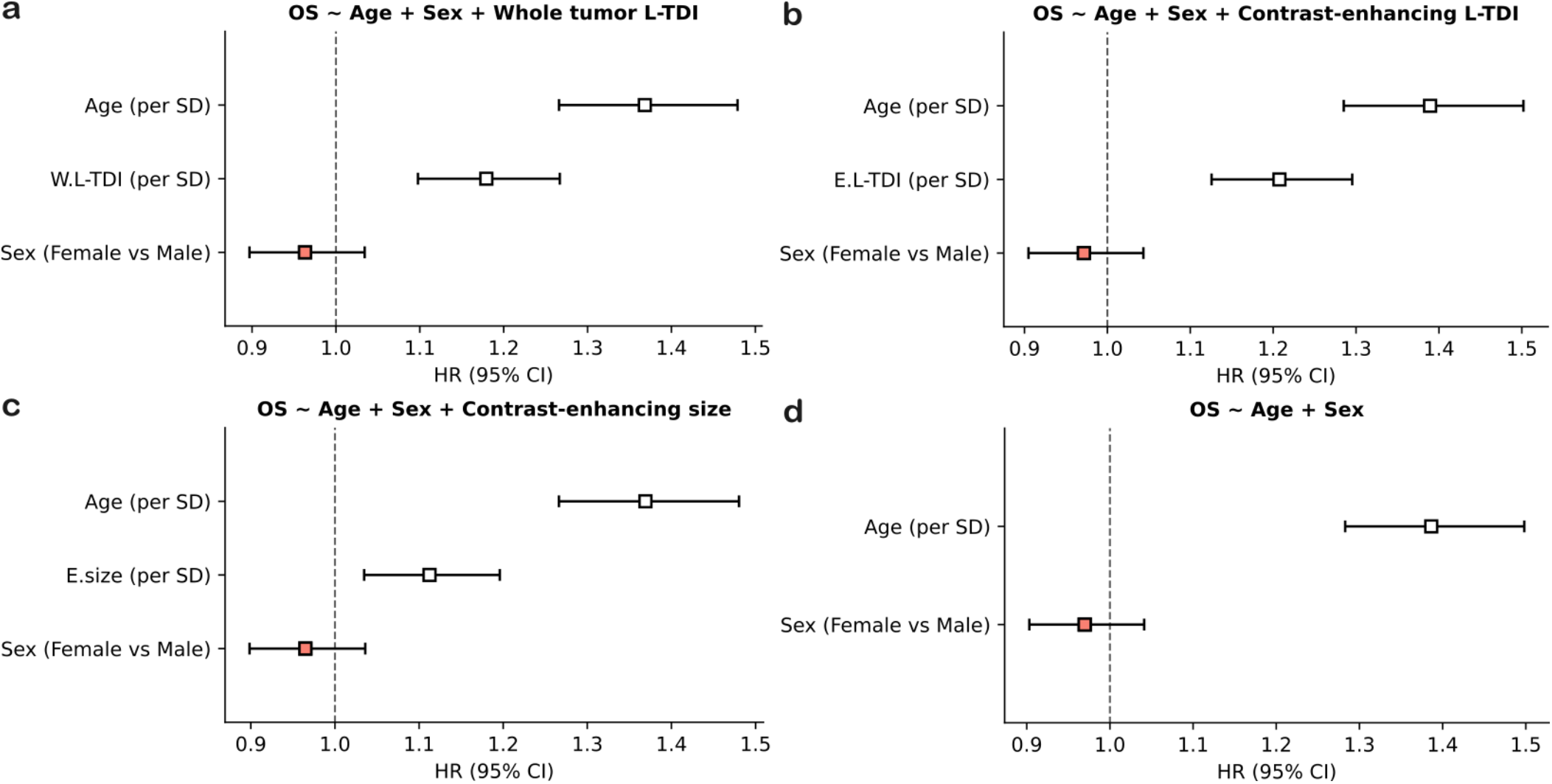
Multivariate Cox survival models with features available pre-surgically. Hazard ratios corresponding to models incorporating **(a)** the L-TDI derived from the whole tumor lesion mask, **(b)** the L-TDI derived from the contrast-enhancing tumor mask, **(c)** the size of the contrast-enhancing tumor, and **(d)** no additional imaging features. Black error bars depict hazard ratios with statistical significance (two-sided Wald’s z test), while red markers depict otherwise.

**Figure S7.**
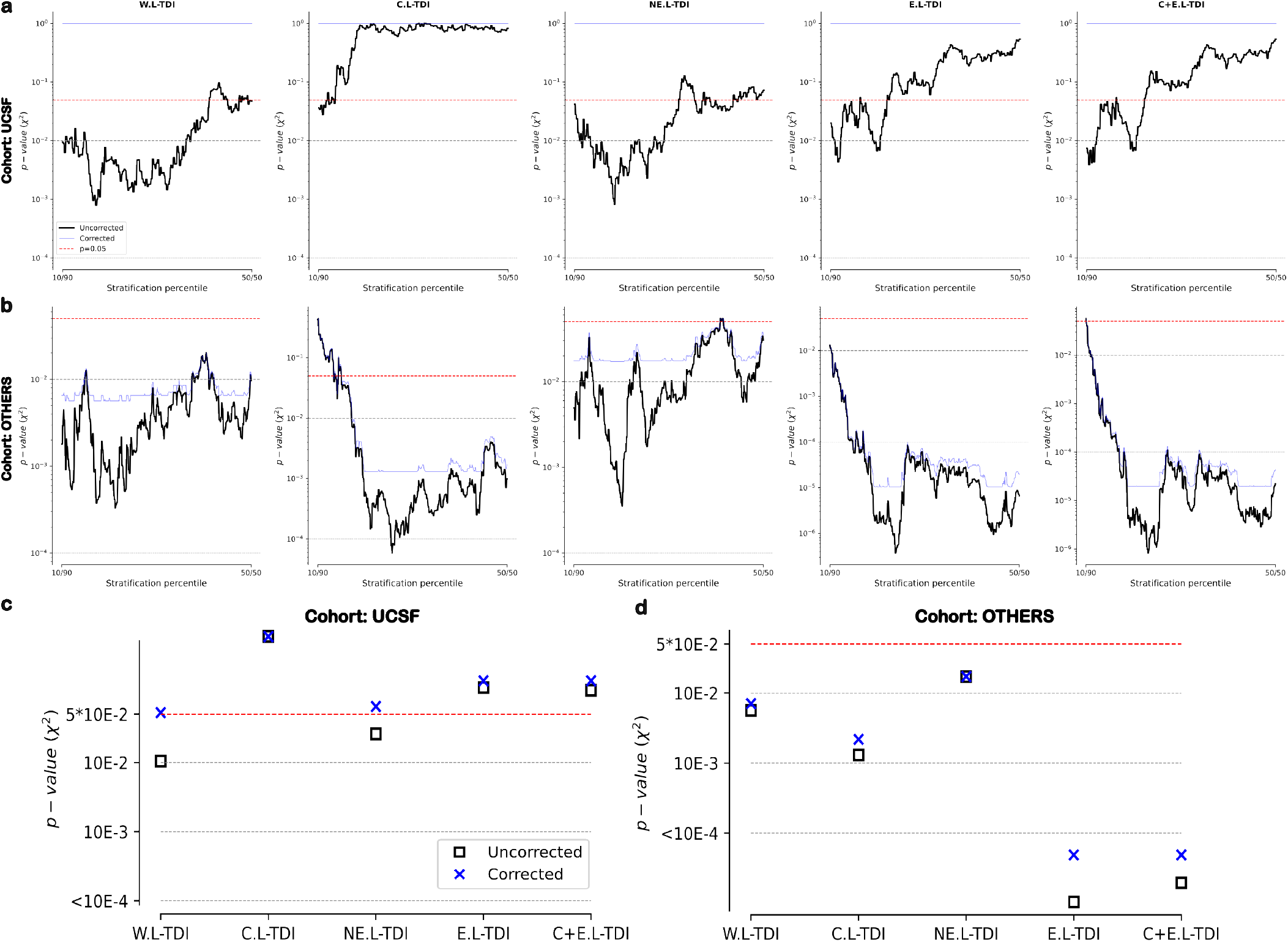
L-TDI and the Benjamini-Bogomolov procedure across independent cohorts. **(a-b)** For the UCSF **(a)** and OTHER **(b)** cohorts, two-sided log-rank tests comparing survival rates for a given stratification percentile across L-TDIs derived from all the tumor compartments. Raw and BB corrected p-values are shown in black and blue, respectively. The OTHER cohort contains patients from the UPENN, TCGA, and RHUH according to the results described in the main text. **(c-d)** For the UCSF **(c)** and OTHER **(d)** cohorts, Simes combined p-values (black squares), shown on a log scale for clarity. Blue crosses indicate the FDR corrected family-wise p-values. Dashed horizontal lines depict different significance thresholds (red: p<0.05, gray: p<0.01, p<0.001, and p<0.0001, respectively).

**Figure S8.**
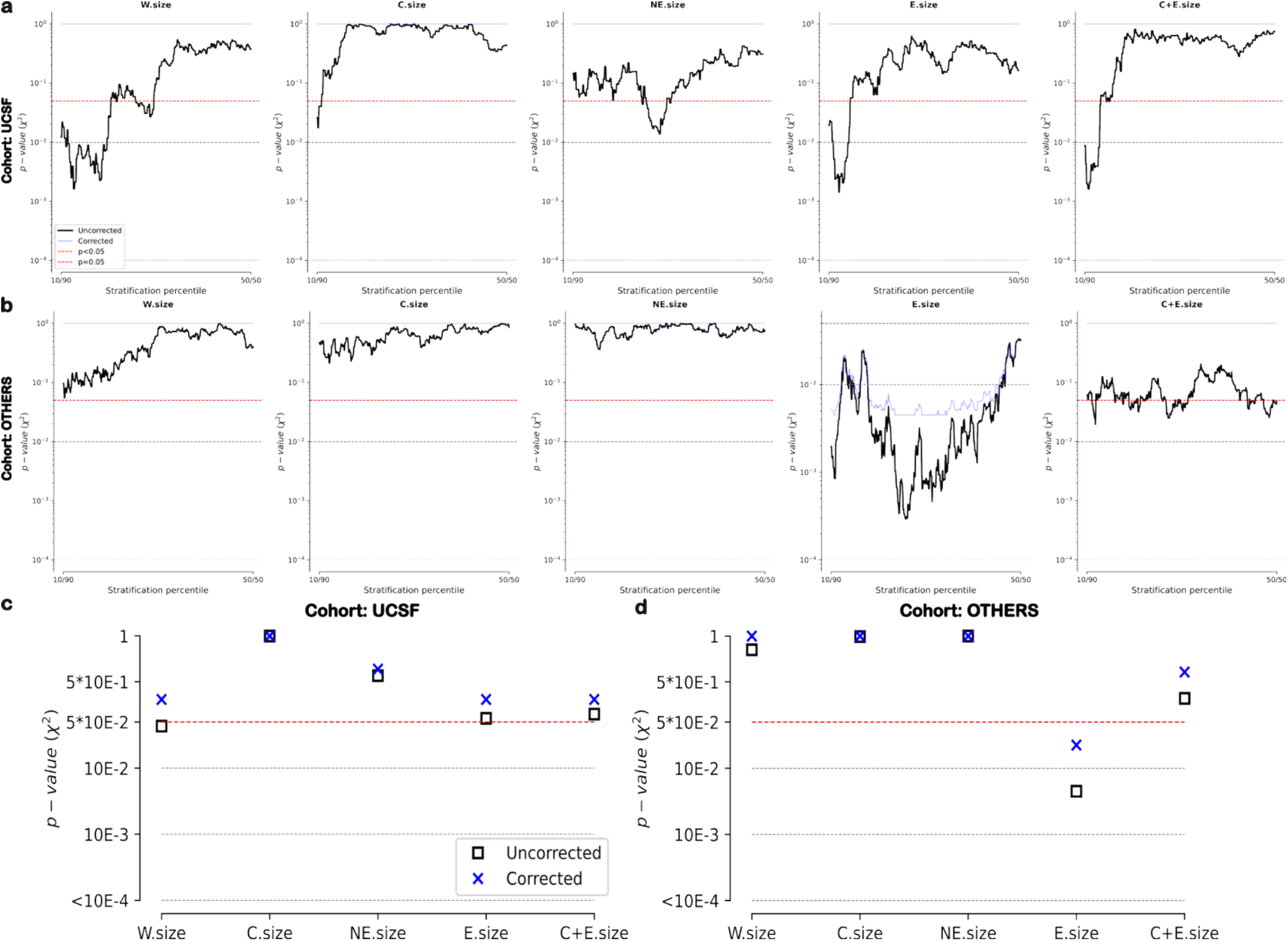
Tumor volume and the Benjamini-Bogomolov procedure across independent cohorts. **(a-b)** For the UCSF **(a)** and OTHER **(b)** cohorts, two-sided log-rank tests comparing survival rates for a given stratification percentile across tumor tissues. Raw and BB corrected p-values are shown in black and blue, respectively. The OTHER cohort contains patients from the UPENN, TCGA, and RHUH according to the results described in the main text. **(c-d)** For the UCSF **(c)** and OTHER **(d)** cohorts, Simes combined p-values (black squares), shown on a log scale for clarity. Blue crosses indicate the FDR corrected family-wise p-values. Dashed horizontal lines depict different significance thresholds (red: p<0.05, gray: p<0.01, p<0.001, and p<0.0001, respectively).

---

**Table S1.** Univariate Harrell and Uno’s concordance indices. Companion to Table 2 in the main text, it shows that the effect of right-censoring on the concordance index estimates remains very small without altering the main conclusions, thus highlighting the improved ranking ability of the L-TDI framework.

| Tissue feature | Harrell's C-index | Uno's C-index |
| --- | --- | --- |
| W.size | 0.5274 | 0.5252 |
| C.size | 0.5127 | 0.5112 |
| NE.size | 0.5159 | 0.5147 |
| E.size | 0.5487 | 0.5451 |
| C+E.size | 0.5355 | 0.5325 |
| W.L-TDI | 0.5646 | 0.5593 |
| C.L-TDI | 0.5450 | 0.5424 |
| NE.L-TDI | 0.5594 | 0.5544 |
| E.L-TDI | 0.5649 | 0.5609 |
| C+E.L-TDI | 0.5640 | 0.5598 |

**Table S2.**
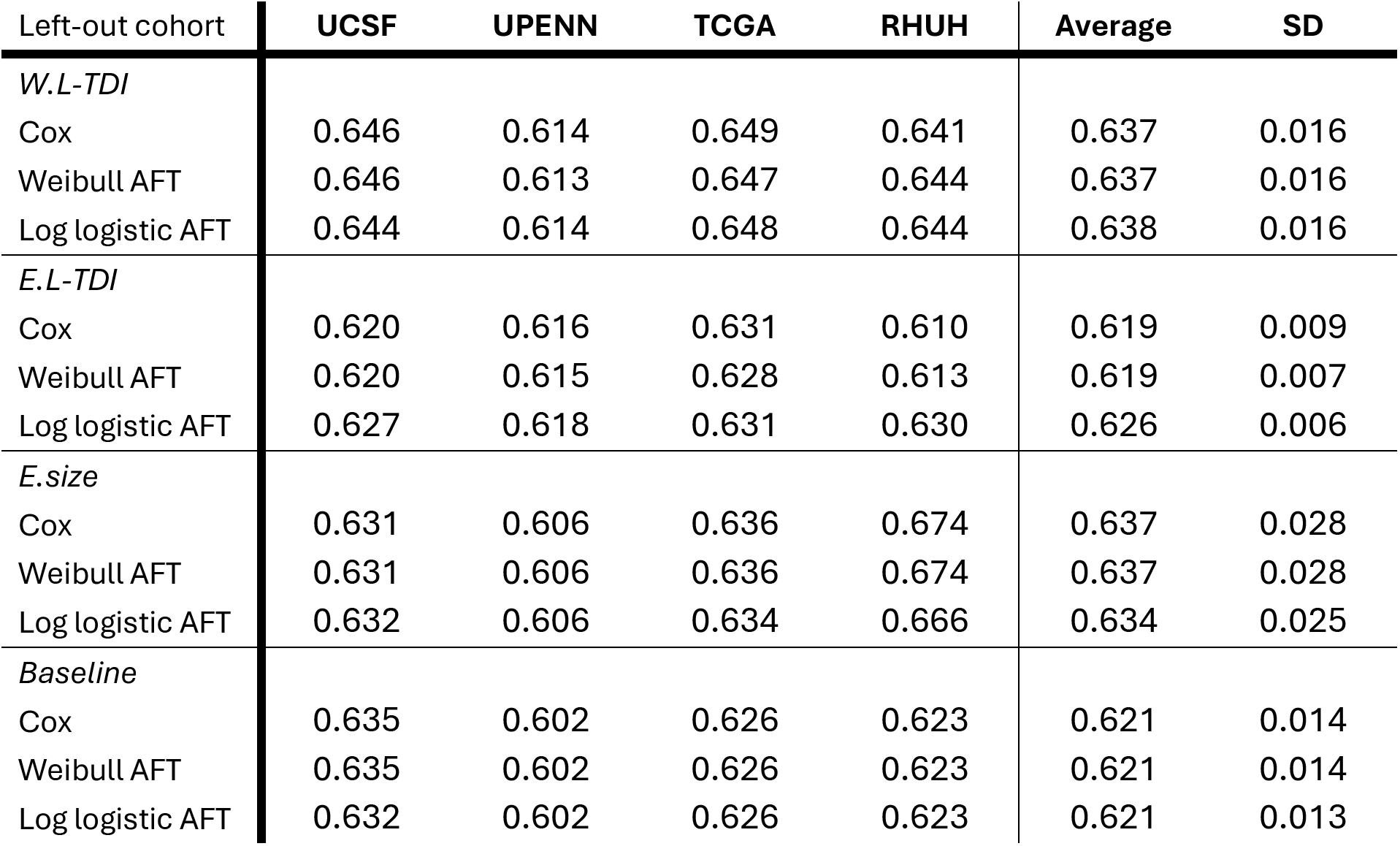
Leave-one-cohort-out concordance indices. Harrell C-indices computed on the left-out cohort (i.e., test) using three different survival models. The baseline models were fit using only age and biological factors as covariates of interest, while the rest were fit with the imaging feature of interest. The average corresponds to the arithmetic mean of the 4 results, and SD stands for “standard deviation”. The choice of the survival model had no apparent effect on the outcome. The largest increases came from the inclusion of the W.L-TDI and E.size features. Crucially, the variability when considering the size of the contrast-enhancing compartment was larger compared to the baseline and W.L-TDI models. This again demonstrates an increased robustness of the connectomic description of tumor burden, here instantiated by the L-TDI.

**Table S3.**
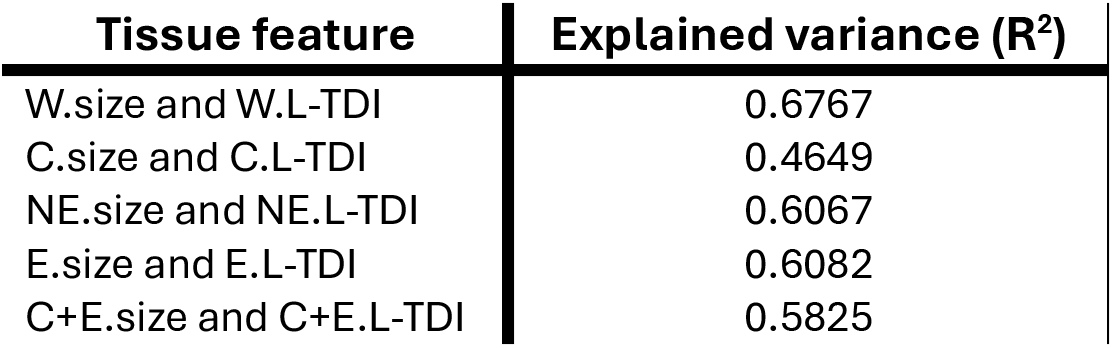
Explained variance (R^2^) between the size and L-TDI for each tumor compartment. Explained variance was computed assuming a linear dependence between the size of a given tumor compartment and the corresponding L-TDI. In this case, the R^2^ coefficient is equivalent to the square of the Pearson correlation between the two measurements.

| Tissue feature | Explained variance ($R^2$) |
| --- | --- |
| W.size and W.L-TDI | 0.6767 |
| C.size and C.L-TDI | 0.4649 |
| NE.size and NE.L-TDI | 0.6067 |
| E.size and E.L-TDI | 0.6082 |
| C+E.size and C+E.L-TDI | 0.5825 |

