## Supplementary material B and C for "Beyond tumor volume: connectomic measures of tumor burden provide superior prognostic information across tumor compartments in glioblastoma": Falco_Roget_et_al_Suppl-B.pdf

### UCSF, UPENN, TCGA, RHUH -- pooled database and site correction

```
NormWM_4_Glioblastoma/createDatabase.py /home/joan/Desktop/PROJECTS/Glioblastomas RESULTS-GBM_4-cohorts_Tissues --idh W
T --grade IV --stream-th 0 --format both --seed 42 --pairwise --log --cohorts UCSF UPENN TCGA RHUH --adjust-covariates
age sex eor mgmt --site-reference UCSF --tipping-plausible 2 --n-perms 5000
```

#### 1. Cohorts

Partition of the cohorts into the two site groups (default). Group 0 is the reference, whose survival times are left untouched; group 1's are rescaled by the fitted coefficient.

| cohort | id | n | events | pct_censored | site | site_group |
|---|---|---|---|---|---|---|
| UCSF | 0 | 367 | 223 | 39.2 | 0 | UCSF |
| UPENN | 1 | 496 | 482 | 2.8 | 1 | UPENN+TCGA+RHUH |
| TCGA | 2 | 103 | 82 | 20.4 | 1 | UPENN+TCGA+RHUH |
| RHUH | 3 | 33 | 24 | 27.3 | 1 | UPENN+TCGA+RHUH |

#### 2. Survival before correction

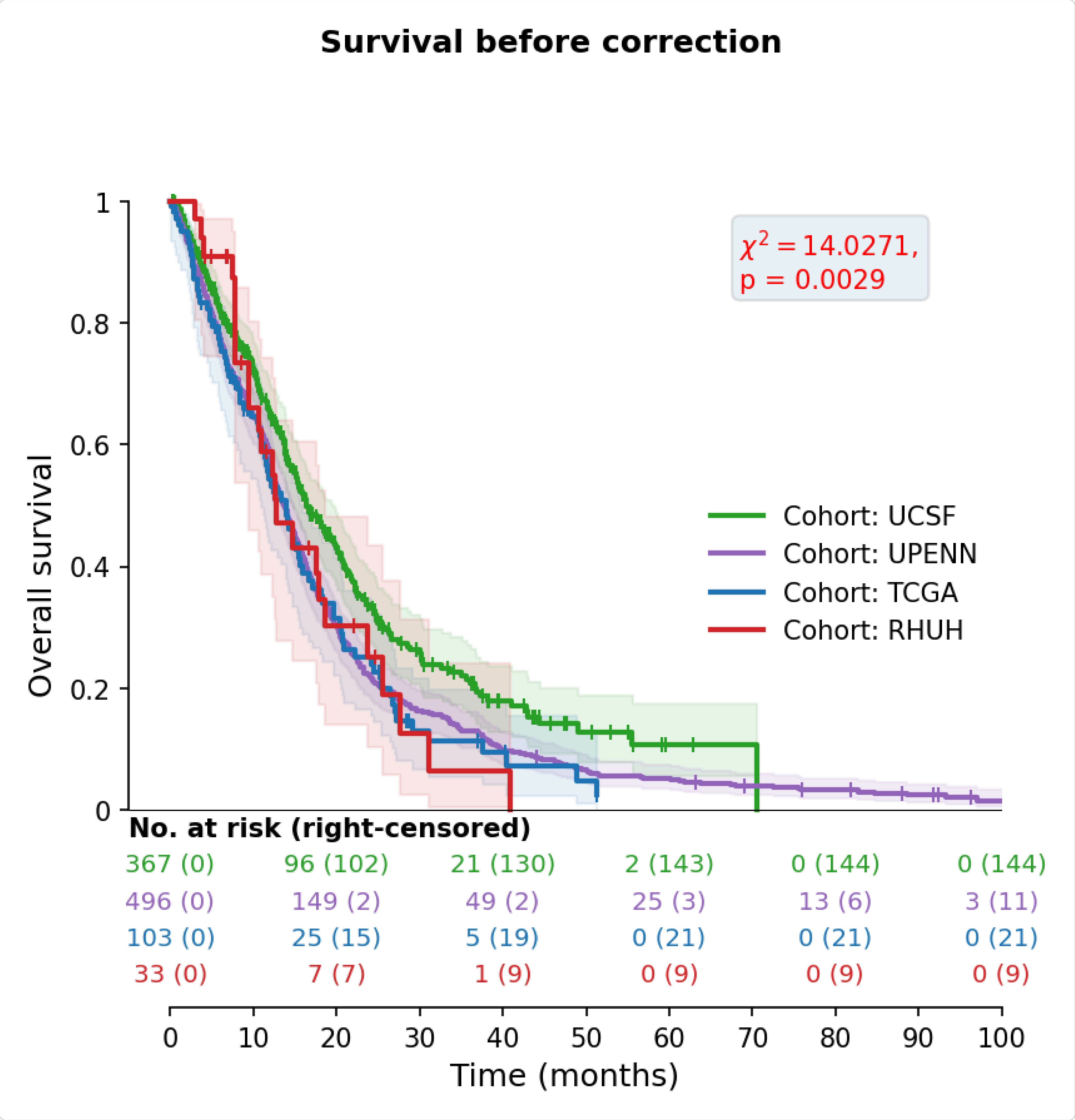


OS-cohorts_before-correction

#### 3. Pairwise cohort comparisons

One figure per pair of cohorts. The left panels diagnose the model whose coefficient the right panel applies, so an adjusted pair is diagnosed adjusted; the cloglog curves and their log-rank stay marginal, since a Kaplan-Meier curve has no covariates to hold fixed.

##### 3.1 UCSF vs UPENN

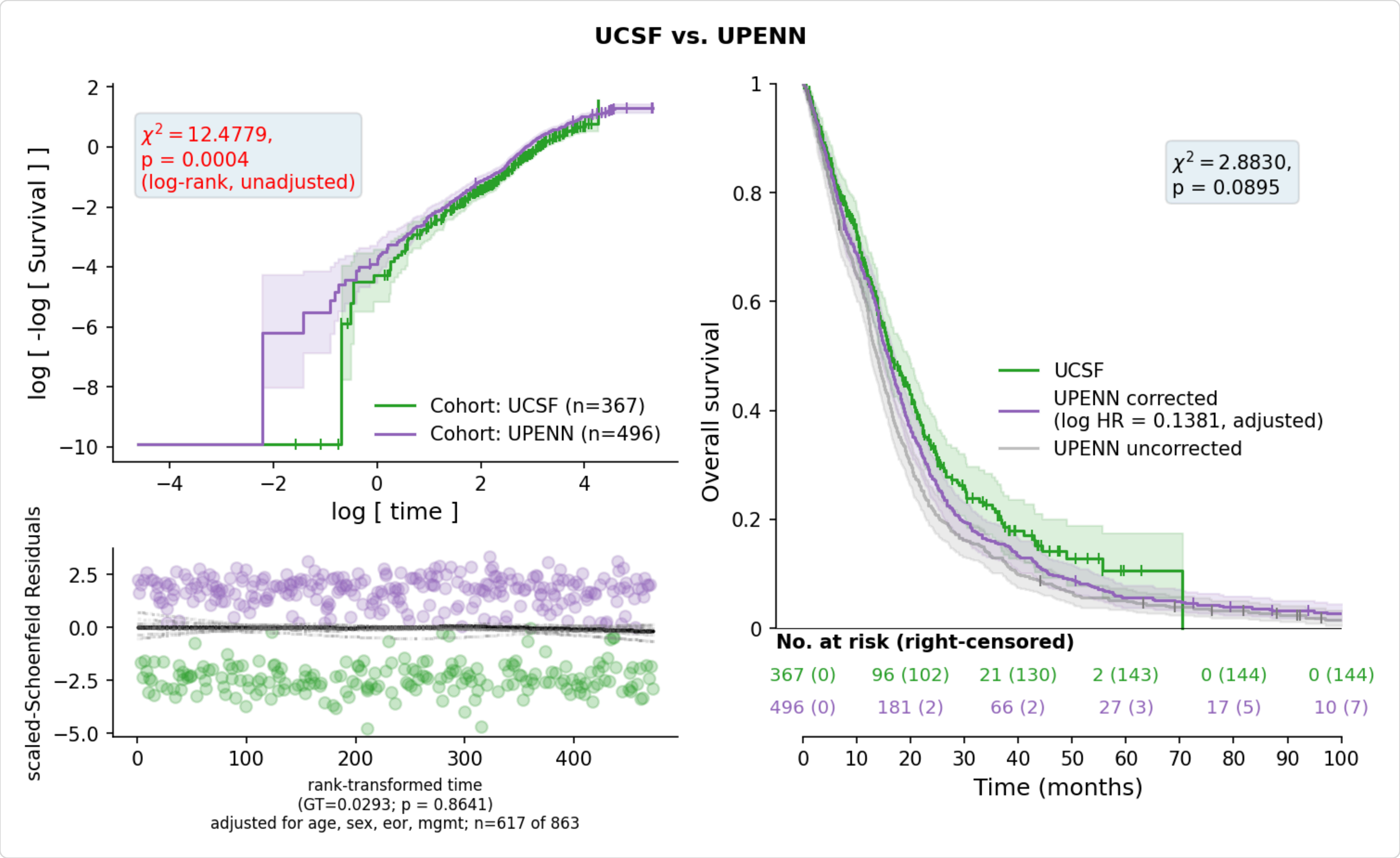


Site-effects_Survival-times_UCSF-UPENN_adjusted

###### 3.1.1 Proportional hazards: UCSF vs UPENN

Grambsch-Therneau [4] against rank-transformed time, for every term of the model the coefficient comes from -- not the site term alone. A term with a small p has an effect that drifts over follow-up, so the single coefficient reported for it is an average over the whole curve. Both the raw p and the Bonferroni-adjusted one are given: the raw p is one term's evidence, and p_bonferroni reads it against the 7 terms tested at once. A term is flagged, and described over time below, on the adjusted p -- the whole model is screened in one pass, so the raw p of the worst term is not the evidence it appears to be. lifelines offers no equivalent of R's cox.zph global row, so the smallest adjusted p stands in for it.

| term | test_statistic | p | p_bonferroni | violates |
|---|---|---|---|---|
| Age (per year) | 13.9105 | 0.0002 | 0.0013 | True |
| MGMT: Intermediate | 12.1783 | 0.0005 | 0.0034 | True |
| MGMT: Methylated | 1.9787 | 0.1595 | 1.0000 | False |
| Sex: Female | 0.9551 | 0.3284 | 1.0000 | False |
| EOR: Subtotal (<90%) | 0.4298 | 0.5121 | 1.0000 | False |
| pair | 0.0216 | 0.8832 | 1.0000 | False |
| EOR: Gross total (>=90%) | 0.0056 | 0.9401 | 1.0000 | False |

2 term(s) fail the test. Each is refitted with a log-time interaction -- $\beta(t) = \beta + \theta \log(t/t_{\text{ref}})$, the data split at the event times -- and compared against the constant-coefficient model by likelihood ratio. $\theta$ is the change in log hazard ratio per unit of log time, $\beta$ the log hazard ratio at the median event time $t_{\text{ref}}$, and $\theta = 0$ is proportional hazards. The equations are in the Method section at the end.

| term | beta | theta | theta_ci_low | theta_ci_high | lr_chi2 | p | ref_months | rows | reason |
|---|---|---|---|---|---|---|---|---|---|
| Age (per year) | 0.0242 | -0.0188 | -0.0277 | -0.0100 | 17.8381 | <0.0001 | 12.7890 | 113501 | |
| MGMT: Intermediate | -0.2181 | -0.4103 | -0.7062 | -0.1145 | 6.9901 | 0.0082 | 12.7890 | 113501 | |

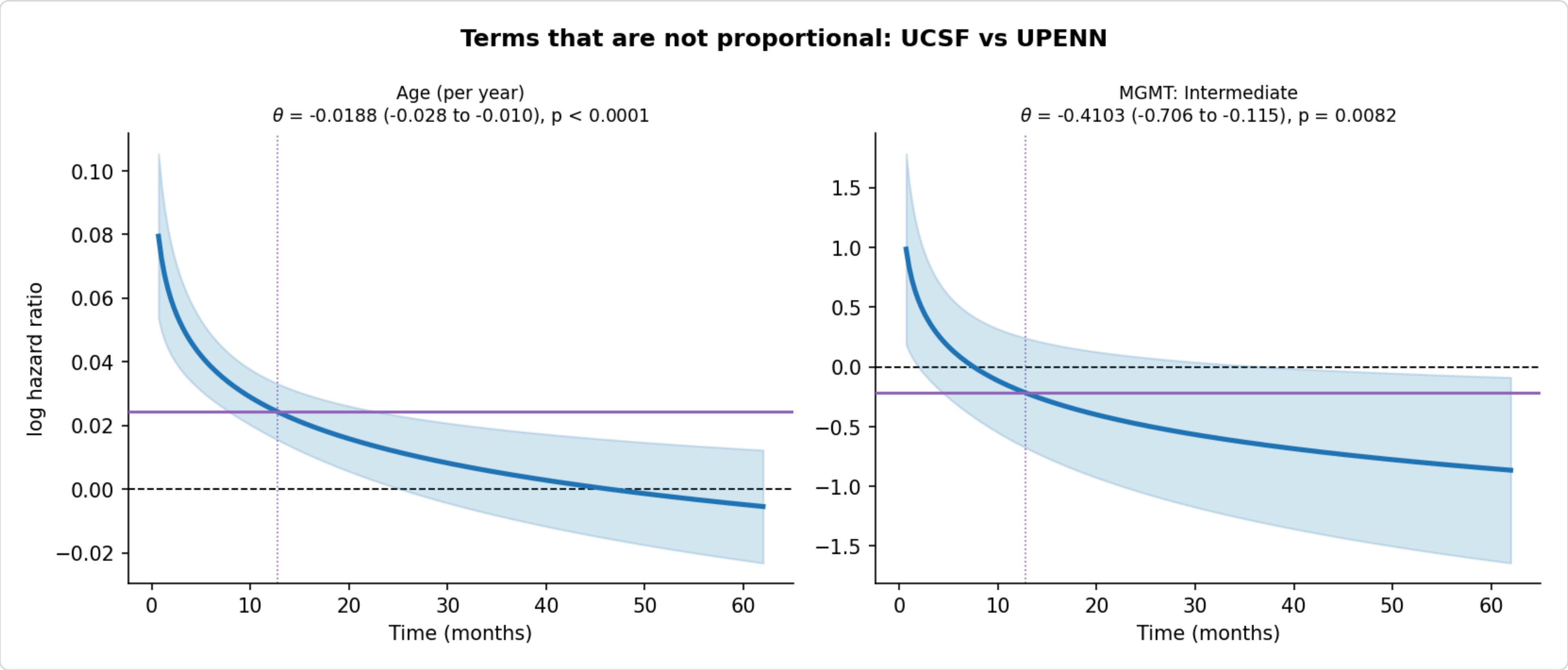


Site-effects_non-proportional-terms_UCSF-UPENN

##### 3.2 UCSF vs TCGA

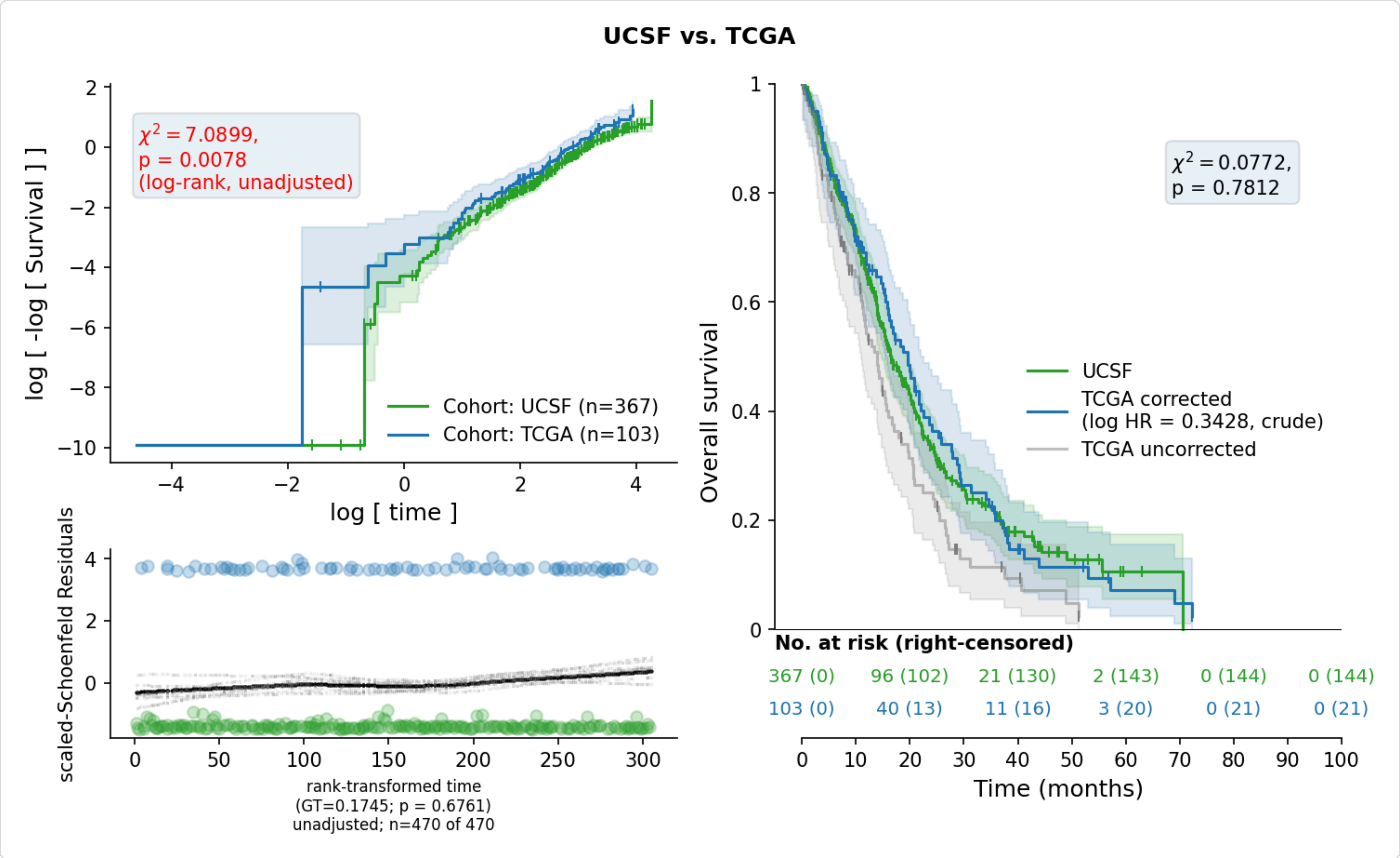


Site-effects_Survival-times_UCSF-TCGA

###### 3.2.1 Proportional hazards: UCSF vs TCGA

Not assessed: ConvergenceError: delta contains nan value(s). Convergence halted. Please see the following tips in the lifelines documentation: https://lifelines.readthedocs.io/en/latest/Exampl

##### 3.3 UCSF vs RHUH

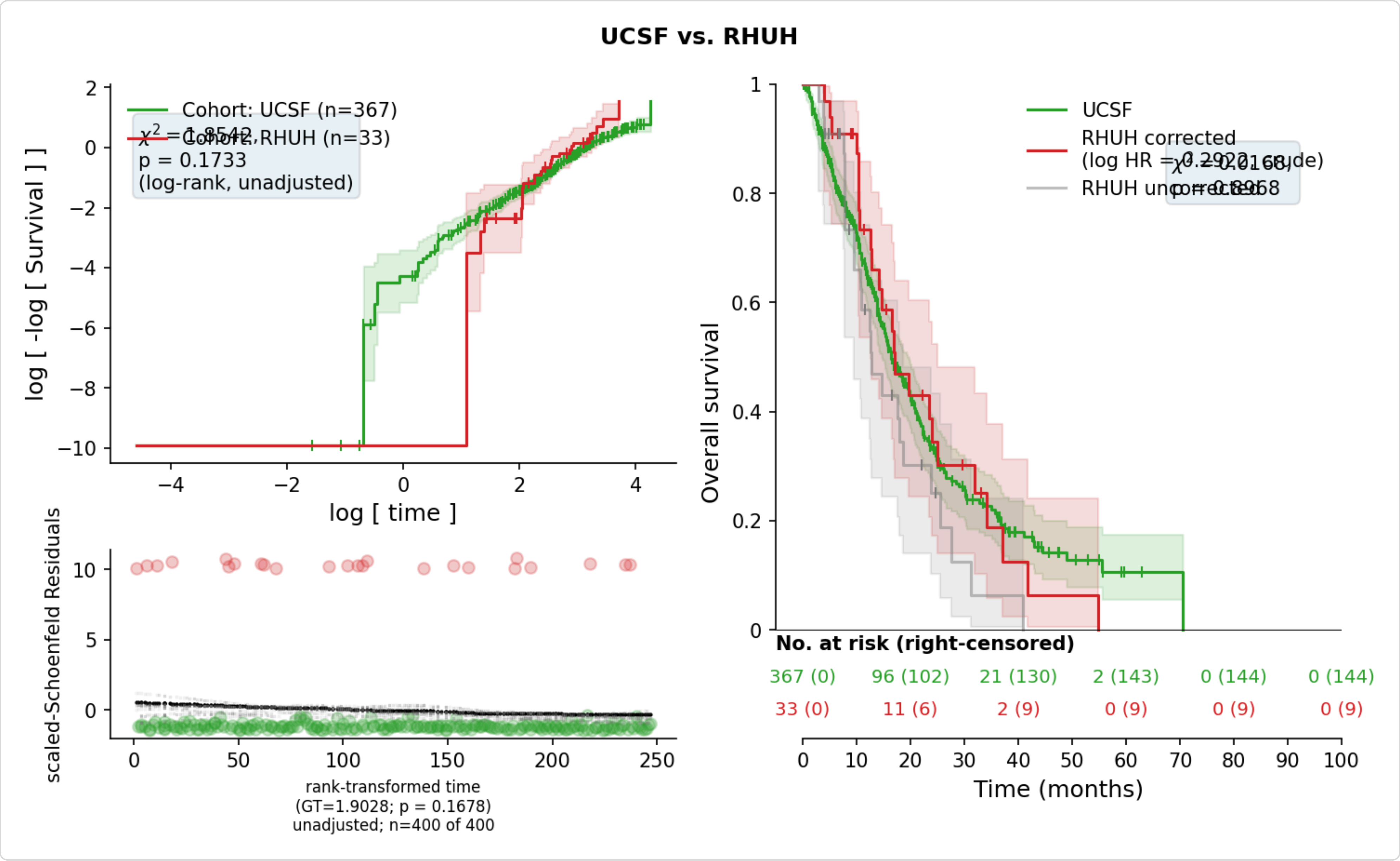


Site-effects_Survival-times_UCSF-RHUH

##### 3.3.1 Proportional hazards: UCSF vs RHUH

Not assessed: ConvergenceError: delta contains nan value(s). Convergence halted. Please see the following tips in the lifelines documentation: https://lifelines.readthedocs.io/en/latest/Exampl

#### 3.4 UPENN vs TCGA

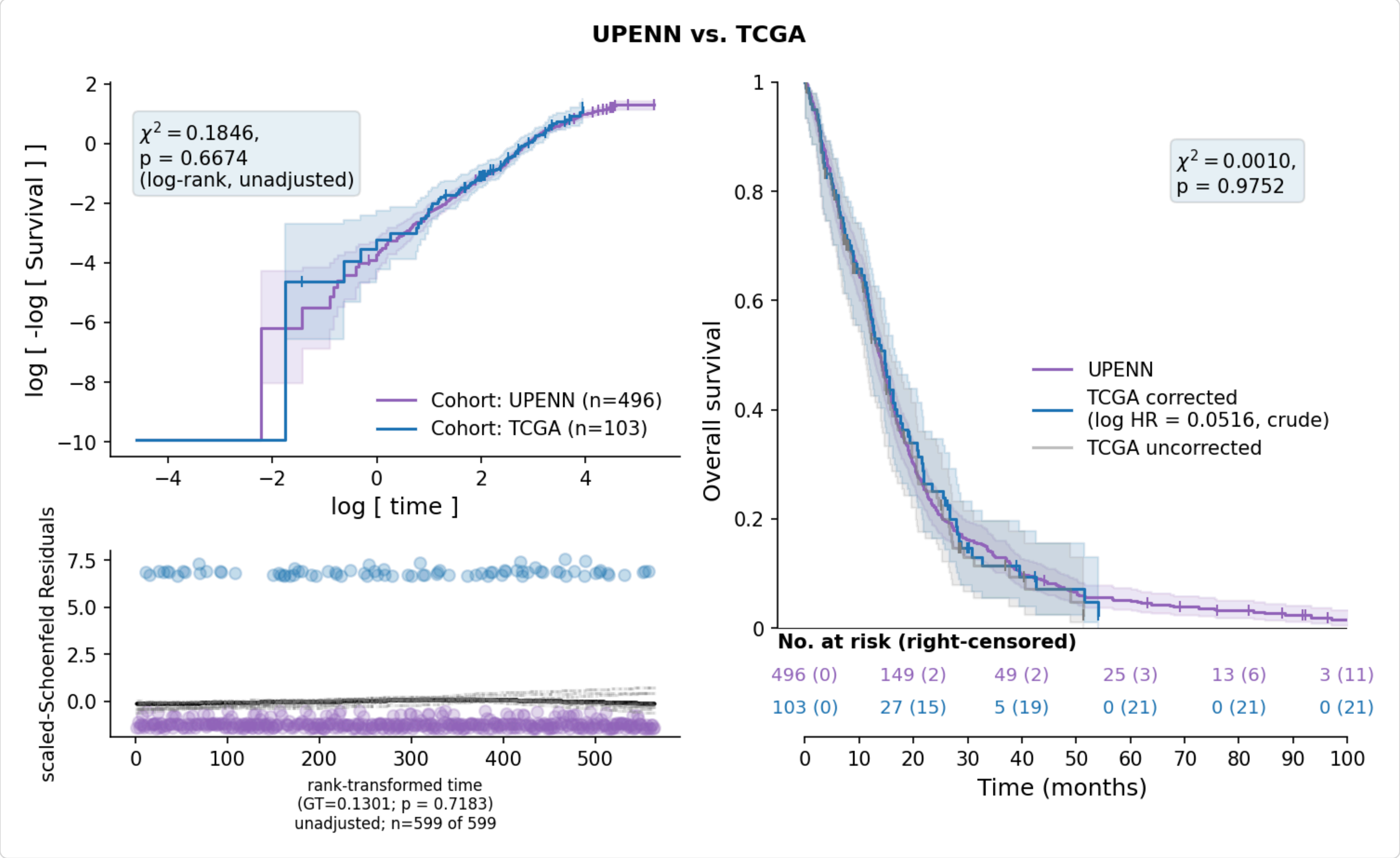


Site-effects_Survival-times_UPENN-TCGA

##### 3.4.1 Proportional hazards: UPENN vs TCGA

Not assessed: ConvergenceError: delta contains nan value(s). Convergence halted. Please see the following tips in the lifelines documentation: https://lifelines.readthedocs.io/en/latest/Exampl

##### 3.5 UPENN vs RHUH

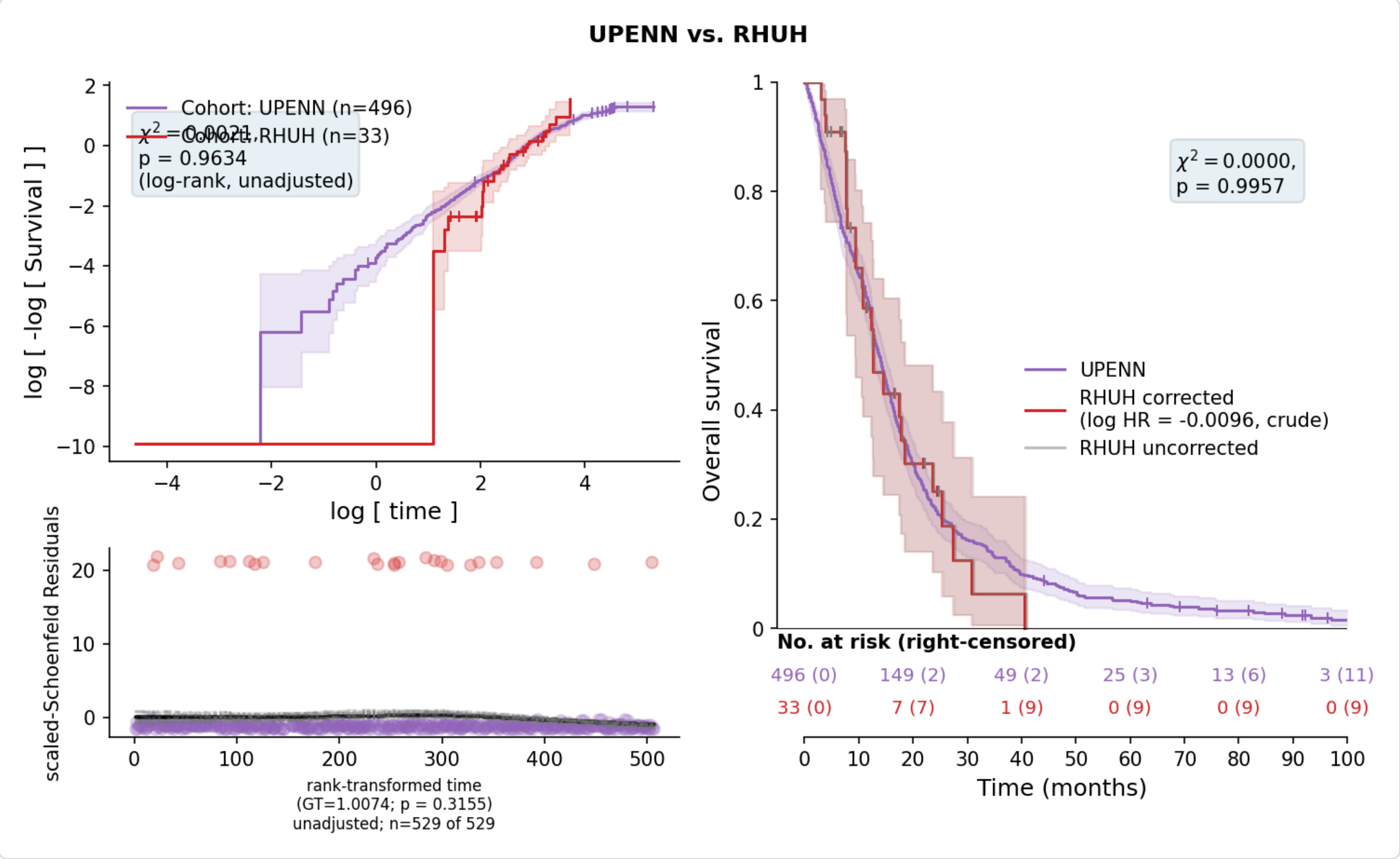


Site-effects_Survival-times_UPENN-RHUH

###### 3.5.1 Proportional hazards: UPENN vs RHUH

Not assessed: ConvergenceError: delta contains nan value(s). Convergence halted. Please see the following tips in the lifelines documentation: https://lifelines.readthedocs.io/en/latest/Exampl

##### 3.6 TCGA vs RHUH

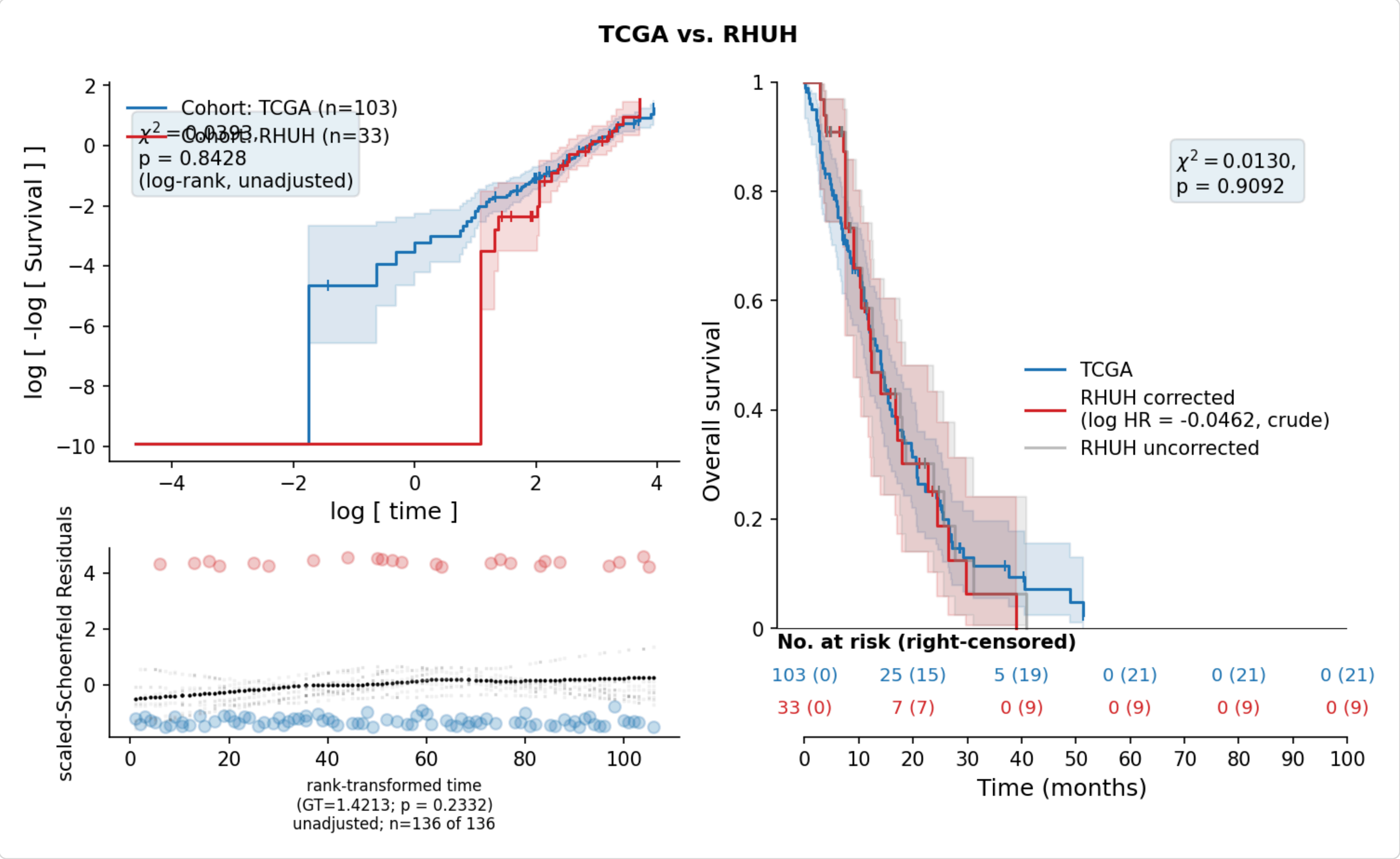

###### 3.6.1 Proportional hazards: TCGA vs RHUH

Not assessed: ConvergenceError: delta contains nan value(s). Convergence halted. Please see the following tips in the lifelines documentation: https://lifelines.readthedocs.io/en/latest/Exampl

---

#### 4. Site diagnostics: case-mix, entry point or censoring?

A survival difference between the site groups can come from three places. Case-mix: the groups enrolled different patients (older, fewer resections, less methylated MGMT). Entry point: the survival clock starts at a different event in one group, e.g. at the preoperative MRI in one and at diagnosis in another. Censoring: one group lost more of its patients to follow-up, and the patients it lost were not like those it kept. The three are not exclusive, and they call for different responses: case-mix is real and must be kept, entry point is an artefact the corrected column is meant to remove, and censoring is an artefact that no rescaling removes.

The sections below take them in turn. Balance and the adjustment ladder (the first two) measure case-mix: what is left of the site effect once the covariates are held fixed. The four follow-up and censoring sections ask whether censoring could produce that remainder. Entry point has no direct measurement in these tables -- nothing records when each cohort started its clock -- so it can only be a candidate for what is left once the other two are accounted for, alongside case-mix nobody recorded, differences in treatment after the clock starts, and chance. Both entry point and censoring tend to concentrate the difference in the first months, which the proportional-hazards section would show as a site effect that drifts over follow-up; they cannot be separated from each other with these data.

Covariates walked: age, sex, eor, mgmt. Site groups: 0 = UCSF; 1 = UPENN+TCGA+RHUH.

##### 4.1 Balance and missingness between site groups

SMD is the standardised mean difference: the difference between the two site groups' means divided by their pooled standard deviation (defined under Methods). For a categorical covariate it is computed per level, on the 0/1 indicator of that level, so the means are proportions. |SMD| > 0.10 marks an imbalance worth adjusting for. A covariate a site never records cannot be balanced by any adjustment, so the two pct_missing columns are read alongside the SMD.

| variable | level | site0 | site1 | smd | pct_missing_site0 | pct_missing_site1 |
|---|---|---|---|---|---|---|
| Age (per year) | mean (SD) | 61.8 (12.0) | 62.7 (11.7) | 0.074 | 0.000 | 0.000 |
| Sex | Male | 218 (59.4%) | 382 (60.4%) | 0.021 | 0.000 | 0.000 |
| Sex | Female | 149 (40.6%) | 250 (39.6%) | 0.021 | 0.000 | 0.000 |
| EOR | Biopsy | 41 (11.2%) | 0 (0.0%) | 0.501 | 0.000 | 20.095 |
| EOR | Subtotal (<90%) | 112 (30.5%) | 180 (35.6%) | 0.109 | 0.000 | 20.095 |
| EOR | Gross total (>=90%) | 214 (58.3%) | 325 (64.4%) | 0.124 | 0.000 | 20.095 |
| MGMT | Unmethylated | 104 (29.3%) | 188 (54.7%) | 0.531 | 3.270 | 45.570 |
| MGMT | Intermediate | 5 (1.4%) | 25 (7.3%) | 0.290 | 3.270 | 45.570 |
| MGMT | Methylated | 246 (69.3%) | 131 (38.1%) | 0.658 | 3.270 | 45.570 |

##### 4.2 Adjustment ladder

Every rung is fitted on the subjects reporting every covariate in the ladder, so the rows differ only in what is adjusted for and never in who is in the model. pct_of_crude_removed is measured against the complete-case crude rung, not the all-subjects one.

| model | n | events | logHR | HR | logHR_ci_low | logHR_ci_high | p | pct_of_crude_removed | ph_[illegible] |
|---|---|---|---|---|---|---|---|---|---|
| crude (all subjects) | 999 | 811 | 0.2922 | 1.3394 | 0.1374 | 0.4471 | 0.0002 | 3.8380 | 0.97[illegible] |
| crude (complete-case sample) | 617 | 472 | 0.3039 | 1.3551 | 0.1213 | 0.4864 | 0.0011 | 0.0000 | 0.86[illegible] |
| + age | 617 | 472 | 0.2409 | 1.2724 | 0.0571 | 0.4248 | 0.0102 | 20.7136 | 0.93[illegible] |
| + age + sex | 617 | 472 | 0.2316 | 1.2606 | 0.0471 | 0.4160 | 0.0139 | 23.8022 | 0.96[illegible] |
| + age + sex + eor | 617 | 472 | 0.2686 | 1.3082 | 0.0747 | 0.4625 | 0.0066 | 11.6061 | 0.47[illegible] |
| + age + sex + eor + mgmt | 617 | 472 | 0.1381 | 1.1481 | -0.0594 | 0.3356 | 0.1706 | 54.5632 | 0.88[illegible] |

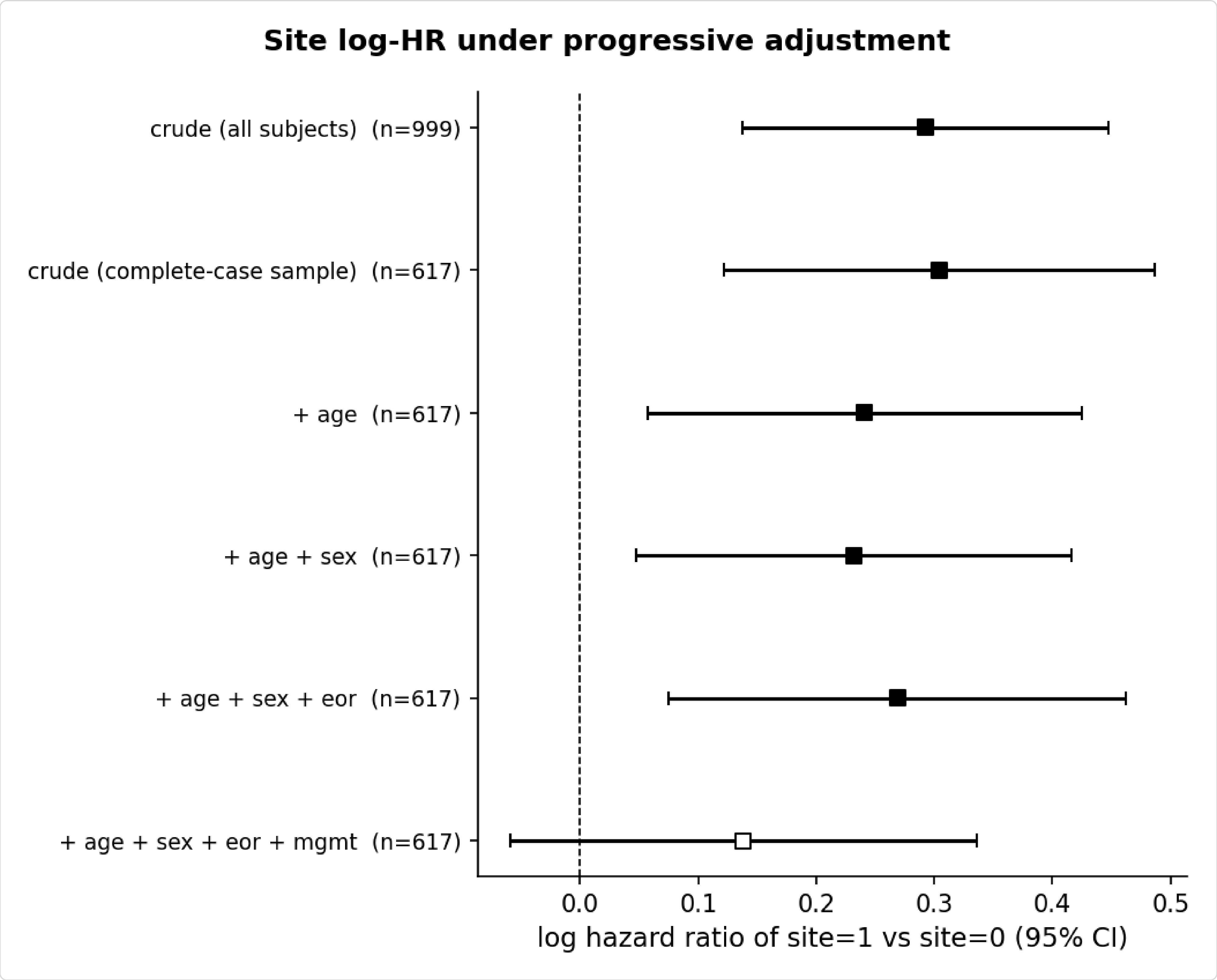


Site-diagnostics_adjustment-ladder

###### 4.3 Follow-up by site group (reverse Kaplan-Meier)

Median potential follow-up, i.e. the Kaplan-Meier estimator refitted with the event indicator flipped. A difference here is a difference in how long the groups were watched, not in how long they survived.

| group | n | events | pct_censored | median_OS_days | median_followup_days | max_followup_days | |
|---|---|---|---|---|---|---|---|
| UCSF | 367 | 223 | 39.2 | 499.0 | 798.0 | 2144.0 | |
| UPENN+TCGA+RHUH | 632 | 588 | 7.0 | 419.0 | 2808.0 | 6160.0 | * under 10% cer |

Log-rank test on the censoring distributions: chi2 = 182.8751, p < 0.0001.

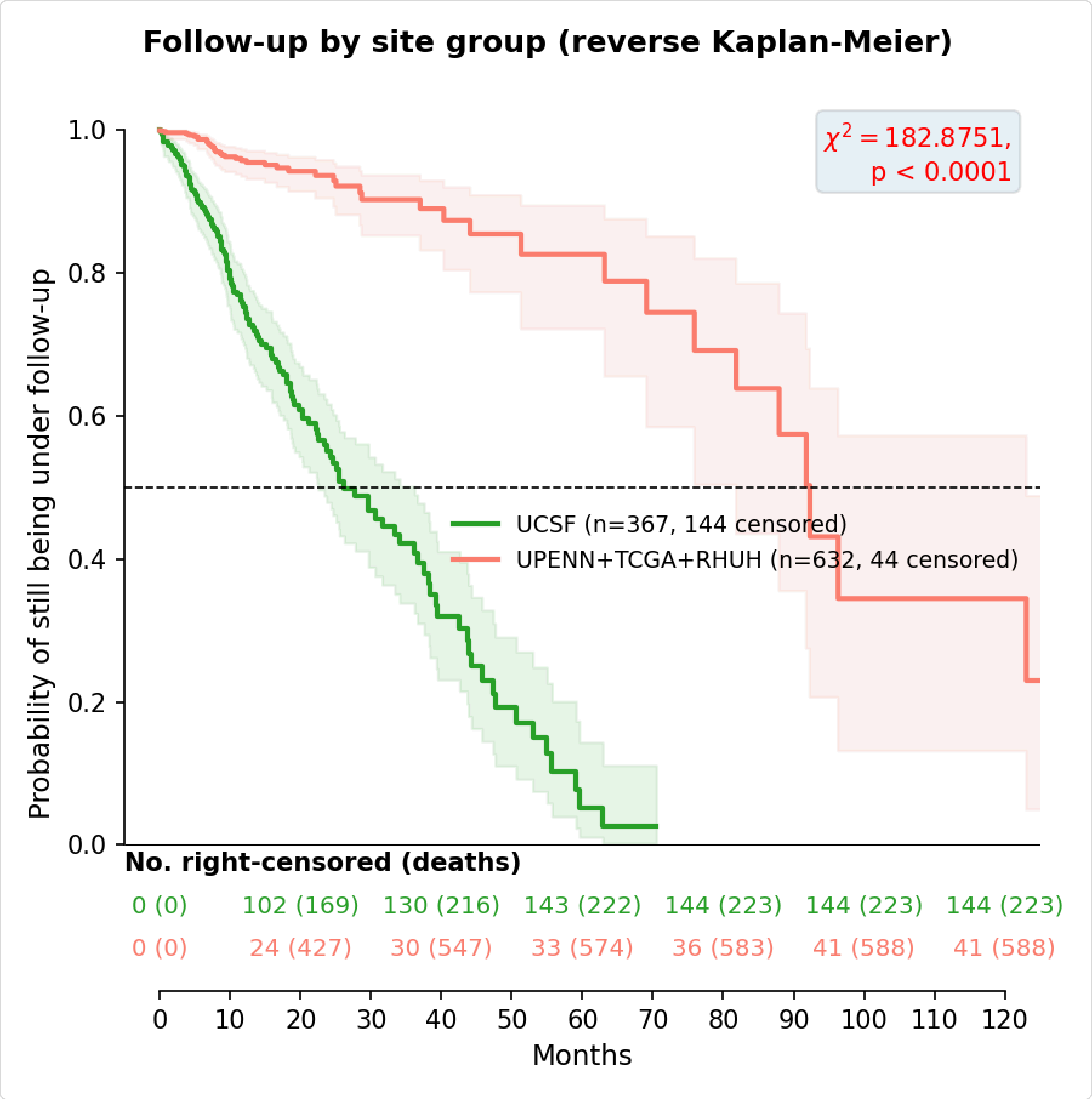


Site-diagnostics_reverse-KM

##### 4.4 Completeness of follow-up

The median follow-up above says how LONG each group was watched. This table says how COMPLETELY: of the follow-up each group owed up to a horizon (12 and 24 months), how much was actually observed. A patient who dies before the horizon has been followed completely -- their outcome is known -- so only patients censored before the horizon count as incomplete. Every column runs from 0 to 1, 1 meaning nobody was lost. Indented rows are the cohorts inside a site group that pools several.

lost_before_horizon: patients censored before the horizon, i.e. lost to follow-up.

percentage: the fraction of patients not lost. It treats a patient lost at month 11 as if lost at month 0, so it is the harshest reading.

reverse_km: the reverse Kaplan-Meier curve of the figure above, read at the horizon. It counts every death as a loss of follow-up too, so a group with many early deaths looks poorly followed; shown for comparison only.

CCI: Clark's completeness index [12]: observed follow-up time divided by the time that would have been observed had nobody been lost, assuming each lost patient would have lived to the horizon. That assumption owes them too much time, so CCI is a lower bound.

SPT: Xue's simplified person-time rate [11]: lost patients credited with the time they were actually seen, every other patient with the whole horizon. It credits deaths with time they did not live, so it is a slight upper bound.

FPT: Xue's formal person-time rate [11]: observed follow-up time divided by the time owed had nobody been lost, with the time owed estimated from the group's own survival curve. The best single estimate; it lies between CCI and SPT.

| group | horizon_months | n | lost_before_horizon | percentage | reverse_km | CCI | SPT | FPT |
|---|---|---|---|---|---|---|---|---|
| UCSF | 12 | 367 | 74 | 0.798 | 0.753 | 0.885 | 0.903 | 0.898 |
| UCSF | 24 | 367 | 110 | 0.700 | 0.551 | 0.749 | 0.822 | 0.810 |
| UPENN+TCGA+RHUH | 12 | 632 | 20 | 0.968 | 0.958 | 0.982 | 0.986 | 0.984 |

| group | horizon_months | n | lost_before_horizon | percentage | reverse_km | CCI | SPT | FPT |
|---|---|---|---|---|---|---|---|---|
| UPENN+TCGA+RHUH | 24 | 632 | 25 | 0.960 | 0.937 | 0.957 | 0.975 | 0.973 |
| UPENN | 12 | 496 | 2 | 0.996 | 0.995 | 0.996 | 0.997 | 0.997 |
| UPENN | 24 | 496 | 2 | 0.996 | 0.995 | 0.994 | 0.997 | 0.997 |
| TCGA | 12 | 103 | 12 | 0.883 | 0.840 | 0.936 | 0.950 | 0.944 |
| TCGA | 24 | 103 | 15 | 0.854 | 0.772 | 0.845 | 0.906 | 0.897 |
| RHUH | 12 | 33 | 6 | 0.818 | 0.774 | 0.915 | 0.926 | 0.928 |
| RHUH | 24 | 33 | 8 | 0.758 | 0.603 | 0.793 | 0.861 | 0.865 |

What to look at: FPT, and the gap in it between the site groups. Groups with similar FPT lost similar shares of their follow-up, and censoring is unlikely to separate them. A group whose FPT is well below the other's (say 0.80 against 0.97) lost follow-up that the other did not, and its survival curve rests on the assumption that the patients it lost were no sicker than those it kept -- which the next section tests. The percentage and reverse_km columns will look worse than FPT; that is how they are built, not a second problem.

##### 4.5 What predicts censoring?

Were the patients lost to follow-up different from those who stayed? Within each site group, two Cox models on the same patients and covariates: one where the 'event' is being censored (HR_censoring), and the ordinary one where the event is death (HR_death).

Reading a row: a hazard ratio above 1 means the covariate makes that outcome come sooner, below 1 later. same_side = yes when both hazard ratios are above 1 or both below 1: the patients the covariate makes more likely to be lost are then also the ones it makes more likely to die. An HR_censoring of 1.03 per year of age next to an HR_death of 1.04 means older patients both drop out sooner and die sooner. When the covariates that predict censoring all do this, the patients lost to follow-up were, on average, the sicker ones.

| group | n | censored | term | HR_censoring | p_censoring | HR_death | p_death | same_side |
|---|---|---|---|---|---|---|---|---|
| UCSF | 355 | 139 | Age (per year) | 1.0344 | <0.0001 | 1.0356 | <0.0001 | yes |
| UCSF | 355 | 139 | Sex: Female | 0.6661 | 0.0255 | 0.7737 | 0.0755 | yes |
| UCSF | 355 | 139 | EOR: Subtotal (<90%) | 0.4807 | 0.0459 | 0.3227 | <0.0001 | yes |
| UCSF | 355 | 139 | EOR: Gross total (>=90%) | 0.3515 | 0.0032 | 0.1636 | <0.0001 | yes |
| UCSF | 355 | 139 | MGMT: Intermediate | 0.6387 | 0.6635 | 1.0905 | 0.8712 | no |
| UCSF | 355 | 139 | MGMT: Methylated | 0.8870 | 0.5407 | 0.7816 | 0.1046 | yes |
| UPENN+TCGA+RHUH | 262 | 6 | (not modelled) | n/a | n/a | n/a | n/a | n/a |

UCSF: do the covariates predict censoring at all? Likelihood ratio test against a model without them, chi2 = 27.3586 on 6 df, p = 0.0001.

UPENN+TCGA+RHUH is not modelled: only 6 censored among the complete cases; at least 10 needed. The model needs every covariate, so only complete cases enter it, and in this group they are UPENN 262 of 496; TCGA 0 of 103 (EOR never recorded); RHUH 0 of 33 (MGMT never recorded).

Why it matters. The adjusted site model conditions on these same covariates, so censoring that runs through them does not bias the adjusted site hazard ratio. It does bias every Kaplan-Meier curve, crude or corrected, since a curve conditions on nothing. And a group that loses its sicker patients on the recorded covariates may also be losing them on an unrecorded one, such as performance status, which no adjustment can reach; the next section asks how much that could matter.

##### 4.6 Tipping point for informative censoring

The question. A censored patient's death was never observed. Every estimate above assumes that, after being censored, they went on to die at the same rate as comparable patients who stayed in follow-up -- same covariates, same site. That is 'independent censoring', and the data cannot confirm it, because what happened after censoring is exactly what is missing. What can be done is to ask: if the censored patients had in fact died faster (or slower) than that, how much would the adjusted site effect change?

How it is done [13]. delta is how much faster than a comparable patient who stayed. delta = 1 is the independent-censoring assumption itself; delta = 2 says they died at twice the rate; delta = 0.5 at half. For one value of delta: (1) every censored patient of one site group is given a plausible death time, drawn from the adjusted Cox model with their own covariates and site, later than the day they were censored, and with the hazard after censoring multiplied by delta; (2) with those deaths now in the data, the adjusted site model is fitted again; (3) steps 1-2 are repeated 20 times, since the death times are drawn at random, and the site hazard ratios are combined by Rubin's rules [14], whose interval includes the spread between the repetitions. This is done for each value of delta, and separately for the censored patients of each site group, since which group lost its patients informatively is not known.

| censored_group | delta | imputed | imputations | logHR | HR | HR_ci_low | HR_ci_high | p |
|---|---|---|---|---|---|---|---|---|
| UCSF | 0.20 | 139 | 20 | 0.6111 | 1.8424 | 1.5033 | 2.2579 | <0.0001 |

| censored_group | delta | imputed | imputations | logHR | HR | HR_ci_low | HR_ci_high | p |
|---|---|---|---|---|---|---|---|---|
| UCSF | 0.33 | 139 | 20 | 0.4845 | 1.6234 | 1.3262 | 1.9873 | <0.0001 |
| UCSF | 0.50 | 139 | 20 | 0.3926 | 1.4808 | 1.2119 | 1.8094 | 0.0001 |
| UCSF | 0.75 | 139 | 20 | 0.2361 | 1.2663 | 1.0440 | 1.5359 | 0.0165 |
| UCSF | 1.00 | 139 | 20 | 0.1515 | 1.1636 | 0.9583 | 1.4130 | 0.1260 |
| UCSF | 1.25 | 139 | 20 | 0.0691 | 1.0715 | 0.8816 | 1.3025 | 0.4877 |
| UCSF | 1.50 | 139 | 20 | 0.0189 | 1.0190 | 0.8483 | 1.2241 | 0.8403 |
| UCSF | 2.00 | 139 | 20 | -0.0827 | 0.9206 | 0.7662 | 1.1062 | 0.3774 |
| UCSF | 3.00 | 139 | 20 | -0.1759 | 0.8387 | 0.7018 | 1.0023 | 0.0530 |
| UCSF | 5.00 | 139 | 20 | -0.2525 | 0.7769 | 0.6523 | 0.9252 | 0.0046 |
| UPENN+TCGA+RHUH | 0.20 | 5 | 20 | 0.1353 | 1.1449 | 0.9397 | 1.3950 | 0.1793 |
| UPENN+TCGA+RHUH | 0.33 | 5 | 20 | 0.1357 | 1.1454 | 0.9401 | 1.3955 | 0.1781 |
| UPENN+TCGA+RHUH | 0.50 | 5 | 20 | 0.1361 | 1.1458 | 0.9404 | 1.3960 | 0.1769 |
| UPENN+TCGA+RHUH | 0.75 | 5 | 20 | 0.1364 | 1.1462 | 0.9408 | 1.3965 | 0.1757 |
| UPENN+TCGA+RHUH | 1.00 | 5 | 20 | 0.1364 | 1.1462 | 0.9408 | 1.3964 | 0.1757 |
| UPENN+TCGA+RHUH | 1.25 | 5 | 20 | 0.1376 | 1.1475 | 0.9419 | 1.3980 | 0.1720 |
| UPENN+TCGA+RHUH | 1.50 | 5 | 20 | 0.1379 | 1.1478 | 0.9421 | 1.3984 | 0.1712 |
| UPENN+TCGA+RHUH | 2.00 | 5 | 20 | 0.1395 | 1.1496 | 0.9437 | 1.4005 | 0.1661 |
| UPENN+TCGA+RHUH | 3.00 | 5 | 20 | 0.1398 | 1.1500 | 0.9440 | 1.4010 | 0.1651 |
| UPENN+TCGA+RHUH | 5.00 | 5 | 20 | 0.1424 | 1.1531 | 0.9466 | 1.4046 | 0.1571 |

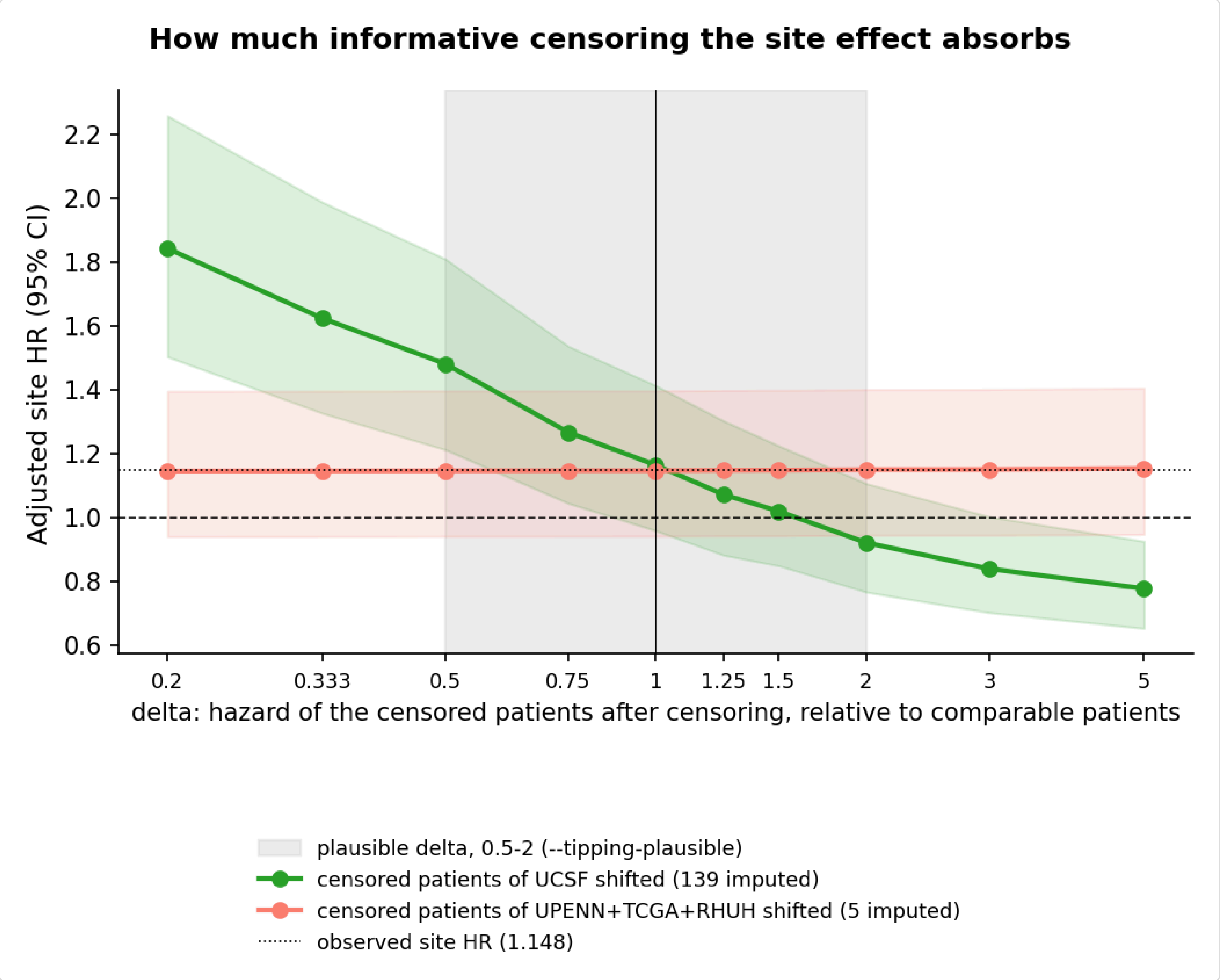


Site-diagnostics_censoring-tipping-point

How to read it. Each row is one delta for one group: 'imputed' is how many censored patients received a death time, and HR is the adjusted site hazard ratio after they did. At delta = 1 the HR should be close to the one actually estimated (1.1481, dotted line in the figure). The number to look for is the delta at which the HR reaches 1 (dashed line): how different the lost patients would have to be for the whole site effect to be a product of censoring. The shaded band, delta between 0.5 and 2, is the range this run was told to treat as plausible (--tipping-plausible, a judgement rather than an established threshold). If the HR reaches 1 inside the band, lost patients need only have died up to 2 times as fast as comparable patients who stayed for censoring to account for the whole site effect, so the site effect cannot be told apart from informative censoring. If it reaches 1 only outside the band, or not at all, censoring would have to be implausibly informative to explain it. A group with few censored patients barely moves the HR whatever delta is, because there is little to impute.

Censored patients of UCSF: the site HR reaches 1 at delta = 1.58, inside the band. If they died about 1.6 times as fast as comparable patients who stayed, there would be no site effect at all.

Censored patients of UPENN+TCGA+RHUH: the site HR does not reach 1 anywhere in delta = 0.2-5, the whole range tested.

What this can and cannot conclude. Inside the band, censoring is a sufficient explanation, so the site effect cannot be attributed to entry point. Outside the band, censoring is an unlikely sole explanation, but that does not make the remainder entry point: it rules out one source, not all others. The remainder could equally be case-mix nobody recorded (KPS is missing for all of UCSF), a genuine difference in treatment or care after the clock starts, or chance. And the analysis only has something to explain when the adjusted site effect is itself distinguishable from zero; when its confidence interval already covers HR = 1, the question is how easily censoring could have produced or hidden an effect, not where an established one came from. Finally, delta shifts every censored patient of a group equally. Censoring that is informative for some of them and not for others, as when some leave for hospice and others simply move, is not what the table describes.

##### 4.7 Proportional hazards: pooled site comparison

Grambsch-Therneau [4] against rank-transformed time, for every term of the model the coefficient comes from -- not the site term alone. A term with a small p has an effect that drifts over follow-up, so the single coefficient reported for it is an average over the whole curve. Both the raw p and the Bonferroni-adjusted one are given: the raw p is one term's evidence, and p_bonferroni reads it against the 7 terms tested at once. A term is flagged, and described over time below, on the adjusted p -- the whole model is screened in one pass, so the raw p of the worst term is not the evidence it appears to be. lifelines offers no equivalent of R's cox.zph global row, so the smallest adjusted p stands in for it.

| term | test_statistic | p | p_bonferroni | violates |
|---|---|---|---|---|
| Age (per year) | 13.9105 | 0.0002 | 0.0013 | True |
| MGMT: Intermediate | 12.1783 | 0.0005 | 0.0034 | True |
| MGMT: Methylated | 1.9787 | 0.1595 | 1.0000 | False |
| Sex: Female | 0.9551 | 0.3284 | 1.0000 | False |
| EOR: Subtotal (<90%) | 0.4298 | 0.5121 | 1.0000 | False |
| site | 0.0216 | 0.8832 | 1.0000 | False |
| EOR: Gross total (>=90%) | 0.0056 | 0.9401 | 1.0000 | False |

2 term(s) fail the test. Each is refitted with a log-time interaction -- $\beta(t) = \beta + \theta \log(t/t_{\mathrm{ref}})$, the data split at the event times -- and compared against the constant-coefficient model by likelihood ratio. $\theta$ is the change in log hazard ratio per unit of log time, $\beta$ the log hazard ratio at the median event time $t_{\mathrm{ref}}$, and $\theta = 0$ is proportional hazards. The equations are in the Method section at the end.

| term | beta | theta | theta_ci_low | theta_ci_high | lr_chi2 | p | ref_months | rows | reason |
|---|---|---|---|---|---|---|---|---|---|
| Age (per year) | 0.0242 | -0.0188 | -0.0277 | -0.0100 | 17.8381 | <0.0001 | 12.7890 | 113501 | |
| MGMT: Intermediate | -0.2181 | -0.4103 | -0.7062 | -0.1145 | 6.9901 | 0.0082 | 12.7890 | 113501 | |

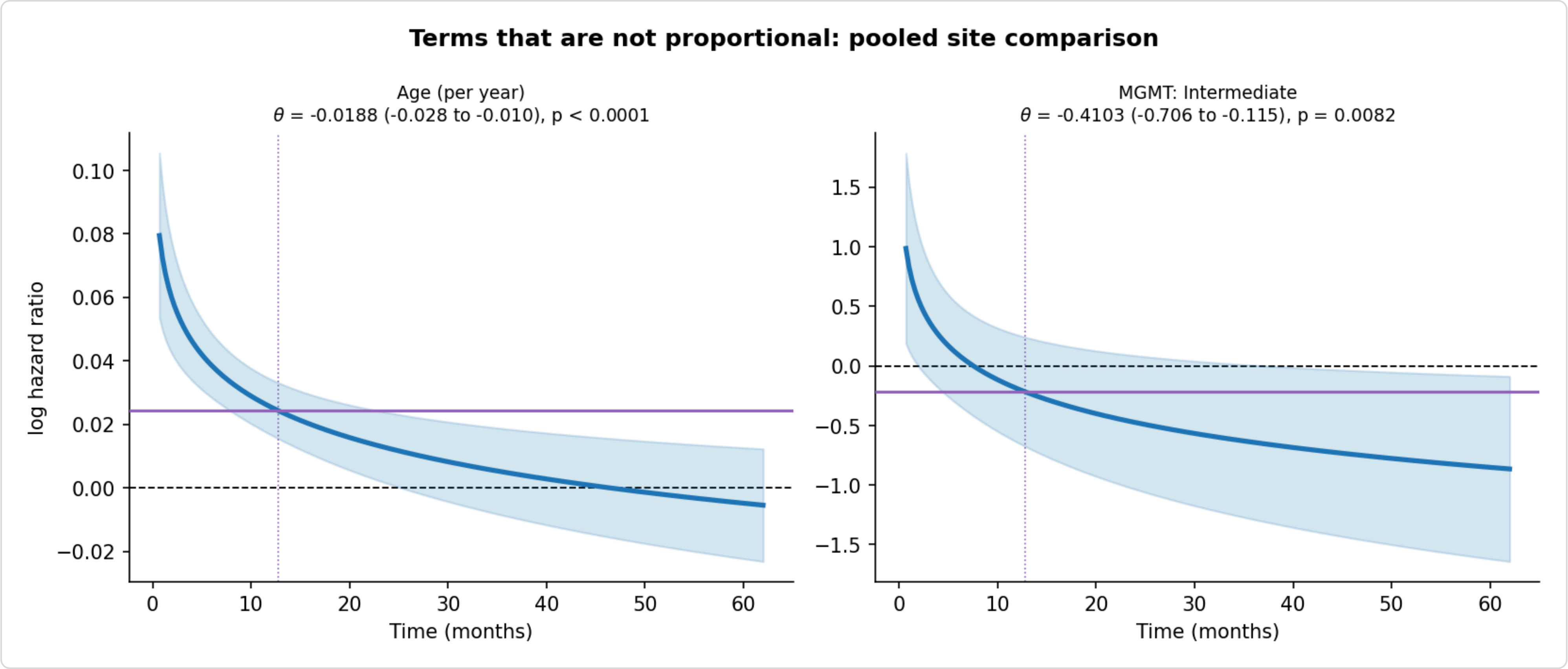


Site-diagnostics_non-proportional-terms

#### 5. Site effect: the coefficient applied to the survival times

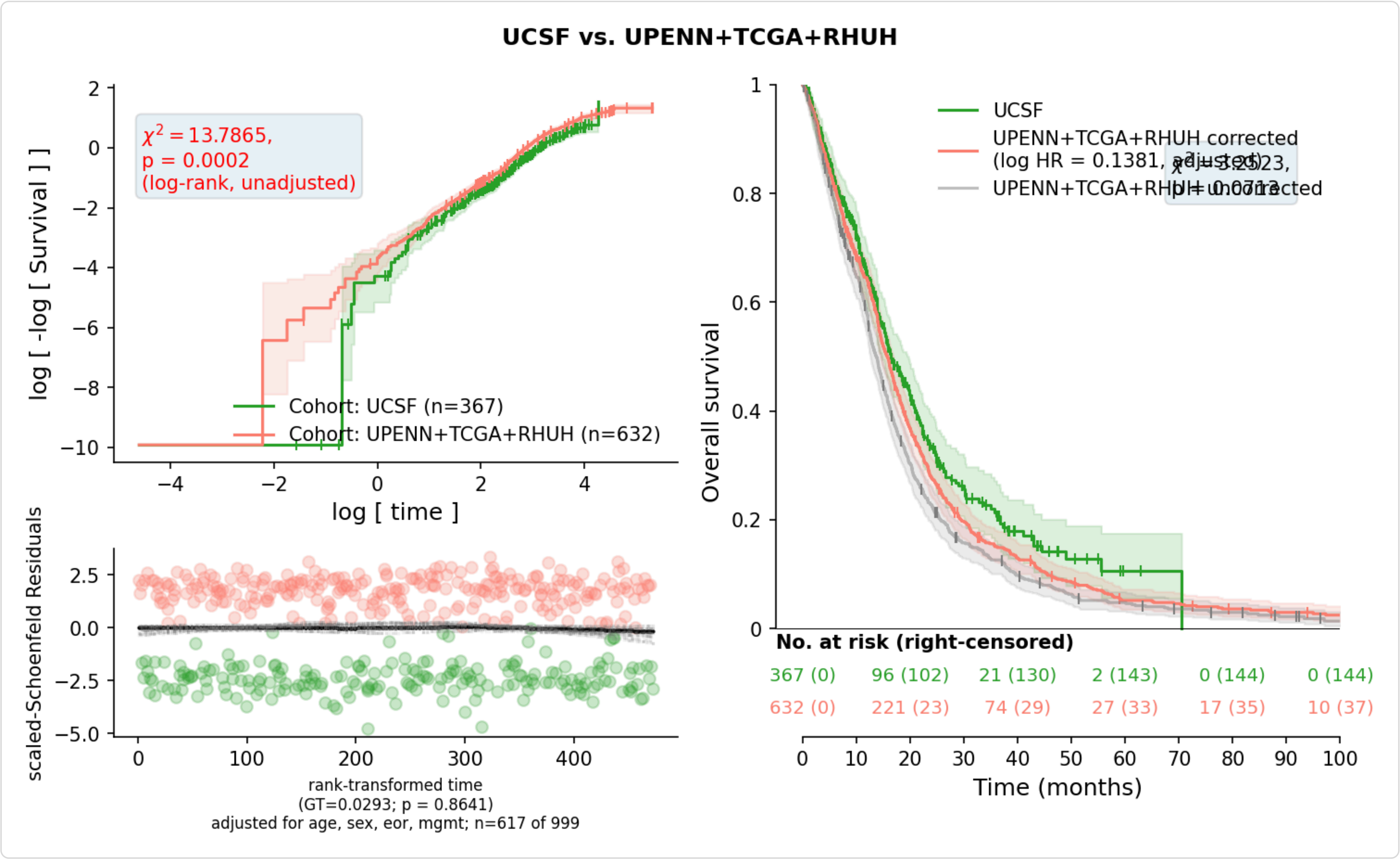


Site-effects_Survival-times_UCSF-UPENN+TCGA+RHUH_adjusted

#### 6. Survival after correction

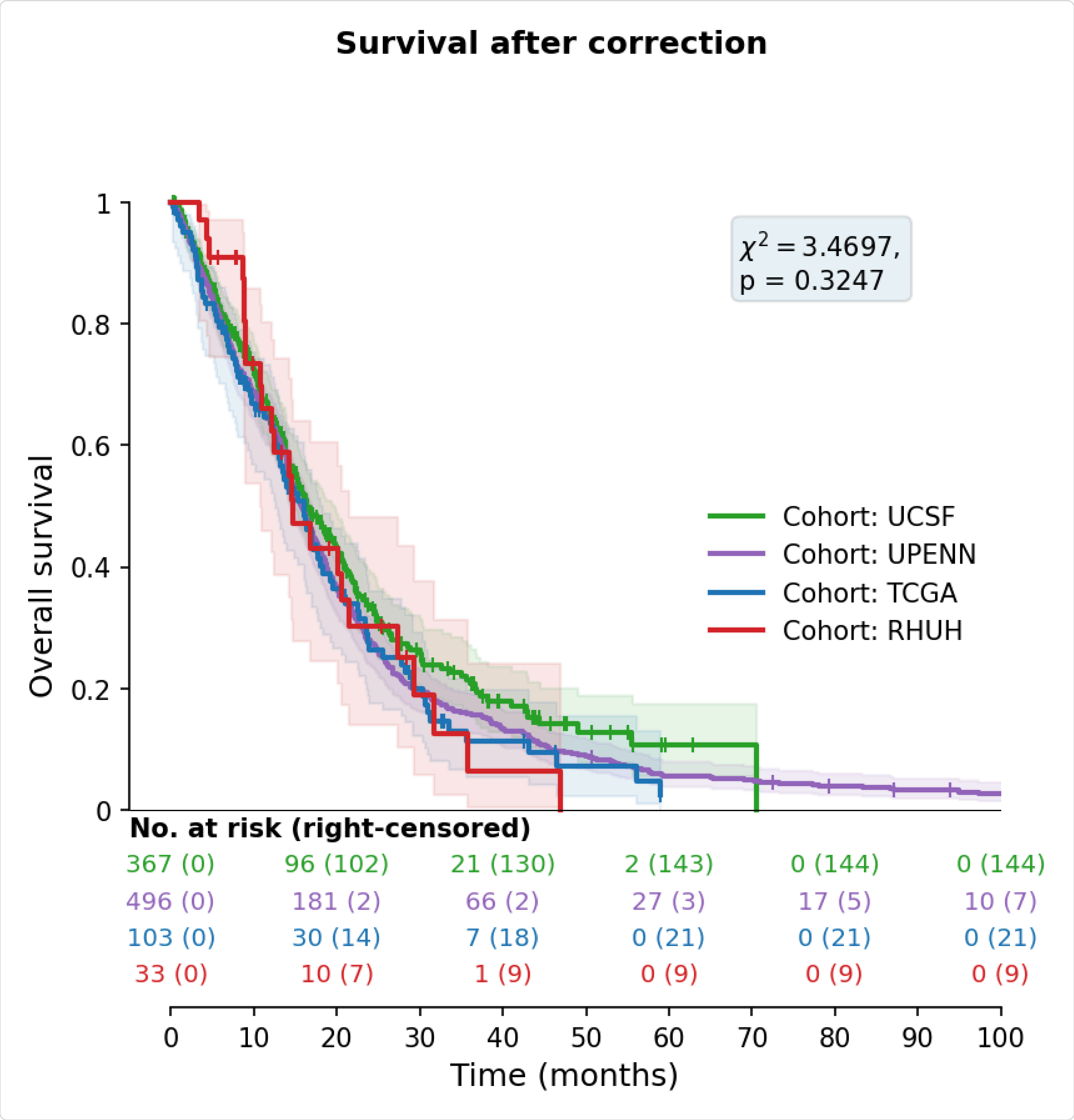


OS-cohorts_after-correction

#### 7. Recommendation: raw or corrected survival times?

> Use the RAW survival times, and adjust or stratify for cohort downstream: once case-mix is held fixed, the site effect is no longer distinguishable from zero. Downstream analyses should read 'OS (days)'.

A site coefficient is worth dividing out of the outcome only if it survives the case-mix that could have produced it. That is what the adjustment ladder above tests, on every run, whatever coefficient was applied -- so this section can disagree with the column the table actually carries, and says so when it does.

##### 7.1 What this run found

Crude site effect, UCSF (site 0) vs UPENN+TCGA+RHUH (site 1): log HR 0.2922 (HR 1.339, p = 0.0002) on all 999 subjects.

Adjusted site effect (age + sex + eor + mgmt): log HR 0.1381 (95% CI -0.0594 to 0.3356, p = 0.1706), fitted on 617 subjects with 472 events.

Case-mix accounts for 54.6% of the site effect measured on the same complete-case sample.

The adjusted confidence interval covers 0 at alpha = 0.05, so what the crude comparison showed is accounted for by who the patients are, and what remains is too small to attribute to entry point or to censoring. Rescaling the outcome by it would remove prognostic signal that belongs to the covariates, and a downstream model that also adjusts for them would remove it twice.

##### 7.2 What qualifies it

The site term is proportional, but 2 covariate(s) in the adjustment are not (Age (per year), MGMT: Intermediate). The site coefficient is therefore estimated while holding fixed a covariate whose own effect drifts with follow-up, so it is an average over a model that does not hold at every time.

The log-time fits report the drift for each of them.

The adjusted estimate rests on 617 of 999 subjects; 382 were dropped for an incomplete covariate. Missingness here is cohort-structured, not random, so the complete cases over-represent the cohorts that record everything.

The largest imbalance between the groups is MGMT = Methylated (|SMD| 0.66, above 0.1), which is the case-mix the adjustment is working against.

The censoring distributions differ between the groups (log-rank p < 0.0001): they were watched for different lengths of time. That costs precision, not unbiasedness, as long as censoring is independent -- which the next points probe but cannot prove.

By 24 months UCSF observed 81.0% of the person-time it owed and UPENN+TCGA+RHUH 97.3% (formal person-time rate): the loss to follow-up is concentrated in one group.

Censoring in UCSF depends on the covariates (p = 0.0001): Age (per year) (censoring HR 1.034, death HR 1.036), Sex: Female (censoring HR 0.666, death HR 0.774), EOR: Subtotal (<90%) (censoring HR 0.481, death HR 0.323), EOR: Gross total (>=90%) (censoring HR 0.351, death HR 0.164). For each, both hazard ratios lie on the same side of 1: the patients more likely to be lost were also more likely to die, so those lost to follow-up were the sicker ones. The adjusted site model absorbs the part of this that runs through its covariates; the Kaplan-Meier curves, crude and corrected, do not.

If the censored patients of UCSF died at delta times the rate the adjusted model predicts for comparable patients who stayed, the site HR would reach 1 at delta = 1.58. The site effect would be significant at alpha = 0.05 for delta = 0.2 (HR 1.842, p < 0.0001); delta = 0.333 (HR 1.623, p < 0.0001); delta = 0.5 (HR 1.481, p = 0.0001); delta = 0.75 (HR 1.266, p = 0.0165); delta = 5 (HR 0.777, p = 0.0046). That is inside the band taken as plausible (0.5-2, --tipping-plausible), so censoring alone could account for the adjusted site effect, and it cannot be attributed to entry point.

This run still wrote a correction: log HR 0.138076 (rescale-adjusted). The corrected column is therefore present but not recommended for this selection; nothing downstream has to use it.

Whichever column is used, use one of the two remedies and not both: rescaling the outcome and stratifying the baseline hazard by cohort are alternatives, and applying them together corrects the same difference twice.

#### 8. Method, and the choices behind it

##### 8.1 What the correction assumes

A survival gap between two groups of cohorts has three possible sources, and they call for different responses. If the clock starts at a different event in one cohort (entry point), that is an artefact of record-keeping and should be removed. If the cohorts genuinely hold different patients -- more methylated MGMT, more gross-total resections, older patients (case-mix) -- that is a real prognostic difference and must be kept, because removing it by rescaling the outcome destroys the signal the analysis is trying to measure; worse, a downstream model that also adjusts for those covariates would then remove the same effect twice. And if one cohort lost its sicker patients to follow-up (informative censoring), its survival looks better than it was; that is an artefact too, but not one a constant rescaling of the survival times can undo.

The correction can only express the first. So the diagnostics work by elimination: the adjustment ladder takes out case-mix, the censoring diagnostics ask how much of the remainder censoring could produce, and only a remainder that survives both is a candidate for entry point -- one candidate among several, since unrecorded case-mix, differences in treatment after the clock starts, and chance leave the same trace. The sources also overlap. Censoring that depends on the covariates is a case-mix effect as far as the adjusted hazard ratio is concerned, and is absorbed by the adjustment; censoring that depends on something unrecorded is not. Entry point and censoring both act mostly in the first months after the clock starts, so a remainder concentrated early is consistent with either, and these data cannot tell them apart.

The correction itself multiplies group 1's survival times by exp(logHR), leaving the reference group untouched. That is a single constant for everyone in the group, so it is the right shape only if the site log hazard ratio is the same early and late -- which is why proportional hazards is tested rather than assumed. Stratifying the baseline hazard by cohort [1] is the equivalent alternative and needs no rescaling at all; the two are alternatives, and applying both corrects the same difference twice.

##### 8.2 Covariate adjustment and missing data

The adjusted coefficient is estimated on the subjects reporting every chosen covariate and then applied to all of them, so the assembled table never shrinks. Missingness here is severe and cohort-structured rather than random -- EOR is unrecorded for all of TCGA, MGMT for all of RHUH, KPS for all of UCSF and LUMIERE -- so a complete-case table would cost most of the sample. The transfer assumes the site effect is the same in complete and incomplete cases, which is an assumption doing real work; the balance table's missingness columns are the evidence to weigh it against. Imbalance is measured by the standardised mean difference (SMD) [7], which unlike a p-value does not shrink as the sample grows. For a covariate with mean $\bar{x}_g$ and sample variance $s_g^2$ in site group $g$,

$$\mathrm{SMD} = \frac{|\bar{x}_1 - \bar{x}_0|}{\sqrt{(s_0^2 + s_1^2)/2}}, \qquad s_g^2 = \frac{1}{n_g - 1} \sum_{i \in g} (x_i - \bar{x}_g)^2.$$

Each $s_g^2$ is the unbiased sample variance, and the two are averaged unweighted, so the larger group does not dominate the scale and the groups are not assumed to share a variance. A categorical covariate is compared one level at a time on the 0/1 indicator of that level; $\bar{x}_g$ is then the proportion $p_g$ and the same formula gives $s_g^2 = p_g(1-p_g)\,n_g/(n_g-1)$ -- one definition for both kinds of covariate, rather than the plug-in $p_g(1-p_g)$ often used for proportions [7], from which it differs only by $n_g/(n_g-1)$.

##### 8.3 Proportional hazards, and what is done when it fails

Every coefficient in this report comes from a Cox model [1]. For subject $i$ with site indicator $s_i$ and case-mix covariates $x_{i1},\dots,x_{ip}$ (the dummy-coded columns of the adjustment set), the hazard is

$$h_i(t) = h_0(t)\,\exp\Big(\beta_{\text{site}}\,s_i + \sum_{k=1}^{p}\beta_k\,x_{ik}\Big),$$

where $h_0(t)$ is left unspecified. Proportional hazards is the claim that each $\beta$ is a constant, so that the hazard ratio between any two subjects does not depend on time:

$$\frac{h_i(t)}{h_j(t)} = \exp\big(\boldsymbol{\beta}^\top(\mathbf{x}_i-\mathbf{x}_j)\big)\quad\text{for every } t.$$

It is tested for every term of the model the coefficient comes from, not for the site term alone: $\hat{\beta}_{\text{site}}$ is estimated while holding the covariates fixed, so a covariate whose own effect drifts with time makes that adjustment a misspecified one. The alternative each term is tested against lets its coefficient move with a known function of time $g(t)$:

$$\beta_k(t) = \beta_k + \theta_k\,g(t),\qquad H_0:\ \theta_k = 0.$$

The Grambsch-Therneau test [4, 5] is the score test for $\theta_k$, and needs only the constant-coefficient fit. Order the $D$ event times $t_1 \le \cdots \le t_D$, let $i_j$ be the subject failing at $t_j$ and $R(t_j)$ everyone still at risk then. The Schoenfeld residual is the failing subject's covariates minus their risk-weighted average over the risk set, and scaling it by the inverse information turns it into a noisy reading of the coefficient at that moment:

$$\mathbf{r}_j = \mathbf{x}_{i_j} - \frac{\sum_{l\in R(t_j)}\mathbf{x}_l\,e^{\hat{\boldsymbol{\beta}}^\top\mathbf{x}_l}}{\sum_{l\in R(t_j)}e^{\hat{\boldsymbol{\beta}}^\top\mathbf{x}_l}},\qquad \mathbf{s}_j^* = D\,\widehat{\mathrm{Var}}(\hat{\boldsymbol{\beta}})\,\mathbf{r}_j,\qquad \mathbb{E}\big[s_{kj}^*\big] + \hat{\beta}_k \approx \beta_k(t_j).$$

A time trend in $\beta_k$ is therefore a correlation between $s_{kj}^*$ and $g(t_j)$, and the statistic measures it:

$$T_k = \frac{\Big[\sum_{j=1}^{D}(g_j-\bar{g})\,s_{kj}^*\Big]^2}{D\,\widehat{\mathrm{Var}}(\hat{\beta}_k)\,\sum_{j=1}^{D}(g_j-\bar{g})^2}\ \sim\ \chi_1^2\text{ under }H_0,\qquad g_j = j.$$

$g_j = j$ is the rank transform, the lifelines [9] default; R's cox.zph defaults to the Kaplan-Meier transform instead. With $K$ terms screened in one pass, the p-value that decides which terms are followed up is the Bonferroni-adjusted one, not the raw one; the raw p is reported beside it so the more sensitive reading stays visible:

$$p_k^{\text{Bonf}} = \min\big(1,\ K\,p_k\big),\qquad \text{term } k \text{ is followed up if } p_k^{\text{Bonf}} < 0.05.$$

A term that fails is then described rather than merely flagged. It is refitted with a log-time interaction, one model per failing term, the other terms keeping constant coefficients:

$$h_i(t) = h_0(t)\,\exp\Big(\sum_l \beta_l\,x_{il} + \theta_k\,x_{ik}\,\log\frac{t}{t_{\text{ref}}}\Big)\quad\Longleftrightarrow\quad \beta_k(t) = \beta_k + \theta_k\log\frac{t}{t_{\text{ref}}},$$

with $t_{\text{ref}}$ the median event time, so $\beta_k$ is the log hazard ratio at that moment rather than at $t = 1$ day. Equivalently $\mathrm{HR}_k(t) = e^{\beta_k}\,(t/t_{\text{ref}})^{\theta_k}$: the hazard ratio is multiplied by $2^{\theta_k}$ each time follow-up doubles, and $\theta_k = 0$ is proportional hazards. Because the covariate $x_{ik}\log(t/t_{\text{ref}})$ changes with time, the data are split at the event times $t_{(1)} < \cdots < t_{(D)}$ into intervals $\big(t_{(m-1)}, t_{(m)}\big]$, and the model is fitted by the partial likelihood (written here without the tie correction)

$$\ell(\boldsymbol{\beta},\theta_k) = \sum_{m=1}^{D}\Big[\eta_{i_m}(\tau_m) - \log\sum_{l\in R(t_{(m)})}e^{\eta_l(\tau_m)}\Big],\qquad \eta_l(\tau) = \sum_q \beta_q x_{lq} + \theta_k\,x_{lk}\log\frac{\tau}{t_{\text{ref}}},\qquad \tau_m = \max\big(t_{(m-1)},\ 1\text{ day}\big).$$

Three details in that expression decide whether the fit is an estimate or an artefact, and all three are deliberate. First, the split is at the event times rather than on a fixed grid, so the whole risk set $R(t_{(m)})$ shares one interval and one value of log time; a monthly grid attenuates $\theta_k$ by roughly a third. Second, the interaction is evaluated at $\tau_m$, the interval's start, the value every member of the risk set shares -- evaluating it at the interval end hands the subject who fails a systematically smaller time than the controls, which on simulated data with no time trend at all rejects proportional hazards in twelve of twelve replicates. Third, there is no main effect of time: a term $\gamma \log \tau_m$ is the same for every subject in the sum, so $e^{\gamma \log \tau_m}$ cancels between numerator and denominator, $\gamma$ never enters $\ell$, and the fit cannot converge. When a pool has more than 1000 distinct event times the cuts are taken at 1000 quantiles instead, which shrinks $\theta_k$ slightly toward zero.

$\theta_k$ is tested by likelihood ratio against the constant-coefficient fit $\tilde{\boldsymbol{\beta}}$ on the same split data, and the band drawn around $\hat{\beta}_k(t)$ is the pointwise 95% interval from the joint covariance of $(\hat{\beta}_k, \hat{\theta}_k)$:

$$\Lambda_k = 2\big[\ell(\hat{\boldsymbol{\beta}}, \hat{\theta}_k) - \ell(\tilde{\boldsymbol{\beta}}, 0)\big] \;\sim\; \chi^2_1,$$

$$\hat{\beta}_k(t) \pm 1.96\sqrt{\widehat{\mathrm{Var}}(\hat{\beta}_k) + u^2\,\widehat{\mathrm{Var}}(\hat{\theta}_k) + 2u\,\widehat{\mathrm{Cov}}(\hat{\beta}_k, \hat{\theta}_k)}, \qquad u = \log\frac{t}{t_{\mathrm{ref}}}.$$

The band is narrowest at $t_{\mathrm{ref}}$, where $u = 0$, and widens in both directions; it is pointwise, not a simultaneous band over the whole curve.

##### 8.4 Censoring: what is measured and what is only stressed

Under independent censoring the Cox partial likelihood is consistent however differently the two groups were censored, so differential follow-up costs precision rather than unbiasedness. Three diagnostics probe the assumption. The reverse Kaplan-Meier [6] says how long each group was watched; the person-time rates [11, 12] say how completely, without counting a death as a loss. The censoring-hazard models say whether censoring depends on the recorded covariates: if it does, the adjusted site model is still consistent, because censoring is independent given what it conditions on, but no marginal Kaplan-Meier curve is. The tipping point [13] then asks how far censoring would have to depart from independence to move the adjusted site effect. For each censored subject $i$ of the shifted group, with censoring time $c_i$ and linear predictor $\eta_i$ from the adjusted model, a death time is drawn from

$$H_0(T_i^*) = H_0(c_i) + \frac{E_i}{\delta\, e^{\eta_i}}, \qquad E_i \sim \mathrm{Exp}(1),$$

with $H_0$ the Breslow baseline recomputed under coefficients redrawn from their sampling distribution for every imputation, and the site coefficients of the refitted models pooled by Rubin's rules [14]. $\delta = 1$ is independent censoring; $\delta = 2$ says the patients lost to follow-up died twice as fast as comparable patients who stayed. The shift is applied to each group in turn, and to every censored subject of it -- administrative censorings included, since the tables do not say which censorings were administrative.

##### 8.5 What cannot be checked here

Whether censoring depends on something nobody recorded. UCSF records no KPS at all, and performance status is the obvious reason a patient stops returning to the centre that diagnosed them. The tipping point bounds how much that could matter; it cannot say which delta is true. If censoring is informative beyond the covariates, the bias sits inside every estimate above.

##### 8.6 Estimation details

The baseline hazard is Breslow's estimator [2] throughout, requested explicitly. Ties in the partial likelihood are a separate matter: lifelines [9] implements Efron's approximation [3] only and offers no way to change it, so every diagnostic fit here breaks ties by Efron, while the coefficient actually applied to the survival times comes from scikit-survival [10], which uses Breslow for both. With many tied survival days the two differ in the second or third decimal. Survival curves are Kaplan-Meier estimates [8] with log-log confidence bands.

#### 9. Provenance

How the correction was obtained, as written to data-clinical_TD-tissues_4-cohorts_site-correction.json next to the table.


```
{
    "adjusted_for": [
        "age",
        "sex",
        "eor",
        "mgmt"
    ],
    "design_columns": [
        "Age (per year)",
        "Sex: Female",
        "EOR: Subtotal (<90%)",
        "EOR: Gross total (>=90%)",
        "MGMT: Intermediate",
        "MGMT: Methylated"
    ],
    "missing_strategy": "complete-case",
    "logHR": 0.13807578399151754,
    "logHR_crude": 0.29214548678235003,
    "n_model": 617,
    "events_model": 472,
    "n_applied": 999,
    "ph_test_p": 0.8832334625187894,
    "notes": [
        "Sex: reference level is Male.",
        "EOR: reference level is Biopsy.",
        "MGMT: reference level is Unmethylated.",
        "382 subject(s) dropped from the model for an incomplete covariate (the coefficient is still applied to all 999)."
    ],
    "mode": "rescale-adjusted",
    "HR": 1.1480625517401606,
    "se": 0.10075552160087767,
    "ci95_logHR": [
        -0.059401409589750126,
        0.33555297757278524
    ],
    "ci95_HR": [
        0.9423284335584865,
        1.3987136286770263
    ],
    "p": 0.17056078027771748,
    "applied_as": "OS (days) - corrected = OS (days) * exp(logHR * site)",
    "script": "createDatabase.py",
    "timestamp": "2026-10-01 09:40:55",
    "command": "NormWM_4_Glioblastoma/createDatabase.py /home/joan/Desktop/PROJECTS/Glioblastomas RESULTS-GBM_4-cohorts_Tissue
    "cohorts": [
        "UCSF",
        "UPENN",
        "TCGA",
        "RHUH"
    ],
    "cohort_ids": {
        "UCSF": 0,
        "UPENN": 1,
        "TCGA": 2,
        "RHUH": 3
    },
    "site_codes": {
        "UCSF": 0,
        "UPENN": 1,
        "TCGA": 1,
        "RHUH": 1
    },
    "site_labels": {
        "0": "UCSF",
        "1": "UPENN+TCGA+RHUH"
    },
    "site_reference": [
        "UCSF"
    ],
    "site_partition": "default",
    "output": "data-clinical_TD-tissues_4-cohorts.csv",
```

"recommended_outcome": "raw"
}


#### 10. Run log

Everything this run printed -- sample sizes, censoring, every fit and every warning, in the order they happened -- is in the text file below, not in this page. It is the same content the sections above are drawn from, at the level of detail a log has rather than a report.

Run log

```
/home/joan/Desktop/PROJECTS/Glioblastomas/RESULTS-GBM_4-cohorts_Tissues/createDatabase_log.txt
```
