## Supplementary material B and C for "Beyond tumor volume: connectomic measures of tumor burden provide superior prognostic information across tumor compartments in glioblastoma": Falco_Roget_et_al_Suppl-C1.pdf

### L-TDI or tumour volume: Cox models of survival

```
LTDI-Volume_comparison.py /home/joan/Desktop/PROJECTS/Glioblastomas RESULTS-GBM_4-cohorts_Tissues --data data-clinical_TD-tissues_4-cohorts.csv --duration-col OS (days) --event-col status --stratify-for none --seed 42 --format both --log
```

#### 1. Run settings

| Setting | Value |
|---|---|
| Data | data-clinical_TD-tissues_4-cohorts.csv (995 patients with every marker) |
| Survival column | OS (days) |
| Event column | status |
| Stratification (in-sample Cox) | none |
| Standardization | off — HRs in native units: per year, per cm³, per L-TDI unit, per category level |
| Cross-validation | same survival column, never stratified |
| Bootstrap resamples / seed | 5000 / 42 |
| File tag | strata-none_standardized-false |

Terms. HR: hazard ratio, exp(β), the multiplicative change in the hazard per unit of the covariate (above 1 means shorter survival). 95% CI: Wald confidence interval. C-index: Harrell's concordance, the fraction of comparable patient pairs whose predicted risks order their survival correctly (0.5 is chance), pooled across strata; its interval is a percentile bootstrap over patients with the fitted model held fixed. AIC: Akaike information criterion of the partial likelihood, 2k – 2 log L, lower is better. LLR: likelihood-ratio test of a model against the base it extends by one covariate ($\chi^2$, 1 degree of freedom).

#### 2. L-TDI or volume, head to head

Two markers enter one Cox model together, without clinical covariates, so each hazard ratio is that marker's association with survival holding the other fixed. A marker whose interval still excludes 1 carries information the other lacks.

Hazard ratios of the head-to-head models.

| Model | Covariate | HR (95% CI) | p | N | C-index |
|---|---|---:|---:|---:|---:|
| E.size + W.L-TDI | E.size (per cm³) | 1.0015 (0.9970–1.0061) | 0.5070 | 995 | 0.567 |
| E.size + W.L-TDI | W.L-TDI | 1.0953 (1.0423–1.1510) | 0.0003 | 995 | 0.567 |
| E.size + E.L-TDI | E.size (per cm³) | 1.0062 (1.0026–1.0098) | 0.0007 | 995 | 0.575 |
| E.size + E.L-TDI | E.L-TDI | 1.0031 (1.0018–1.0043) | <0.0001 | 995 | 0.575 |
| E.L-TDI + W.L-TDI | E.L-TDI | 1.0025 (1.0013–1.0038) | <0.0001 | 995 | 0.581 |
| E.L-TDI + W.L-TDI | W.L-TDI | 1.0857 (1.0420–1.1314) | <0.0001 | 995 | 0.581 |

#### 3. Pre-surgical models

The base model is Age + Sex; each marker model adds one marker to it. ΔAIC and the likelihood-ratio p-value compare each marker model with the base (same patients), and the AIC column ranks the three markers against one another.

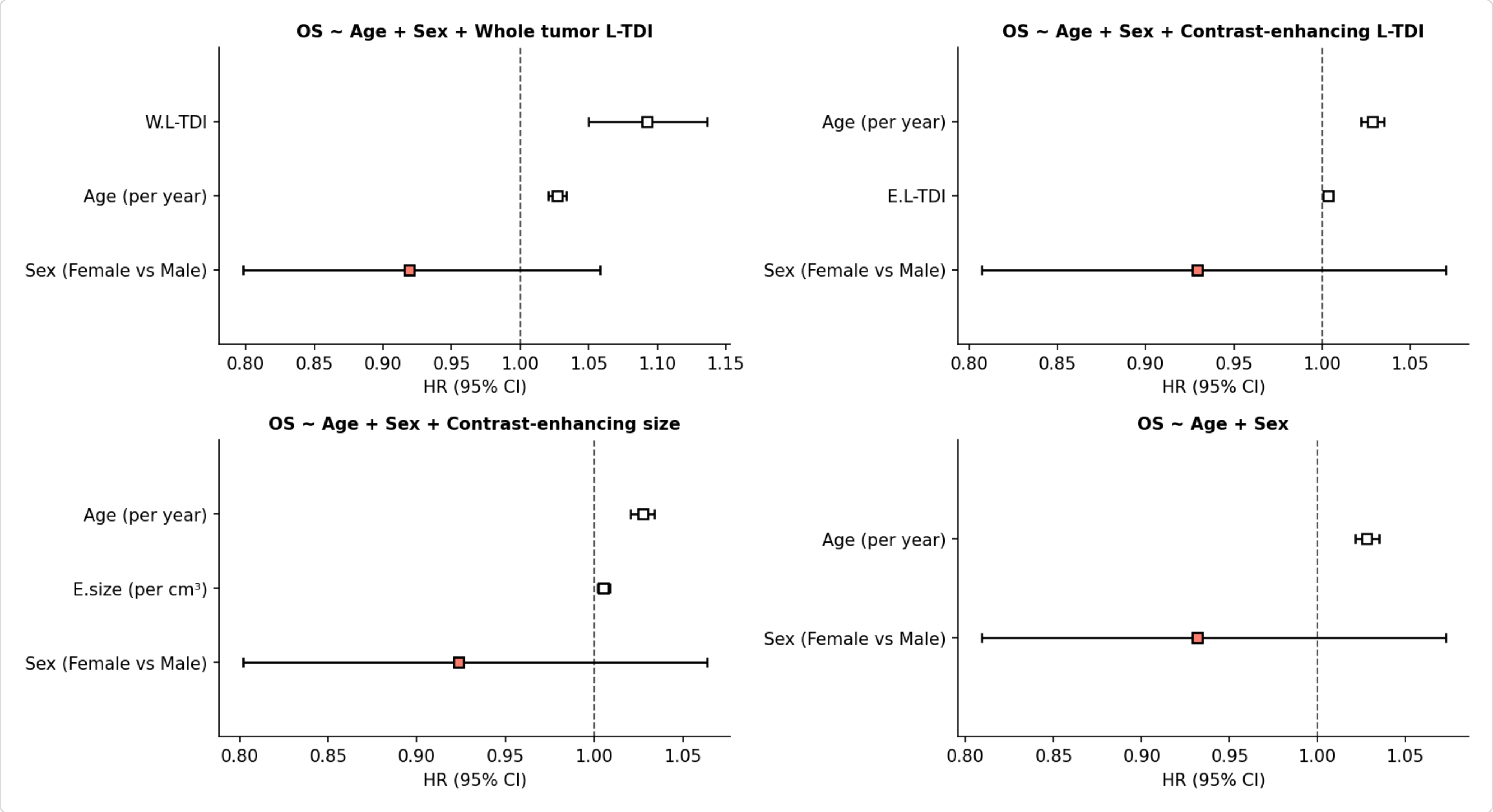


Hazard ratios with 95% CI. A salmon square marks a covariate whose interval crosses HR = 1; a white one, an interval that excludes it.

Fit of the pre-surgical models.

| Model | N | Events | C-index (95% CI) | Log-likelihood | AIC | ΔAIC vs base | LLR p vs base |
|---|---|---|---|---|---|---|---|
| W.L-TDI | 995 | 807 | 0.631 (0.610–0.652) | -4763.8557 | 9533.7114 | -17.2542 | <0.0001 |
| E.L-TDI | 995 | 807 | 0.630 (0.609–0.652) | -4760.3404 | 9526.6808 | -24.2848 | <0.0001 |
| E.size | 995 | 807 | 0.622 (0.601–0.644) | -4769.2150 | 9544.4299 | -6.5357 | 0.0035 |
| Base | 995 | 807 | 0.618 (0.596–0.640) | -4773.4828 | 9550.9656 | | |

#### 4. Post-surgical models

The base model is Age + Sex + MGMT + EOR; each marker model adds one marker to it. ΔAIC and the likelihood-ratio p-value compare each marker model with the base (same patients), and the AIC column ranks the three markers against one another.

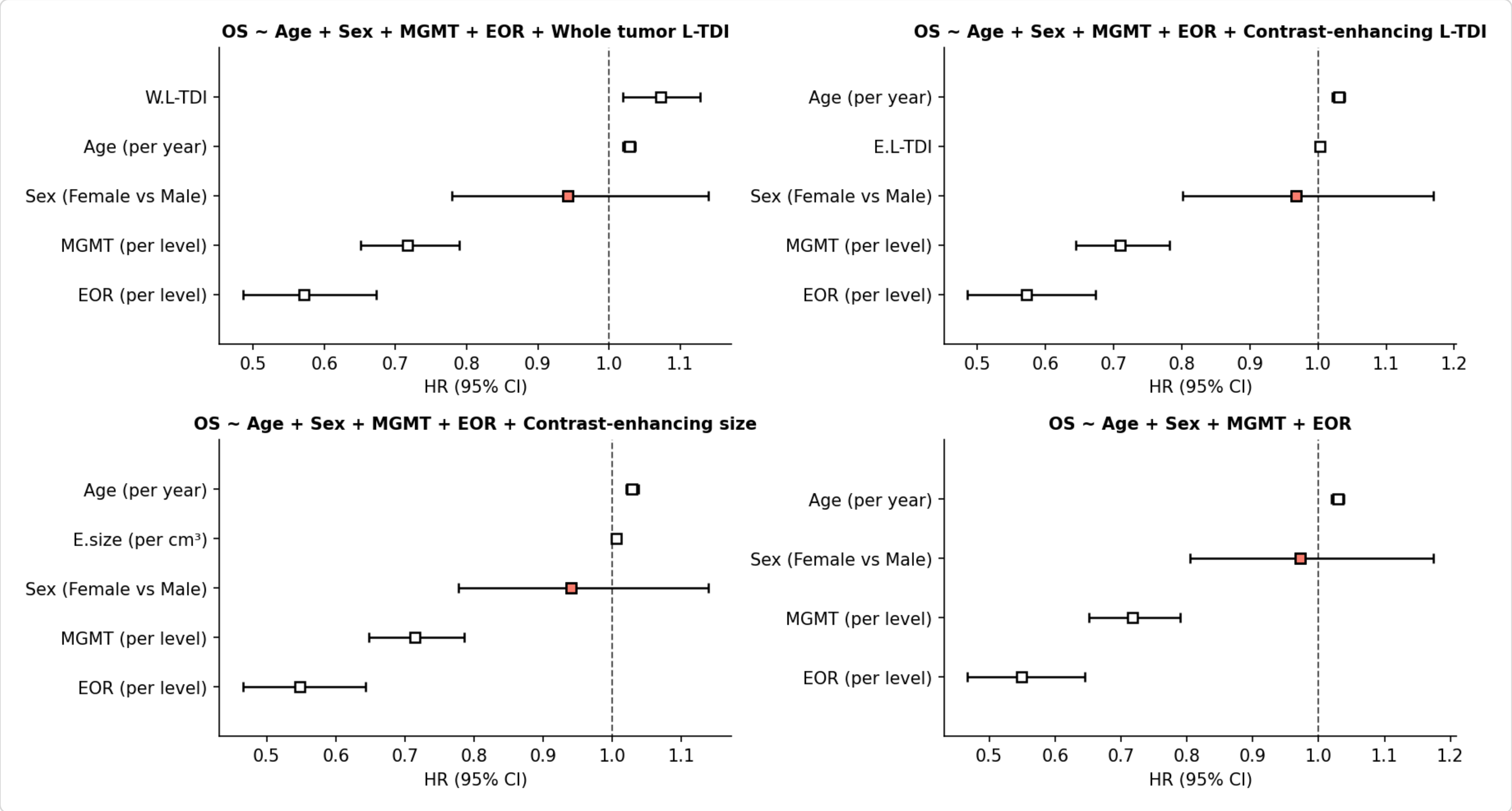

Hazard ratios with 95% CI. A salmon square marks a covariate whose interval crosses HR = 1; a white one, an interval that excludes it.

Fit of the post-surgical models.

| Model | N | Events | C-index (95% CI) | Log-likelihood | AIC | ΔAIC vs base | LLR p vs base |
|---|---|---|---|---|---|---|---|
| W.L-TDI | 615 | 470 | 0.678 (0.651–0.704) | -2507.5885 | 5025.1771 | -5.4321 | 0.0064 |
| E.L-TDI | 615 | 470 | 0.677 (0.649–0.702) | -2505.7371 | 5021.4742 | -9.1350 | 0.0008 |
| E.size | 615 | 470 | 0.674 (0.647–0.700) | -2508.9116 | 5027.8232 | -2.7860 | 0.0287 |
| Base | 615 | 470 | 0.672 (0.644–0.699) | -2511.3046 | 5030.6092 | | |

#### 5. Leave-one-cohort-out validation

Each model is fitted on all cohorts but one and scored on the one left out, analysing 'OS (days)' without strata. Cells are held-out C-indices. Mean, SD and Min summarise them across the held-out cohorts: SD is the sample standard deviation (n – 1), how much discrimination depends on which cohort is left out, and Min is the worst held-out cohort. RMSE is the root mean squared error of the predicted expected survival on held-out patients whose death was observed, averaged over the held-out cohorts.

Held-out C-index by cohort left out.

| Covariates | Fitter | UCSF | UPENN | TCGA | RHUH | Mean | SD | Min | RMSE (days) |
|---|---|---|---|---|---|---|---|---|---|
| Age + Sex + W.L-TDI | Cox | 0.646 | 0.614 | 0.649 | 0.641 | 0.637 | 0.016 | 0.614 | 451.0 |
| Age + Sex + W.L-TDI | Weibull AFT | 0.646 | 0.613 | 0.647 | 0.644 | 0.637 | 0.016 | 0.613 | 402.9 |
| Age + Sex + W.L-TDI | LogLogistic AFT | 0.644 | 0.614 | 0.648 | 0.644 | 0.638 | 0.016 | 0.614 | 518.3 |
| Age + Sex + E.L-TDI | Cox | 0.620 | 0.616 | 0.631 | 0.610 | 0.619 | 0.009 | 0.610 | 472.2 |
| Age + Sex + E.L-TDI | Weibull AFT | 0.620 | 0.615 | 0.628 | 0.613 | 0.619 | 0.007 | 0.613 | 420.8 |
| Age + Sex + E.L-TDI | LogLogistic AFT | 0.627 | 0.617 | 0.631 | 0.630 | 0.626 | 0.006 | 0.617 | 537.4 |
| Age + Sex + E.size | Cox | 0.631 | 0.606 | 0.636 | 0.674 | 0.637 | 0.028 | 0.606 | 433.1 |
| Age + Sex + E.size | Weibull AFT | 0.629 | 0.606 | 0.639 | 0.677 | 0.637 | 0.030 | 0.606 | 390.6 |
| Age + Sex + E.size | LogLogistic AFT | 0.632 | 0.606 | 0.634 | 0.666 | 0.634 | 0.025 | 0.606 | 513.6 |
| Age + Sex | Cox | 0.635 | 0.602 | 0.626 | 0.623 | 0.621 | 0.014 | 0.602 | 455.1 |
| Age + Sex | Weibull AFT | 0.635 | 0.602 | 0.626 | 0.634 | 0.624 | 0.015 | 0.602 | 406.2 |
| Age + Sex | LogLogistic AFT | 0.632 | 0.602 | 0.626 | 0.623 | 0.621 | 0.013 | 0.602 | 533.5 |

#### 6. Files

Full-precision tables: LTDI-Volume_comparison_strata-none_standardized-false_coefficients.csv, LTDI-Volume_comparison_strata-none_standardized-false_model-fit.csv, LTDI-Volume_comparison_strata-none_standardized-false_cross-validation.csv. Run log: LTDI-Volume_comparison_strata-none_standardized-false_log.txt. Per-model forest plots: <PreIPost>surgical_<marker>_strata-none_standardized-false.pdf/svg. All in /home/joan/Desktop/PROJECTS/Glioblastomas/RESULTS-GBM_4-cohorts_Tissues/Forest-plots_Cox-models.
