## Supplementary material B and C for "Beyond tumor volume: connectomic measures of tumor burden provide superior prognostic information across tumor compartments in glioblastoma": Falco_Roget_et_al_Suppl-C2.pdf

### L-TDI or tumour volume: Cox models of survival

```
LTDI-Volume_comparison.py /home/joan/Desktop/PROJECTS/Glioblastomas RESULTS-GBM_4-cohorts_Tissues --data data-clinical_TD-tissues_4-cohorts.csv --duration-col OS (days) --event-col status --stratify-for none --standardize --seed 42 --format both --log
```

#### 1. Run settings

| Setting | Value |
|---|---|
| Data | data-clinical_TD-tissues_4-cohorts.csv (995 patients with every marker) |
| Survival column | OS (days) |
| Event column | status |
| Stratification (in-sample Cox) | none |
| Standardization | on — HRs in continuous covariates per SD; categorical codes on [-1, 1] (sex ±1, MGMT and EOR -1/0/1), so a binary covaria |
| Cross-validation | same survival column, never stratified |
| Bootstrap resamples / seed | 5000 / 42 |
| File tag | strata-none_standardized-true |

Terms. HR: hazard ratio, exp(β), the multiplicative change in the hazard per unit of the covariate (above 1 means shorter survival). 95% CI: Wald confidence interval. C-index: Harrell's concordance, the fraction of comparable patient pairs whose predicted risks order their survival correctly (0.5 is chance), pooled across strata; its interval is a percentile bootstrap over patients with the fitted model held fixed. AIC: Akaike information criterion of the partial likelihood, $2k - 2 \log L$, lower is better. LLR: likelihood-ratio test of a model against the base it extends by one covariate ($\chi^2$, 1 degree of freedom).

#### 2. L-TDI or volume, head to head

Two markers enter one Cox model together, without clinical covariates, so each hazard ratio is that marker's association with survival holding the other fixed. A marker whose interval still excludes 1 carries information the other lacks.

Hazard ratios of the head-to-head models.

| Model | Covariate | HR (95% CI) | p | N | C-index |
|---|---|---:|---:|---:|---:|
| E.size + W.L-TDI | E.size (per SD) | 1.0316 (0.9410–1.1310) | 0.5070 | 995 | 0.567 |
| E.size + W.L-TDI | W.L-TDI (per SD) | 1.1785 (1.0775–1.2888) | 0.0003 | 995 | 0.567 |
| E.size + E.L-TDI | E.size (per SD) | 1.1324 (1.0540–1.2165) | 0.0007 | 995 | 0.575 |
| E.size + E.L-TDI | E.L-TDI (per SD) | 1.1910 (1.1106–1.2773) | <0.0001 | 995 | 0.575 |
| E.L-TDI + W.L-TDI | E.L-TDI (per SD) | 1.1545 (1.0747–1.2402) | <0.0001 | 995 | 0.581 |
| E.L-TDI + W.L-TDI | W.L-TDI (per SD) | 1.1600 (1.0770–1.2494) | <0.0001 | 995 | 0.581 |

#### 3. Pre-surgical models

The base model is Age + Sex; each marker model adds one marker to it. ΔAIC and the likelihood-ratio p-value compare each marker model with the base (same patients), and the AIC column ranks the three markers against one another.

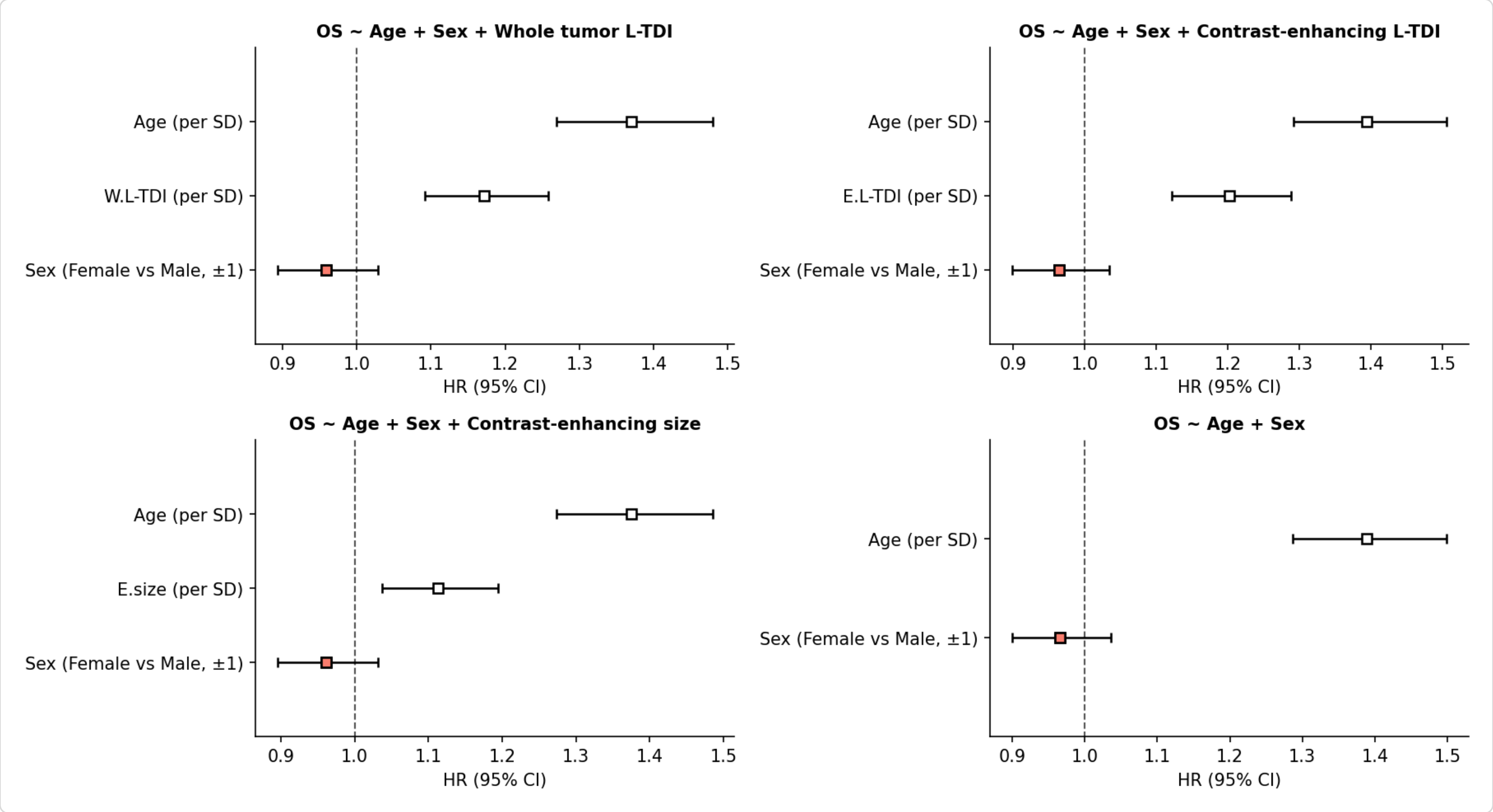


Hazard ratios with 95% CI. A salmon square marks a covariate whose interval crosses HR = 1; a white one, an interval that excludes it.

Fit of the pre-surgical models.

| Model | N | Events | C-index (95% CI) | Log-likelihood | AIC | ΔAIC vs base | LLR p vs base |
|---|---|---|---|---|---|---|---|
| W.L-TDI | 995 | 807 | 0.631 (0.610–0.652) | -4763.8557 | 9533.7114 | -17.2542 | <0.0001 |
| E.L-TDI | 995 | 807 | 0.630 (0.609–0.652) | -4760.3404 | 9526.6808 | -24.2848 | <0.0001 |
| E.size | 995 | 807 | 0.622 (0.601–0.644) | -4769.2150 | 9544.4299 | -6.5357 | 0.0035 |
| Base | 995 | 807 | 0.618 (0.596–0.640) | -4773.4828 | 9550.9656 | | |

#### 4. Post-surgical models

The base model is Age + Sex + MGMT + EOR; each marker model adds one marker to it. ΔAIC and the likelihood-ratio p-value compare each marker model with the base (same patients), and the AIC column ranks the three markers against one another.

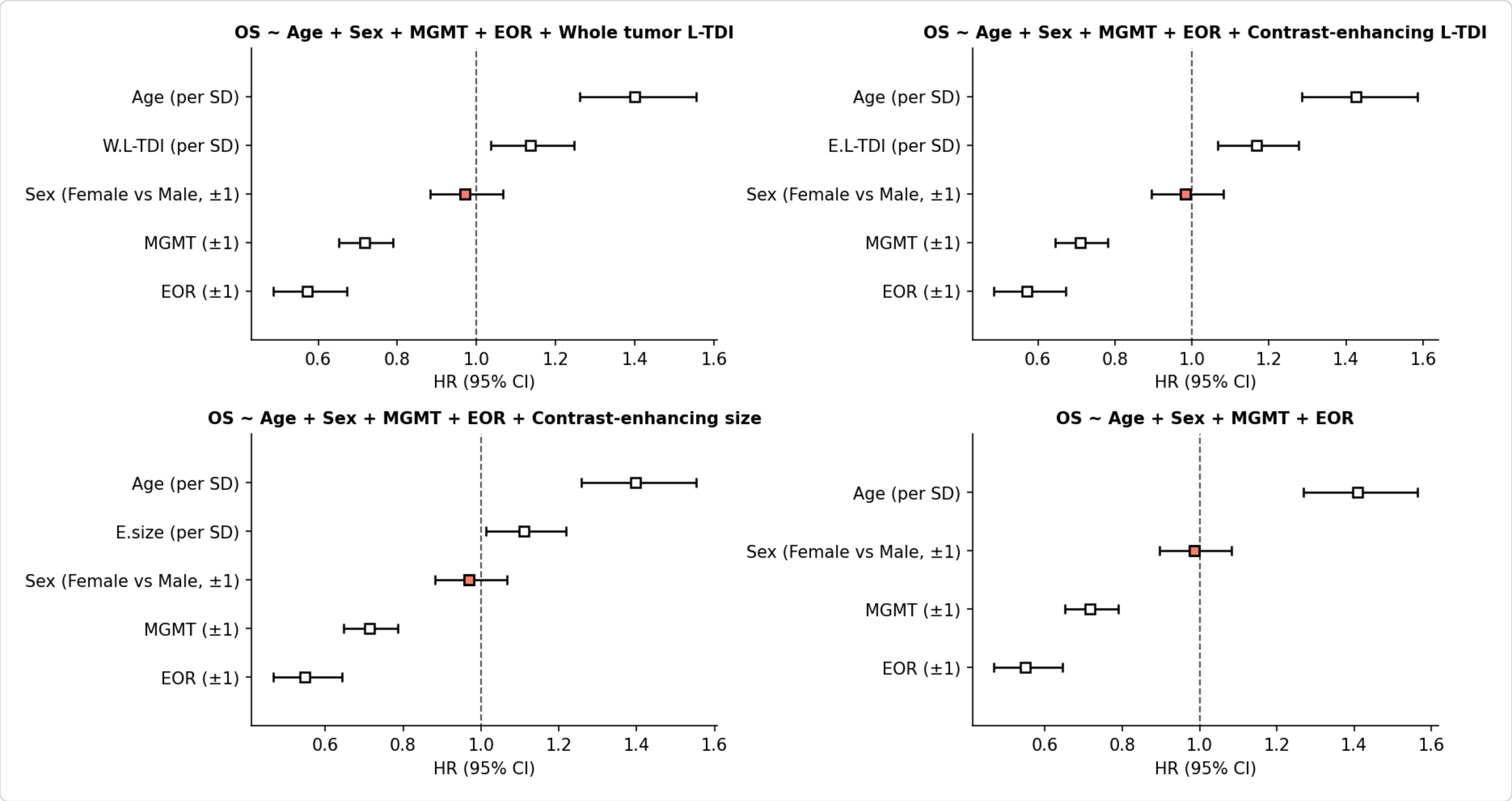

Hazard ratios with 95% CI. A salmon square marks a covariate whose interval crosses HR = 1; a white one, an interval that excludes it.

Fit of the post-surgical models.

| Model | N | Events | C-index (95% CI) | Log-likelihood | AIC | ΔAIC vs base | LLR p vs base |
|---|---|---|---|---|---|---|---|
| W.L-TDI | 615 | 470 | 0.678 (0.651–0.704) | -2507.5885 | 5025.1771 | -5.4321 | 0.0064 |
| E.L-TDI | 615 | 470 | 0.677 (0.649–0.702) | -2505.7371 | 5021.4742 | -9.1350 | 0.0008 |
| E.size | 615 | 470 | 0.674 (0.647–0.700) | -2508.9116 | 5027.8232 | -2.7860 | 0.0287 |
| Base | 615 | 470 | 0.672 (0.644–0.699) | -2511.3046 | 5030.6092 | | |

#### 5. Leave-one-cohort-out validation

Each model is fitted on all cohorts but one and scored on the one left out, analysing 'OS (days)' without strata. Standardization is estimated on the training cohorts and applied unchanged to the held-out cohort. Cells are held-out C-indices. Mean, SD and Min summarise them across the held-out cohorts: SD is the sample standard deviation (n − 1), how much discrimination depends on which cohort is left out, and Min is the worst held-out cohort. RMSE is the root mean squared error of the predicted expected survival on held-out patients whose death was observed, averaged over the held-out cohorts.

Held-out C-index by cohort left out.

| Covariates | Fitter | UCSF | UPENN | TCGA | RHUH | Mean | SD | Min | RMSE (days) |
|---|---|---|---|---|---|---|---|---|---|
| Age + Sex + W.L-TDI | Cox | 0.646 | 0.614 | 0.649 | 0.641 | 0.637 | 0.016 | 0.614 | 451.0 |
| Age + Sex + W.L-TDI | Weibull AFT | 0.646 | 0.613 | 0.647 | 0.644 | 0.637 | 0.016 | 0.613 | 402.9 |
| Age + Sex + W.L-TDI | LogLogistic AFT | 0.644 | 0.614 | 0.648 | 0.644 | 0.638 | 0.016 | 0.614 | 518.7 |
| Age + Sex + E.L-TDI | Cox | 0.620 | 0.616 | 0.631 | 0.610 | 0.619 | 0.009 | 0.610 | 472.2 |
| Age + Sex + E.L-TDI | Weibull AFT | 0.620 | 0.615 | 0.628 | 0.613 | 0.619 | 0.007 | 0.613 | 420.8 |
| Age + Sex + E.L-TDI | LogLogistic AFT | 0.627 | 0.618 | 0.631 | 0.630 | 0.626 | 0.006 | 0.618 | 537.2 |
| Age + Sex + E.size | Cox | 0.631 | 0.606 | 0.636 | 0.674 | 0.637 | 0.028 | 0.606 | 433.1 |
| Age + Sex + E.size | Weibull AFT | 0.629 | 0.606 | 0.639 | 0.677 | 0.637 | 0.030 | 0.606 | 390.6 |
| Age + Sex + E.size | LogLogistic AFT | 0.632 | 0.606 | 0.634 | 0.666 | 0.634 | 0.025 | 0.606 | 513.5 |
| Age + Sex | Cox | 0.635 | 0.602 | 0.626 | 0.623 | 0.621 | 0.014 | 0.602 | 455.1 |
| Age + Sex | Weibull AFT | 0.635 | 0.602 | 0.626 | 0.634 | 0.624 | 0.015 | 0.602 | 406.2 |
| Age + Sex | LogLogistic AFT | 0.632 | 0.602 | 0.626 | 0.623 | 0.621 | 0.013 | 0.602 | 533.1 |

#### 6. Files

Full-precision tables: LTDI-Volume_comparison_strata-none_standardized-true_coefficients.csv, LTDI-Volume_comparison_strata-none_standardized-true_model-fit.csv, LTDI-Volume_comparison_strata-none_standardized-true_cross-validation.csv. Run log: LTDI-Volume_comparison_strata-none_standardized-true_log.txt. Per-model forest plots: <PreIPost>surgical_<marker>_strata-none_standardized-true.pdf/svg. All in /home/joan/Desktop/PROJECTS/Glioblastomas/RESULTS-GBM_4-cohorts_Tissues/Forest-plots_Cox-models.
